# In Vivo CRISPR Gene Editing in Patients with Herpes Stromal Keratitis

**DOI:** 10.1101/2023.02.21.23285822

**Authors:** Anji Wei, Di Yin, Zimeng Zhai, Sikai Ling, Huangying Le, Lijia Tian, Jianjiang Xu, Soren R Paludan, Yujia Cai, Jiaxu Hong

## Abstract

In vivo CRISPR gene therapy holds large clinical potential, but the safety and efficacy remain largely unknown. Here, we injected a single dose of HSV-1-targeting CRISPR formulation in the cornea of three patients with severe refractory herpes stromal keratitis (HSK) during corneal transplantation. Our study is an investigated initiated, open-label, single-arm, non-randomized interventional trial at a single center (NCT04560790). We found neither detectable CRISPR-induced off-target cleavages by GUIDE-seq nor systemic adverse events for 18 months on average in all three patients. The HSV-1 remained undetectable during the study. Our preliminary clinical results suggest that in vivo gene editing targeting the HSV-1 genome holds acceptable safety as a potential therapy for HSK.

**One-Sentence Summary:** Our study is the first in vivo CRISPR therapy for treating infectious disease and the first virus-like particle (VLP)-delivered gene therapy, reporting clinical follow-up to 21 months in HSK patients without seeing virus relapse, HSK recurrence, and CRISPR-associated side effects.

## Introduction

Herpes simplex virus type 1 (HSV-1) is a common human virus, with a global seroprevalence of 50–90% (*1-3*). Ocular HSV-1 infection is the major cause of herpetic stromal keratitis (HSK) which is one of the leading causes of infectious blindness in developed countries (*4*). Approximately, 40,000 people develop visual disability among the 1.5 million new cases of ocular HSV infection each year (*4*). After primary infection in the cornea, HSV-1 establishes a latent reservoir in the trigeminal ganglia (TG), which can be reactivated, leading to the recurrence (*5*).

Currently, no vaccine is available against HSV infection (*6*). Acyclovir (ACV) and analogs that target the viral DNA polymerase are the first-choice treatments for HSK (*7*). However, as the antiviral molecules do not change the existing viral DNA, recurrences are still common. Additionally, resistance to ACV has been associated with longer disease duration and subsequent failure of prophylaxis in patients with recurrent ocular HSV episodes (*7, 8*). Other strategies, including antibodies, peptides, and small molecules are still under development (*9*). Corneal transplantation is recommended for HSK-induced corneal leucoma or perforation. However, the postoperative prognosis is compromised by high rates of virus recurrence (*10*). According to previous studies, there is a high incidence of herpetic keratitis recurrence following penetrating keratoplasty (PK) - even with the topical acyclovir, the recurrent rate is still over 55% (*10, 11*). Various studies have reported the recurrence rate of HSV infection in grafts with prophylactic oral acyclovir therapy ranging from 12% to 33% (*11-13*). Notably, systemic ACV has been associated with kidney injury and neurotoxicity (*11, 14*). Essentially, neither the drugs nor surgical treatments can diminish the virus in the corneas or TG reservoir and cleave the virus genome directly, which drives the exploration of next-gen antiviral strategies with meganucleases and CRISPR (*15-17*).

CRISPR has been remarkably successful in preclinical research (*18-22*). So far, it has been applied in clinical trials for treating hemoglobin diseases, cancers, and HIV infection (*23-27*). However, the published trials are all ex vivo except the recent report by Gillmore et al. in which CRISPR was used for in vivo treatment of transthyretin amyloidosis (*28*). Still, the broader in vivo safety and efficacy profile of CRISPR remains to be established. To overcome the delivery obstacle for in vivo CRISPR therapy, several groups including ours have reported engineered virus-like particles which are able to transport mRNA or ribonucleoprotein, allowing transient gene editing (*29-31*). Our group has developed an mRNA-carrying lentiviral particle (mLP), capable of incorporating mRNA of interest such as SpCas9 in the producer cells via the interaction between aptamer in the mRNA and adaptor in the viral structural protein (*32*). We further designed the SpCas9 mRNA-carrying lentiviral particle, termed HSV-1-erasing lentiviral particle (HELP), to specifically cleave two essential genes for the HSV-1 life cycle, UL8 and UL29, for HSK therapy (*33*). A single intracorneal injection of HELP significantly diminished HSV-1 in the corneas and TG in mice (*33*).

Although we have shown HELP efficiently controlled the HSV replication in vitro, in animal models and ex vivo human corneas, it may be difficult to clear all the viruses in the reservoir in humans. We hypothesized that reducing the virus load to a certain threshold will change the balance between virus and host so that the immune system can control the remaining virus and achieve a functional cure (*34, 35*). Here, we reported data from a clinical trial evaluating the safety and efficacy of one dose HELP injection during the penetrating keratoplasty for three patients with severe refractory HSK and acute corneal perforation (Figure 1), all three had completed the 12 months follow-up (21 months,18 months and 14 months for patient 1, 2 and 3, respectively).

**Fig. 1.**
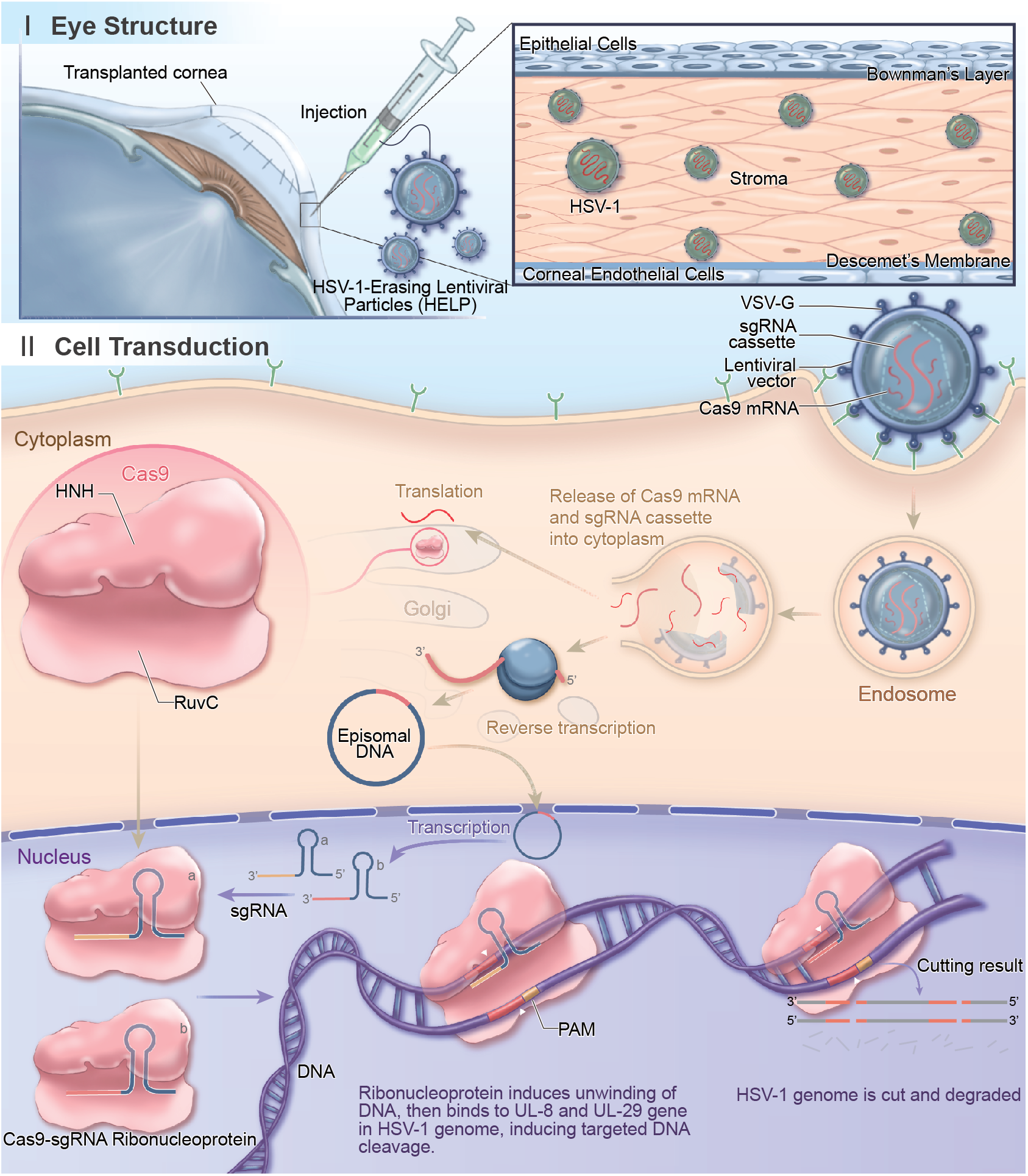
The Antiviral Mechanism of CRISPR in human corneas. HELP is a lentiviral viral particle carrying SpCas9 mRNA and a gRNA-expression cassette. The SpCas9 protein translated from mRNA directly and the gRNA expressed from the reverse transcribed viral DNA assemble into RNPs which cleave the HSV-1 genome at UL8 and UL29 loci for degradation. A total of 0.2 mL (2.4 μg p24) HELP formulation was injected into the recipient graft bed in 6-8 locations by using a 27g ophthalmic syringe after corneal transplantation.

## Results

### Preclinical Results

The presence of HSV-1 in the corneas of the participant was determined by quantitative polymerase chain reaction (qPCR) analysis of the viral genome in tear-swabbed samples. The positive result was a precondition for enrollment (Table 1). HELP was given by intrastromal injection at a dose of 2.4 μg p24 during penetrating keratoplasty illustrated in Fig. 1. The perforated cornea was removed from the patient during the operation for subsequent fluorescence microscopy and immunohistochemistry analysis. The viral capsid protein VP5 was detected in the corneas of three patients by confocal imaging (Extended Figure 1). As HSK is the consequence of excessive virus-induced corneal infiltration of inflammatory cells (*36*), we therefore stained the removed corneal button for T cells (CD4+ and CD8+), myeloid-derived cells (CD11b+) and macrophages (F4/80+). Immunohistochemistry showed their infiltration in the cornea stroma of three patients (Extended Figure 2). We sequenced HSV-1 strains isolated from three participants for genes encoding thymidine kinase (TK) and DNA polymerase (DNA pol). We found changes in amino acids in TK or in DNA pol, but they were not drug-resistance mutations (Figure S1 from Supplemental Information). Additionally, we used a HSV-1 infected healthy cornea to evaluate the functionality of CRISPR and found a significant reduction of HSV-1 genome and viable viruses in addition to the bare detectable VP5 antigen (Figure 2A-E). Also, we found direct evidence of viral genome cleavage in UL8 and UL29 loci with indel frequencies 28.8% and 14.4%, respectively, by deep sequencing (Figure 2F).

**Table 1.** HSV-1 Diagnosis^*^.

| No. | Sex | Age range | Cornea | Aqueous | Tear swab (time post injection) |  |  |  |  |  |  |  |
| --- | --- | --- | --- | --- | --- | --- | --- | --- | --- | --- | --- | --- |
|  |  |  |  |  | -1 day | 1 week | 1 month | 2 months | 3 months | 6 months | 12 months | Last visit<sup>#</sup> |
| 1 | M | 70-74 | + | + | + | - | + | - | - | - | - | - |
|  |  |  |  |  | + | + | + | - | - | - | - | - |
|  |  |  |  |  | + | - | + | - | - | - | - | - |
| 2 | M | 50-54 | + | +/- | + | - | - | - | - | - | - | - |
|  |  |  |  |  | + | - | - | - | - | - | - | - |
|  |  |  |  |  | + | - | - | - | - | - | - | - |
| 3 | M | 65-69 | + | - | + | - | - | - | - | - | - | - |
|  |  |  |  |  | - | - | - | - | - | - | - | - |
|  |  |  |  |  | - | - | - | - | - | - | - | - |

\* HSV-1 diagnosis was via nucleic acids detection by quantitative polymerase chain reaction.
<sup>#</sup> Last visit: patient 1 = 21 months, patient 2 = 18 months, patient 3 = 14 months.
+, positive, Ct value $\leq 36$ .
+/-, weak positive, 36<Ct value<40.
-, negative, Ct value = 40 or “no Ct”.

**Fig. 2.**
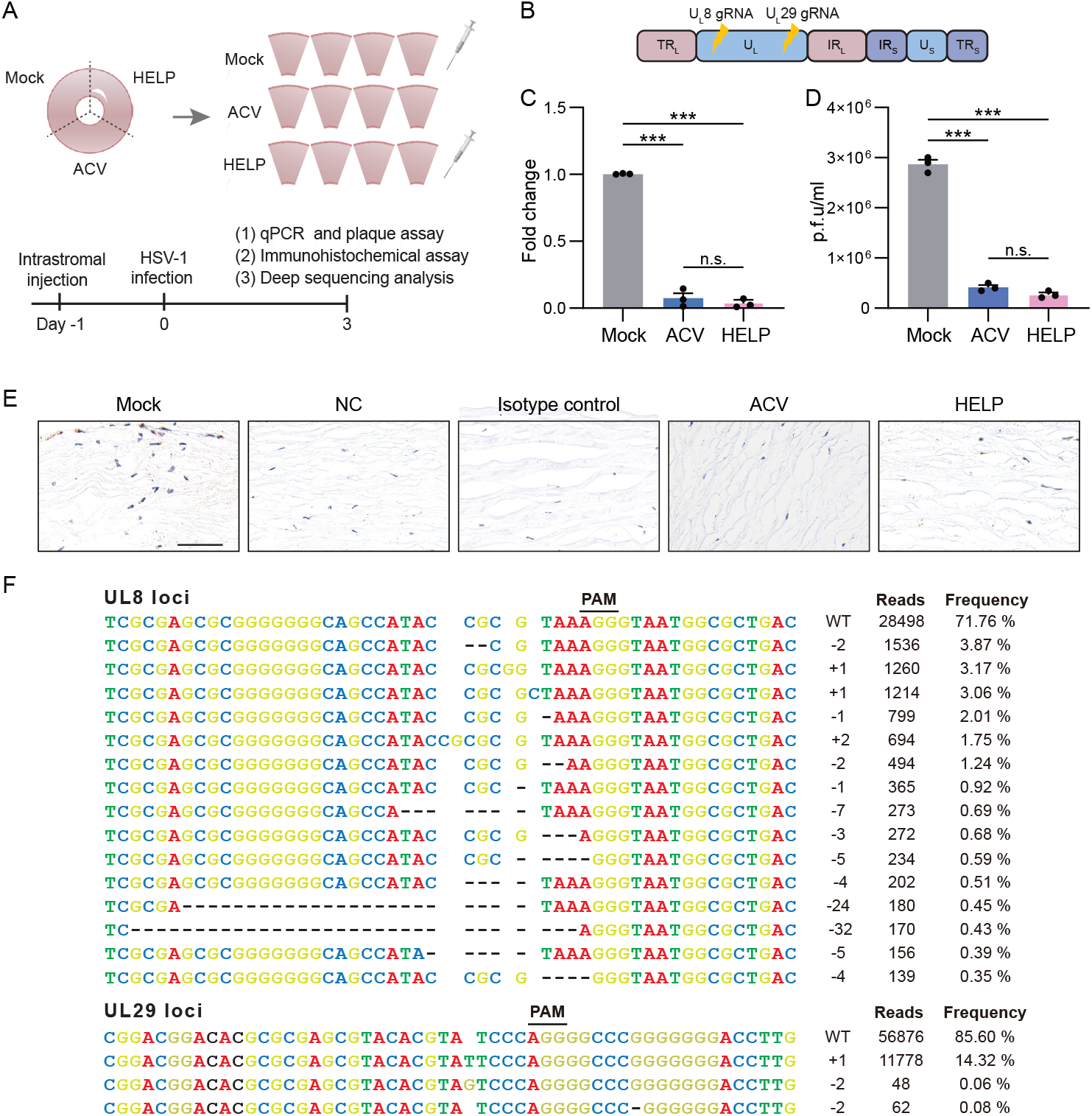
Preclinical results of HELP eliminating HSV-1 in tissue culture of a healthy cornea. **(A)** Flowchart for evaluating the antiviral effects of HELP in HSV-1 infected healthy human cornea. (**B**) Schematic illustration of the HSV-1 genome and gRNA loci. TRL, terminal repeat long; UL, unique long; IRL, internal repeat long; IRS, internal repeat short; US, unique short; TRS, terminal repeat short. (**C**) qPCR analysis of HSV-1 genome (fold change); P<0.0001; (**D**) HSV-1 titer of supernatants from human corneal cultures measured by plaque assay; mock versus AVC and HELP, P<0.0001, respectively. (**E**) Immunohistochemistry analysis of HSV-1 dissemination in a human cornea; scale bar, 20 μm. (**F**) The profile of HELP-induced mutations in HSV-1 UL8 and UL29 loci in a human cornea. Frequencies>0.035 and 0.07% in UL8 and UL29 loci were shown, respectively. The minus symbol indicates a deletion and the plus symbol indicates an insertion. PAM, protospacer adjacent motif; WT, wildtype. Data and error bars represent mean ± s.e.m.; n.s, not significant. One-way ANOVA with Dunnett’s multiple comparisons test was performed.

### Patient Demographics and Outcomes

#### Patient 1

Patient 1 was a senior male in his early 70s who was admitted in November 2020 for corneal perforation due to recurrent HSK in the right eye and had initial HSK nearly 30 years ago with a recurrence interval of every 1-2 years. He had been prescribed a high dose of antiviral medications yet still failed to ameliorate the keratitis. His left eye was diagnosed with traumatic corneal leucoma. On examination, uncorrected visual acuity was light perception for both eyes. The slit-lamp image of the right eye showed an ill-defined central corneal ulcer of about 3 mm×3 mm with iris incarcerated in the perforation. The left eye showed a cloudy cornea without conjunctival congestion. Patient 1 had mild anemia and a history of diabetes with a slightly high glycated hemoglobin of 6.2%.

Topical ganciclovir eye gel was discontinued for the patient one day before the uncomplicated PK. During the PK operation, the iris was carefully separated and severe lens opacity was found in the surgical eye. Patient 1 received HELP injection according to the protocol (Figure 3A). Topical 0.5% levofloxacin and 1% prednisolone acetate were administered three times per day after the surgery. Postoperative hyphema was identified immediately after the surgery, which was absorbed on day 7. His visual acuity of the right eye has remained finger count owing to the severe cataract since day 3. At 6 months after PK, patient 1 developed an uncontrolled corneal ulcer related to neurotrophic keratopathy (NK), which mainly resulted from the long-lasting damage to the corneal sensory nerve induced by chronic HSV infection. In accordance with his decrease in corneal sensitivity, in vivo confocal microscopy (IVCM) showed significantly impaired corneal subbasal nerves in the central cornea graft at the latest follow-up (Extended Figure 3). To further improve the patient’s vision, we therefore conducted the second corneal transplantation and phacoemulsification with intraocular lens implantation on his right eye. He had regained an uncorrected visual acuity (decimal) of 20/100 in the surgical eye by the time of discharge (Figure 3B). The postoperative intraocular pressure was within the normal range (Table S1 from Supplemental Information). Optical coherence tomography (OCT) showed a relatively shallow anterior chamber in patient 1 (Extended Figure 4). His corneal graft had remained transparent since the second PK after 12 months post-injection (Figure 3A).

**Fig. 3.**
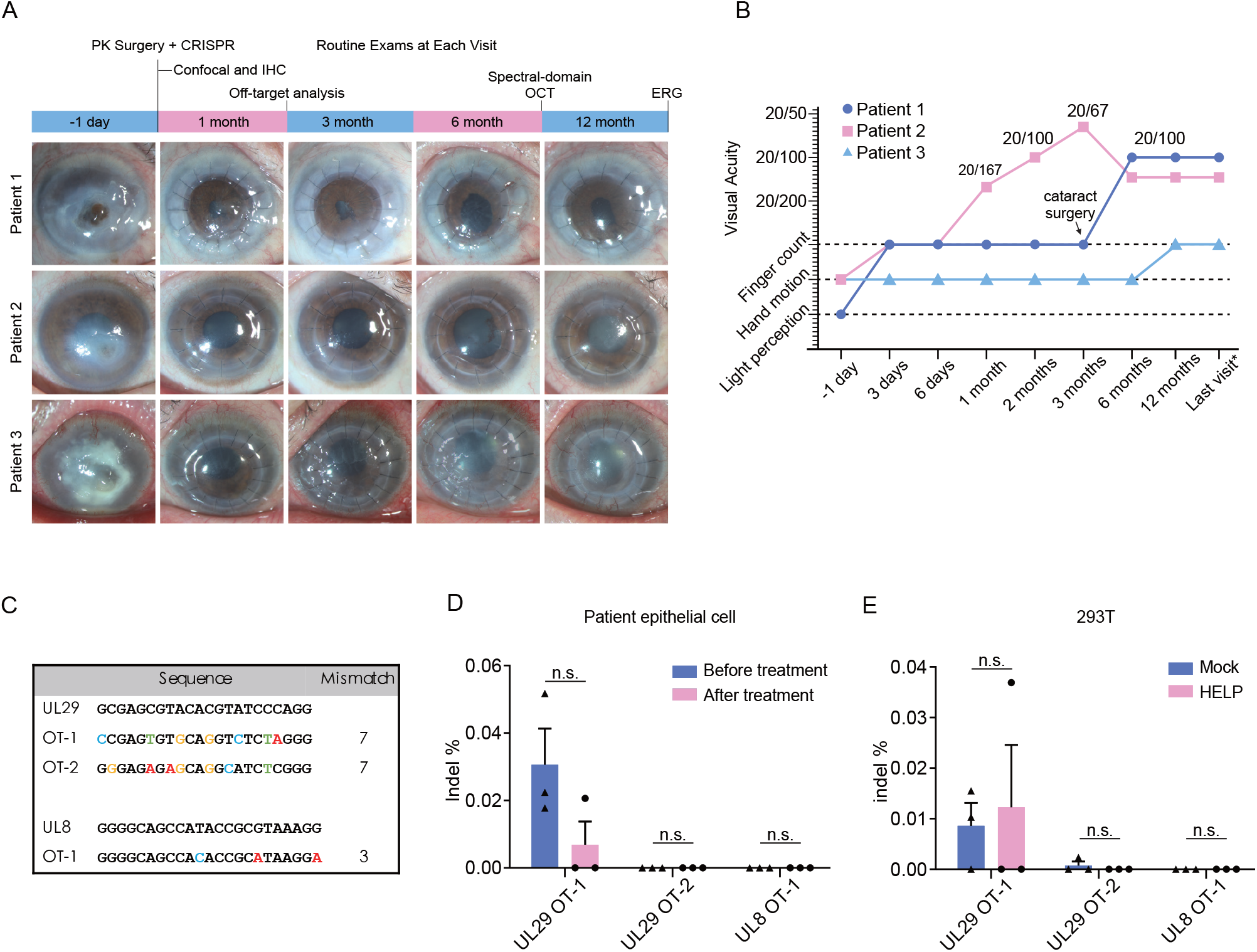
The Main Clinical Results of CRISPR-treated HSK patients. (**A**) The time scheme and corresponding anterior segment photographs of three participants. (**B**) The changes of visual acuity in each participant during follow-ups, wherein last visit means 14 months, 18 months and 21 months for patient 1, 2 and 3, respectively. (**C**) Potential off-target loci of HELP in the human genome identified by the GUIDE-seq. (**D**) Deep sequencing results on the potential off-target sites in the patients’ corneal epithelial cells at 1 month post HELP treatment. (**E**) Deep sequencing results on the potential off-target sites in 293T cells. The genome was isolated 48 h post HELP transduction. OT represents off-target site. Data and error bars represent mean ± s.e.m.; n.s., non-significant; unpaired two-tailed Student’s t-tests.

Before treatment, both the corneal button and aqueous humour were diagnosed positive for HSV-1 using a Triplex HSV-1 DNA Diagnostic Kit with cycle threshold (Ct) values of 21.35 and 28.37, respectively (Table S1). However, tear swabs have maintained negative since his 2-month visit (Table 1). Interestingly, the second corneal transplantation in the sixth month gave us a unique opportunity to explore whether the HELP can diminish the HSV-1 genome in vivo by examining the excised corneal button. Indeed, we found VP5 signals neither around the rim nor in the center of the removed corneal button via fluorescence microscopy and immunohistochemistry analysis (Extended Figure 5 and 6).

#### Patient 2

Patient 2 was a male in his early 50s who had an acute corneal perforation in the right eye caused by recurrent HSK. His right eye had been suffering from repeated redness and pain for more than a decade with a recurrence interval of 2-3 months, which could not be effectively controlled by various antiviral and glucocorticoid eye drops. Five days before admission in January 2021, the patient complained of sharp, sudden pain and fluid leakage in the right eye. On examination, uncorrected visual acuity (decimal) was 20/20 in the left eye yet only hand motion in the right eye. The slit-lamp image of the right eye revealed a 4 mm-diameter lower paracentral corneal ulcer with a pinpoint leakage but no hypopyon in the anterior chamber. The lens of the right eye was slightly cloudy. His left eye was unremarkable. Patient 2 had a history of chronic B-related hepatitis and hyperlipidemia.

Uncomplicated PK was performed for patient 2 who also received HELP injection subsequently according to the protocol. After the surgery, topical 0.5% levofloxacin and 1% prednisolone acetate were applied three times a day whereas the traditional acyclovir eye drops (every two hours) and oral tablets (twice daily) were discontinued. One day after surgery, topical prednisolone acetate was increased to four times daily due to corneal graft edema. Two days later, intraocular pressure of the right eye was 11 mmHg and visual acuity raised to finger count. OCT at this time confirmed a flat iris and open anterior angle in all directions (Extended Figure 7). At later visits, his visual acuity improved to 20/167 immediately at one month visit and stabilized as 20/67 at 3 months after the surgery (Figure 3B). In the sixth month after PK, patient 2 was diagnosed of Staphylococcal endophthalmitis which was confirmed by bacterial and fungal cultures (Table S2) and subsequently contained by the combination of vitrectomy and antibiotic injection. His uncorrected visual acuity had decreased to 20/133 because of the secondary cataract in the surgical eye (Figure 3B). IVCM revealed partial regeneration of corneal nerves at his 12 month-follow-ups (Extended Figure 8). No relapse of herpetic keratitis or graft rejection was identified at the final visit.

Before treatment, the patient was positive for HSV-1 by qPCR analysis of the removed corneal tissue (Ct=26.34) and tear swabs (Ct=35.01), and the aqueous humour which was weak-positive (Ct=36.50) (Table S1). In the subsequent 12- and 18-month follow-ups, however, we found the tear swabs samples during the postoperative visits showed that the HSV-1 tests were all negative by qPCR examination (Table 1).

#### Patient 3

Patient 3 was a male in his late 60s who had six-year of HSK and eventual corneal perforation in the left eye 10 days before admission in May 2021. His HSK relapsed every 2-3 months. The same eye had phacoemulsification with intraocular lens implantation 8 years ago. His right eye was unremarkable. On examination, uncorrected visual acuity (decimal) was 20/50 in the right eye and hand motion in the left eye. Conjunctiva congestion and limbal neovascularization were severe in his left eye, with a gray ulcer of 5 mm×5 mm at the center of the opaque cornea. Subacute cornea perforation and hypopyon indicated a high-risk corneal transplantation. OCT confirmed partial perforation on the cornea (Extended Figure 9). Patient 3 had a history of hypertension.

After the uneventful PK and HELP injection, the patient had been prescribed 0.3% tobramycin and 0.1% dexamethasone ophthalmic ointment four times daily. Brinzolamide timolol drops (twice per day) were prescribed for his elevated intraocular pressure owing to postoperative hyphema. Ganciclovir eye gel (four times daily) and acyclovir oral tablets (twice daily) were discontinued after treatment. One day after the surgery, 0.5% cyclosporine eye drops were added to suppress inflammation of the ocular surface. Due to the intraocular lens opacity induced by corneal perforation, the visual acuity of patient 3 had been hand motion since PK operation and finger count at 12-month (Figure 3B). On his third month follow-up, we observed corneal edema and inflammatory cell infiltration around the corneal sutures, indicating an occurring graft rejection which was controlled after receiving topical treatment for the graft rejection and secondary glaucoma. Postoperative B-scan ultrasonography suggested no significant abnormal changes in the retina (Figure S2).

Though we did not detect HSV-1 in aqueous humour in patient 3 before CRISPR treatment, the patient was diagnosed as HSV-1 positive for detecting HSV-1 in the tear swab (Ct=29.5) and the removed corneal tissue (Ct=34.9) during PK by qPCR, and the viral VP5 antigen using confocal imaging (Table 1, Table S1 and Extended Figure 1). However, in the 12- and 14-month follow-ups, we found the swab samples during the postoperative visits were all negative for HSV-1 (Table 1).

### Additional Safety Assessment

The potential HELP (CRISPR)-relevant side effects are the immune response attacking the CRISPR-transduced cells expressing Cas9 and off-target cleavages. ELISA showed intrastromal injection of HELP did not provoke an anti-vector immune response as no p24-specific IgG was induced (Extended Table 1). Interestingly, by examining the blood samples, all the patients were SpCas9-positive before treatment while the Cas9-specific IgG was not significantly enhanced after treatment (Figure S16). To analyze the potential off-target effects of our virus-targeting HELP in the human genome, we performed GUIDE-seq to reveal genome-wide integrations of double-stranded oligodeoxynucleotides in the double-strand DNA breaks caused by CRISPR (Figure 3C) (*40*). However, in subsequent deep sequencing, we did not detect indels on the three GUIDE-seq identified sites in the detached epithelial cells from wiping the cornea of the patients 1 week after the treatment as well as in 293T cells (Figure 3D, E). For the rest, we performed a complete ophthalmic examination on three patients after HELP treatment (Supplementary Information). The ocular adverse events (AE) we observed included corneal graft edema, hyphema, post-herpes NK, concurrent cataract and secondary glaucoma, which were mainly attributed to the high-risk penetrating keratoplasty with acute corneal perforation (*37-39*) (Table S2). B-scan ultrasonography of the patients on 7 days, 1 month, 6 months and 12 months post-injection showed no abnormal changes in the vitreous body and retina (Figure S2-S4). Retinal OCT demonstrated relatively intact fundus in HELP-injected eyes (Figure S5, S6). Additionally, we found no apparent change in rod or cone responses in the participants by the electroretinography (ERG) (Figure S7-S9). No systemic AEs were found during the study period.

## Discussion

We report three refractory HSK cases with acute corneal perforation receiving in vivo CRISPR therapeutics. After a single intrastromal injection of HELP in combination with corneal transplantation, corneal grafts and tear swabs in the 3 patients were still free of viral relapse at their last visits with the average follow-ups reaching 18 months even though we discontinued the antiviral therapy after the CRISPR injection. In the HSV-1 infected healthy corneas, HELP efficiently diminish the HSV-1 in terms of genome, viable viruses and antigen (Figure 2). Moreover, we did not detect HSV-1 in the removed corneal button from patient 1 at 6 months post HELP-treatment due to neurotrophic keratopathy, adding supporting piece that the HELP could diminish the HSV-1 in the corneas (Extended Figure 5). The laboratory data suggested no CRISPR off-target cleavage in the human genome or Cas9 and vector-specific immune response induced by intrastromal injection of HELP. These results suggest that HELP might be an effective strategy in restricting HSV-1 replication in the human corneas with no remarkable CRISPR-related side effects.

Although the HSV-1 DNA levels became undetectable immediately after treatment in patient 2 and 3, the virus was still detected in the first month for patient 1, but not at later time points. We reason several mechanisms may be responsible for this phenomenon. It is likely that HELP reduces viral load by targeting both productive replication and the latent reservoir without achieving full elimination, especially in cases like patient 1 who had a much higher virus load than patient 2 and patient 3 (Table S1 and Extended Figure 1). Therefore, the acute stress-associated host response is likely to augment viral replication (*41, 42*). Second, immune activities are also likely required to fully inhibit the virus after the HELP treatment. Patient 1 was in his seventies and therefore might have a relatively weak immune system not fully capable of controlling residual viruses in the corneas and TG the first months after treatment. To balance the benefit-risk of the first antiviral CRISPR therapy, patient 1 was chosen because he had very high preoperational viral loads in the cornea, aqueous humour and tear swabs. Therefore, it is not surprising that HELP was unable to clear the virus completely in this patient, but only reduce the virus burden in vivo. However, as the cornea of patient 1 was transparent at the one-month visit without signs of recurrence, we continued antiviral drugs-free protocol on the patient. Notably, his swabs of both cornea and tear film remained negative for HSV-1 from 2-month to the final visit. According to the current study, patient 2 and 3, who had lower pre-operational HSV-1 load compared with patient 1, had obtained a more thorough HSV-1 clearance by HELP judging from their virus tests. Future studies enrolling more patients with different degrees of HSK will answer whether patients with lower pre-operational corneal virus load than the three patients in this study will have an even more beneficial outcome after HELP treatment.

The participants enrolled in our study were all experiencing recurrent HSK episodes for decades, raising the possibility of drug-resistant HSV-1 infection. Accordingly, we found respective rather than simultaneously changes of amino acids in TK and DNA pol in the patients’ isolates that were different from both the KOS and 17syn+ strains of HSV-1 (Figure S1), suggesting that the constant release of HSV-1 from the reservoir, instead of drug resistance, may have contributed to the refractory HSK in these patients. To date, the HSV-1 reservoir in TGs has still been untargetable with antiviral drugs. We have previously shown in the preclinical model that HELP is capable of modulating the virus in the reservoir via retrograde transportation of CRISPR to TG (*33*). High-risk corneal transplantation for the perforated cornea is prone to HSK recurrence and corneal rejection. Therefore, the discontinuation of antiviral therapy after PK could make participants more vulnerable to HSV-1 recurrence than other patients. However, among the 21 tests of HSV-1 on three participants, except 2 tests in the first month of patient 1 were positive without any clinical symptoms, all the rest tests with the longest follow-up up to 21 months after treatment were negative. Thus, all three participants had no HSK recurrence during this study although the specific role of HELP on HSV-1 in the human TG reservoir remains elusive.

We summarized in a table for all the adverse events divided by the patients and by follow-up visits (supplementary information Table S2). Although HELP treatment may not be fully excluded from the observed complications due to the limited cases and lack of controls, this was very unlikely except for corneal edema. Numerous complications, such as graft rejection, hyphema, glaucoma, or cataract could be found in postoperative treatment of penetrating keratoplasty despite the progress of surgical techniques, especially in the high-risk cases with acute corneal perforation (*37*). Hyphema in Patient 1 is mainly attributed to the tear of the iris during the surgery, which finally resolved. Cataract in our three cases may happen because of acute corneal perforation and the usage of corticosteroid after the surgery. The incidence of glaucoma in Patient 1 and 3 is not rare after corneal transplantation. Several etiologic factors have been identified, the most common being synechial angle closure and corticosteroid-induced IOP elevation. IVCM revealed HSV-related corneal epithelium abnormality and low nerve density in all three participants, indicating their high susceptibility to post-herpes NK. Damage to the sensory nerve ending of the trigeminal nerve by HSV-1 could result in decreased corneal sensitivity, loss of epithelial integrity and corneal ulceration in severe cases (*43*). This possibly could explain why patient 1, a senior male with 30 years of HSK history ultimately developed NK 6 months after PK. The endophthalmitis of Patient 2 stemmed from bacterial infection, identified as Gram-positive cocci (*Staphylococcus caprea*) using bacterial culture. The HELP product was produced and stored under GMP-like regulation, with a quality control report (Table S14). Additionally, the HELP was injected into the cornea of Patient 2 which remained transparent during the 18-month follow-up. To sum up, the potential HELP-relevant clinical AEs were minimal after CRISPR treatment.

Immune responses against the viral vector or Cas9 can be problematic for in vivo CRISPR gene therapy (*44, 45*). In our study, no vector-specific IgG was detected after the HELP treatment. Indeed, prednisolone acetate has been prescribed throughout the entire follow-up course of the patients as instructed by the standards for corneal transplantation to prevent rejection, which may have played a role in the restrained inflammatory and immune reactions in the patients (*9*).

Additionally, we have previously characterized that HELP is low immunogenic (*33*), therefore, both the low immunogenicity and prednisolone acetate may have contributed to the absence of inflammatory and immune reactions after the HELP treatment. Finally, the lack of HELP-related adverse effects may also be due to the transient nature of SpCas9 mRNA delivered by HELP.

HELP was originally designed for a single dosage administration. In the case of repeated dosing, the effectiveness of HELP may be compromised if the vector-specific IgG antibodies were induced, as they may block HELP at the entry level. However, the intracorneal injection may not be an effective route to induce anti-vector IgG as all three patients were p24 negative after HELP treatment (Extended Table 1). Additionally, repeated administration may not cause safety problems for HELP as the SpCas9 expressed from mRNA exists only for several days, thus minimizing the chance to be attacked by cytotoxic T cells (*32*).

In conclusion, our clinical results from three HSK patients with an average of 18-month follow-up after receiving HELP suggest that in vivo CRISPR gene editing targeting the HSV-1 genome holds promise as a safe antiviral adjuvant therapy for which the efficacy may further be improved in combination with existing antiviral molecules such as ACV for severe refractory HSK to prevent the virus recurrence (*46*). Dose escalation may clear the virus in high virus-load patients more thoroughly than was achieved in patient 1. As our study is a single-arm pilot trial, a more complete future study including a head-to-head comparison to the conventional ACV treatment is necessary to fully evaluate the safety and efficacy of in vivo CRISPR therapy.

Future studies may also expand the treatment to patients with less severe HSK to protect them from getting refractory HSK and corneal perforation complications. Overall, our study provides clinical proof-of-concept for in vivo CRISPR gene editing as a potential antiviral strategy.

## Materials and Methods

### Study Oversight

The study is designed to evaluate the safety and efficacy of HELP for in vivo treatment of refractory HSK patients. This clinical practice was approved by the ethics committee of the Eye & ENT Hospital of Fudan University, China, the National Health Commission, China (IRB-220113) and registered with ClinicalTrials.gov (NCT04560790). Written informed consent was provided by the patients under video recording. The healthy donors involved in this study all signed the informed consent for biological sample donation and data sharing before the operation, with an agreement that the samples should be used for biomedical research. All the authors had access to the data and vouch for their accuracy and completeness. The manuscript was written and revised by the corresponding authors with the help from all authors and was approved for publication by each of the authors.

### Trial Design and Eligibility

In this investigated initiated, open-label, single-arm, non-randomized interventional trial of 12 months at a single center, participants with refractory HSK were designed to receive a single dose of HELP formulation via intrastromal injection during the penetrating keratoplasty for acute corneal perforation. The trial is a completed analysis for this pilot study. Adult (ages 18 to 70) patients with refractory keratitis who were repeatedly infected with HSV-1 virus (more than three times per year) and unresponsive to the standard regimen for HSK (oral Acyclovir, 800 mg, bid for 10 days, then 400 mg, bid for at least 1 year); or those who were suffering from relapse HSV infections with corneal perforation requiring corneal transplantation were enrolled. Patients who had serious ocular or systemic comorbidities (i.e., trauma, other infections, immune diseases, tumors) judged by investigators that were unsuitable for the trial were excluded from this study. The follow-up strategy of this study consisted of two stages, a treatment period (1 day to 12 months) and a monitoring period (1 year to 15 years). Participants entered the treatment follow-up period on the surgery day after operation, from which researchers monitored predefined indicators according to the study protocol. After the end of the clinical trial (12 months post-injection) patients transitioned into the monitoring period. Every 12 months researchers would appoint a follow-up visit or phone call to acquire the latest data of patient status, continuing for 15 years or until the death of participants.

Compared with the original protocol, the major revisions on the current one were listed as adjusting the contents and priorities of outcomes and modifying the inclusion criteria (Supplementary Table S14). These revisions will not increase the risk to subjects and have been approved by the Ethics Committee. From April 2020 to April 2022, a total of six HSK patients had provided informed consent: two did not meet eligibility criteria, one withdrew before undergoing HELP injection on their own decision, and three received intervention and had >12 months of follow-up; the preliminary analysis of our study was based on these last three patients (Trials Flow Diagram from supplementary material).

### Main Outcomes and Safety

The primary outcome was the ocular surface HSV-1 of the intervention eye on 1 week, 1 month, 2 months, 3 months, 6 months, 12 months post-operation and serious AEs post-injection. The secondary outcome was cornea graft survival and best corrected visual acuity (BCVA) at every follow up visit. Safety assessment included HELP-provoked humoral immune reaction, off-target effects of CRISPR/Cas9 technology, tolerability assessments based on HELP-related AEs, clinical ophthalmic examinations and vital signs, and all-cause mortality. Owing to the small sample size (n=3), the data are primarily descriptive. More details are provided in the Supplementary Information. Routine assessments included a complete ophthalmic examination (measurement of BCVA, and intraocular pressure; slit-lamp examination), IVCM to analyze the corneal structures at the cellular level (corneal epithelium/endothelium and corneal subbasal nerves), time-domain OCT to investigate the anterior segment, the spectral-domain OCT to document the retinal status, postoperative B-scan ultrasonography to evaluate the status of the posterior segment of the eye, the and the ERG to measure the electrical activity generated by the retina in response to a light stimulus. Aqueous humour and the surgically removed corneal button were collected for HSV-1 genome detection. Tear swabs of pre-injection and 1 week, 1 month, 2 months, 3 months, 6 months and 12 months post-injection were also collected and quantified for HSV-1 genome by qPCR using HSV-1 DNA Diagnostic Kit (Triplex International Bioscience). The virus testing outcome was indicated as positive (+) or negative (-) by Ct values that are equal or above 40 for negative, 36-40 for weakly positive, and, equal or below 36 for positive according to the manufacturer’s instruction. Blood samples of the participants were collected at 1-month, 3-month, 6-month and 12-month visits, to confirm whether intrastromal injection of HELP induces SpCas9-specific IgG and vector-specific IgG which were determined by ELISA (*44*). Deep sequencing of the detached epithelial cells by wipes was used for analyzing the potential off-target effects caused by CRISPR in the human genome.

### Preparation and administration of HELP

The SpCas9 mRNA-carrying HELP was produced and quality controlled following Good Manufacturing Practice (GMP) guidelines by OBiO Technology. To manufacture the GMP-like level HELP, 293T cells were seeded in cell factories 24 h before PEI transfection. Six hours after transfection, the media were refreshed and the supernatants were harvested 48 h post-transfection. The virus-rich supernatants were filtered through a 0.45 μm filter and treated with Benzonase. After ultrafiltration/diafiltration using tangential flow filtration, the viral particles were purified by chromatography and then passed through 0.22-μm filters for sterilization.

The final formulation of HELP is a phosphate buffered saline (PBS)-dissolved lentiviral vector which contains two types of RNA ‘all-in-one’ – one is SpCas9 mRNA (4 copies on average in each particle) and the other is a gRNA-expression viral RNA cassette *(33)*. A D64V mutation was introduced in the integrase so that the gRNA-expression cassette will not integrate into the host genome. The gRNA expression cassette is reverse transcribed in the host cells and maintained as circular episomal DNA which encodes two gRNAs targeting UL8 and UL29, respectively. The formulation was kept at -80°C for long-term storage. It was recovered to room temperature 10 min before injection. Each formulation, 0.2 mL in total, was drawn into a 27g ophthalmic syringe for the injection. Penetrating keratoplasty was performed following the routine procedures. After sewing up the donor cornea, the graft bed of the recipient was injected with the HELP formulation in 6-8 locations (Figure 1 and Supplementary Surgical Videos).

### HSV-1 plaque assay

Vero cells were seeded in a 12-well plate and infected with various dilutions of culture supernatants on the following day. Two hours after infection, cells were overlaid with 1% agarose solution. The cells were cultured for 3 days before being fixed with 4% formaldehyde and stained with 1% crystal violet solution. After three washes with water, the number of plaques was counted. HSV-1 titers were calculated as p.f.u. per ml. Each biological sample was performed in triplicate in a plaque assay.

### ELISA

The humoral immune response to p24 was evaluated by an HIV p24 ELISA kit according to the manufacturer’s protocol (Abnove). To detect the humoral immune response to SpCas9 in three patients, recombinant SpCas9 protein (Novoprotein), human albumin (Sigma), or tetanus toxoid (Astarte Biologics) were used to coat the ELISA plates, respectively. The plates were washed five times using ELISA wash buffer after incubation at 4 °C overnight. 1 % BSA blocking solution was used to block plates at room temperature for 1 h, and then washed five times. 50-fold diluted serum samples were added to each well and incubated for 2 hours. Plates were washed three times before adding HRP-conjugated goat anti-human antibody (Sangon) which was diluted 1000-fold in blocking buffer. Plates incubated for 1 h at room temperature and then washed 5 times with washing buffer. TMB Single-Component Substrate solution (Solarbio) was added to the plate and develop for 15 min. The reaction was stopped by ELISA Stop Solution (Solarbio). The OD value of each sample was analyzed by a microplate reader.

### HSV-1 Infection Diagnosis

HSV-1 DNA in tear swabs, corneal buttons, and aqueous humor was amplified using HSV-1 DNA Diagnostic Kit (Triplex International Bioscience) according to the manufacturer’s instructions. Quantitative PCR was performed using a LightCycler 96 real-time PCR system (Roche).

### Immunofluorescence Microscopy

An immunofluorescence protocol was adapted from a previous study *(33)*. Briefly, the corneal buttons were fixed in 4% paraformaldehyde (PFA) overnight at 4 °C and then transferred to 30% sucrose. The optimal cutting temperature compound-embedded tissues were sectioned to 10 μm thickness using a freezing microtome (Leica) and processed for immunofluorescence. Slides were blocked in blocking buffer and incubated with primary antibody against HSV-1 VP5 (Santa Cruz Biotechnology, sc56989) overnight at 4 °C. After washing five times, the slides were incubated with Alexa Fluor 488-conjugated secondary antibody (Jackson ImmunoResearch, 711-545-152). Fluorescent images were collected using a laser scanning confocal microscope (Nikon).

### Immunohistochemistry

Corneal button fixed in 4% formaldehyde and embedded in paraffin. The sections were then blocked with 1% BSA at room temperature for 30 min and incubated with anti-HSV-1 VP5 (Santa Cruz Biotechnology, sc56989), anti-CD4 (Servicebio, gb13064), anti-CD8 (Servicebio, gb11068), anti-CDllb antibody (Servicebio, gb11058) or anti-F4/80 antibody (Servicebio, gb11027) at 4 °C overnight. The sections were then incubated with an anti-rabbit secondary antibody (Servicebio, gb23303) or anti-mouse secondary antibody (Servicebio, gb23301), followed by incubation with freshly prepared DAB substrate solution to detect the antibody. Sections were counterstained with hematoxylin, blued with ammonia water, and then dehydrated and coverslipped.

### CRISPR Off-Target Analysis

The Off-Target Analysis was designed according to the original GUIDE-seq protocol (*40*). Briefly, 293T cells (5×105) were electric transfected with 2 μl double-stranded oligodeoxynucleotide (100 μM), 1.5 μg SpCas9-expressing plasmid and 0.75 μg of UL29-UL8 gRNA-expressing plasmid or an empty plasmid. Genomic DNA was isolated 72 hours after transfection to generate a DNA sequencing library. The remaining steps for sequencing were completed following the GUIDE-seq method described in the study of Montagna et al (*47*). The potential off-target sites identified by GUIDE-seq were PCR amplified and pooled for double-end sequencing using an Illumina nova-seq6000 (Genefund Biotech). Raw data of deep sequencing were analyzed by Cas-analyzer.

### Human corneal HSV-1 infection

Human corneas were supplied by the Eye Bank of the Eye, Ear, Nose and Throat Hospital, Fudan University, under the approval of the hospital ethics committee (EENTIRB-2017-06-07-01). Experiments were conducted according to the Declaration of Helsinki and in compliance with Chinese law. Corneas were evenly divided into twelve halves, and four halves were dosed with 1.5 μg p24 HELP or the same volume PBS by intrastromal injection while the rest four halves were incubated with 20 μM ACV. Corneas were infected with 2 × 10^6^ p.f.u. of HSV-1 17syn+ at 24 hours post treatment. The media were refreshed at two hours post infection and maintained with MEM (containing 10% FBS and 1% P/S). The culture supernatant and cornea were harvested on day 3 to perform the plaque assay, immunohistochemical analysis and RNA/DNA isolation (TaKaRa MiniBEST viral RNA/DNA extraction kit). The UL8 and UL29 on-target sites were PCR amplified and pooled for double-end sequencing using an Illumina nova-seq6000 (Genefund Biotech). Raw data from deep sequencing were analyzed by Cas-analyzer online (version 2016/12/14).

### Best corrected visual acuity (BCVA)

BCVA was assessed at day -1 (pre-injection), week 1, month 1 (M1), M2, M3, M6 and M12 using a standard visual acuity chart via logMAR method. It was scored as the number of correctly read opening directions of letter ‘E’ after adjusting for distance (4 m or 1 m) and described as logMAR to measure the range of acuities from 20/10 to 20/800 (or from −0.30 to +1.6 logMAR). For patients unable to correctly distinguish the opening direction of letter ‘E’ at 1 m, the Berkeley Rudimentary Vision Test battery was performed (*48*) at distances of 1 m and 0.25 m to measure the range of acuities from +1.4 to +2.9 log10 MAR. ‘Light perception’ was assigned +4.0 log10 MAR, ‘hand motion’ and ‘counting fingers’ were assigned +3.0 logMAR and +2.0 logMAR, respectively (*49, 50*).

### Time-domain optical coherence tomography (OCT)

An OCT system (Stratus OCT, Carl Zeiss Meditec, Dublin, CA) developed for ophthalmology research was used to visualize the anterior segment (cornea, anterior chamber, iris, etc.) of participants in this trial at day -1 (pre-injection), week 1, M6 and M12. In this clinical trial, time-domain OCT was mainly used for confirmation of corneal perforation, assessment for synechia of iris and postoperative status of the anterior segment using a 12-mm high-resolution telecentric lens designed for imaging the anterior segment of the eye.

### Electroretinogram (ERG)

A full-field cone and rod ERG was performed via Espion (E2, E3) systems (Diagnosys LLC, Lowell, MA) twelve months post-injection. As described earlier (*51*), ERG recordings were obtained in nondilated eyes using DTL electrodes secured deep in the conjunctival sac. Ground and reference electrodes were taped to the forehead and external canthi of participants, respectively. Flashes and background light were induced by a Ganzfeld color dome to achieve full-field stimulation. By convention, the amplitude and latency of both the a-wave and b-wave generate a total of 4 parameters to define an ERG waveform.

### In vivo corneal confocal microscopy (IVCM)

Laser IVCM (Heidelberg Retina Tomograph 3 with the Rostock Cornea Module, Heidelberg Engineering GmbH, Germany) was routinely conducted on participants twelve months post-injection. As described (*52*), IVCM images were recorded at 30 frames/s. Adjacent images were separated by 1 μm with a lateral resolution of 1 μm/pixel. Performed eyes were topically anesthetized and administered with a layer of gel before reaching the equipment. Fifty to a hundred images of the corneal subbasal layer were obtained and the most representative images were selected by the same masked observer.

### Spectral-domain optical coherence tomography (OCT)

Spectral-domain OCT (Cirrus OCT 5000, Zeiss) was used for the cross-sectional imaging of the retina which were aligned by straightening the major retinal pigment epithelium reflection (*53, 54*). Participants were measured for full retinal thickness, outer nuclear layer thickness, photoreceptor outer segment layer thickness and presence of macular edema on the sixth month post-injection. Quantitative analyses of the photoreceptor cilial anatomy was performed via longitudinal reflectivity profiles. En face imaging with near-infrared illumination was conducted with autofluorescence mode or reflectance mode via a confocal scanning laser ophthalmoscope (*53, 54*).

### Statistics

Data were analysed using GraphPad Prism 8 and presented as mean ± SEM in all experiments (n=3). Unpaired two-tailed Student’s t-tests and one-way ANOVA with Dunnett’s multiple comparisons test were performed to determine the P values (95% confidence interval).

## Supporting information

Protocol

CONSORT extension for Pilot and Feasibility Trials Flow Diagram

## Data Availability

The authors declare that all data supporting the findings of this study are available within the paper and its extended data and supplementary information files. The deep-sequencing data for 293T and patient samples are available at NCBI BioProject. The BioProject IDs are PRJNA823566 and PRJNA823861, respectively. The GUIDE-seq preliminary data is shown on NCBI BioProject with an ID of PRJNA857843. No custom code or mathematical algorithm is used for this study.

## Funding

National Natural Science Foundation of China 81970766 (J.H.)

National Natural Science Foundation of China 82171102 (J.H.)

National Natural Science Foundation of China 31971364 (Y.C.)

Program for Professor of Special Appointment (Eastern Scholar) at Shanghai Institutions of Higher Learning (J.H.)

Shanghai Innovation Development Program 2020-RGZN-02033 (J.H.)

Shanghai Key Clinical Research Program SHDC2020CR3052B (J.H.)

Key forward-looking layout fund from Shanghai Jiao Tong University AF4150049 (Y.C.)

National Key R&D Program of China 2022YFC3400200 (Y.C.)

## Author contributions

Conceptualization: Y.C., J.H.

Methodology: Y.C., J.H., X.J., S.P.

Investigation: A.W., D.Y., Z.Z., S.L., H.L., L.T.

Visualization: A.W., S.L., H.L., L.T.

Funding acquisition: Y.C., J.H.

Project administration: A.W., D.Y., Z.Z.

Supervision: Y.C., J.H.

Writing – original draft: D.Y., Z.Z.

Writing – review & editing: Y.C., J.H., A.W., D.Y., Z.Z.

---

**Extended Fig. 1.**
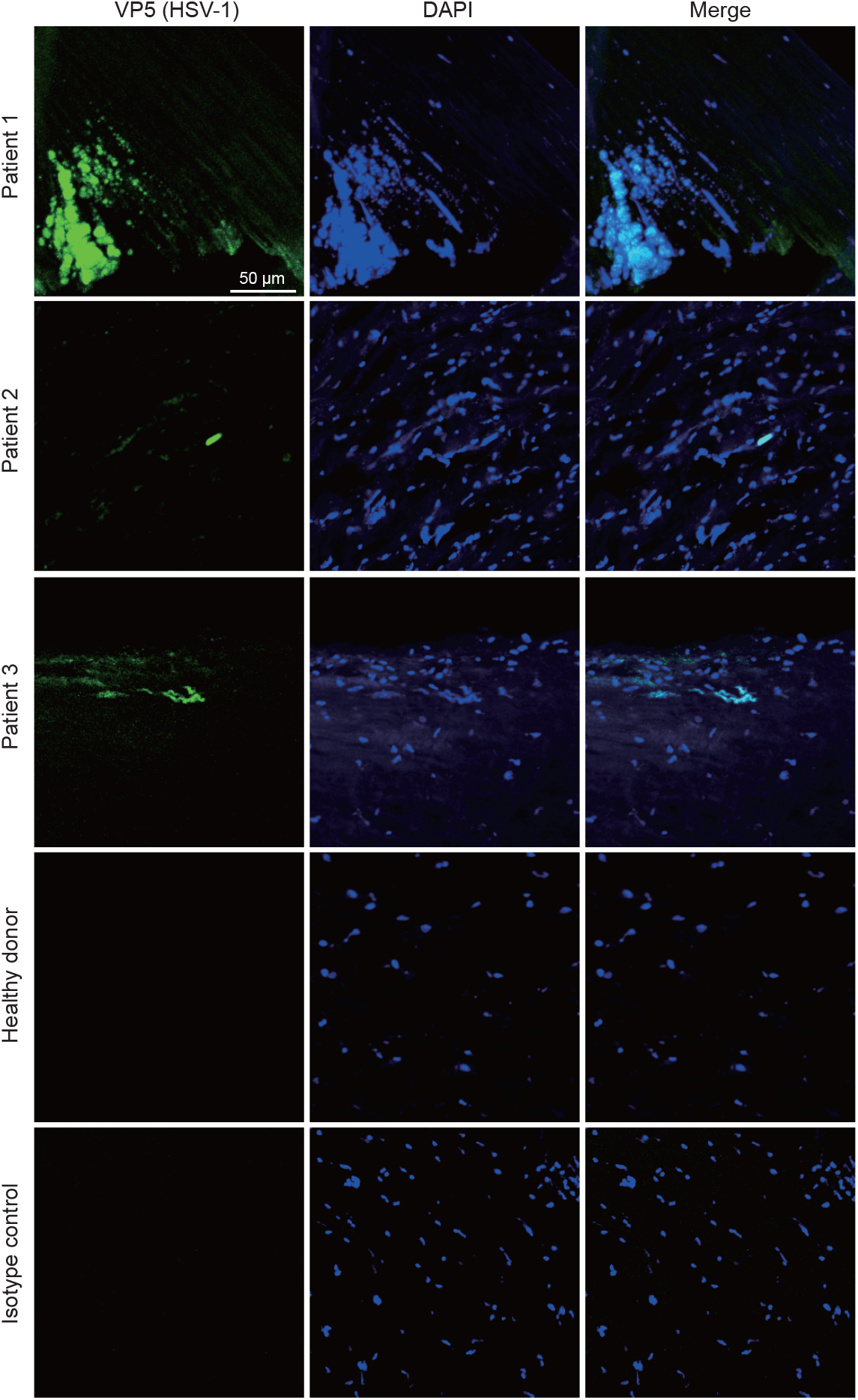
Fluorescence microscopy analysis of HSV-1 in the removed corneal buttons. HSV-1 capsid protein VP5 was presented in green while DAPI was in blue. Their perfect overlap after merging suggests the presence of HSV-1 in the patient corneal tissue before CRISPR treatment.

**Extended Fig. 2.**
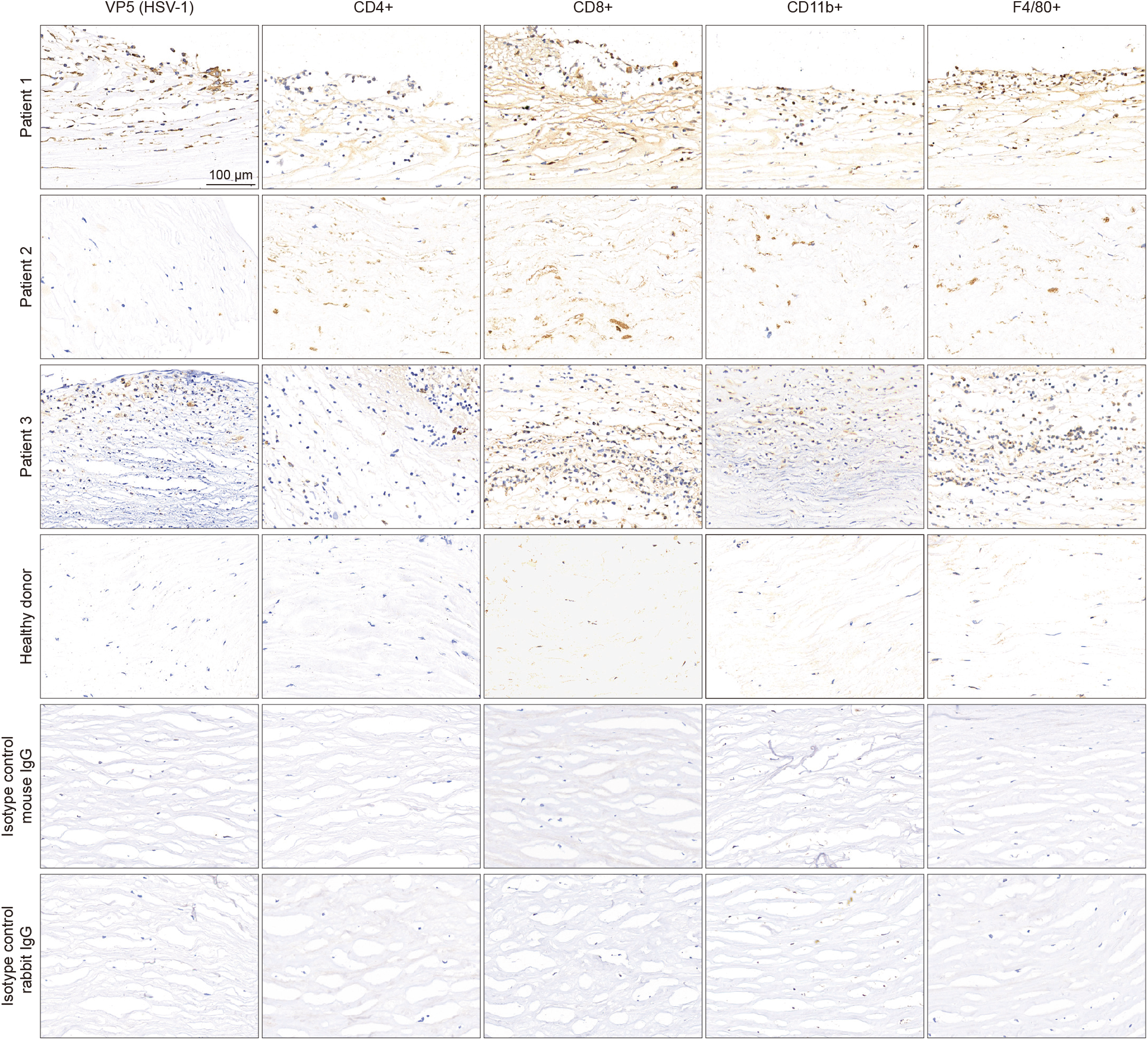
Immunohistochemistry analysis of immune cells in the corneal button. The removed corneal buttons from the patients were stained for T cells (CD4+ and CD8+), myeloid-derived cells (CD11b+) and macrophages (F4/80+). Immunohistochemistry confirmed excess infiltration of inflammatory cells compared to the healthy cornea.

**Extended Fig. 3.**
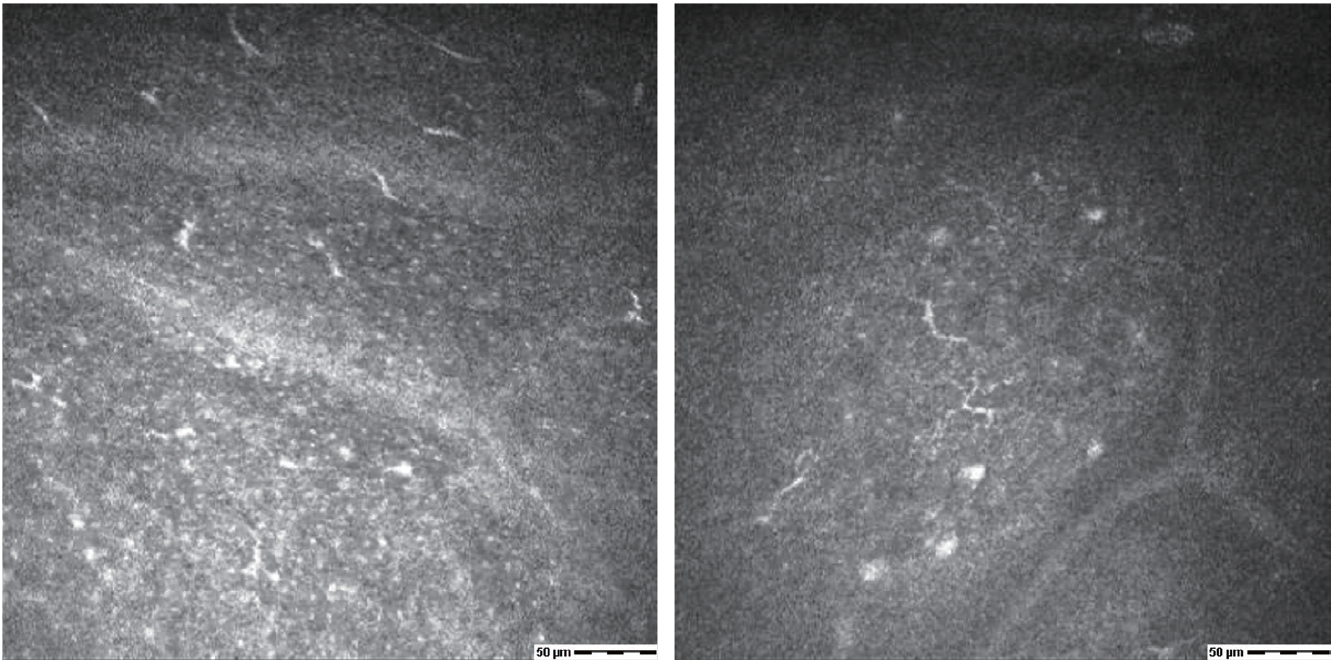
Postoperative IVCM examination of patient 1. IVCM at 12-month post-injection showed very few corneal subbasal nerves in the transplanted corneal graft of patient 1.

**Extended Fig. 4.**
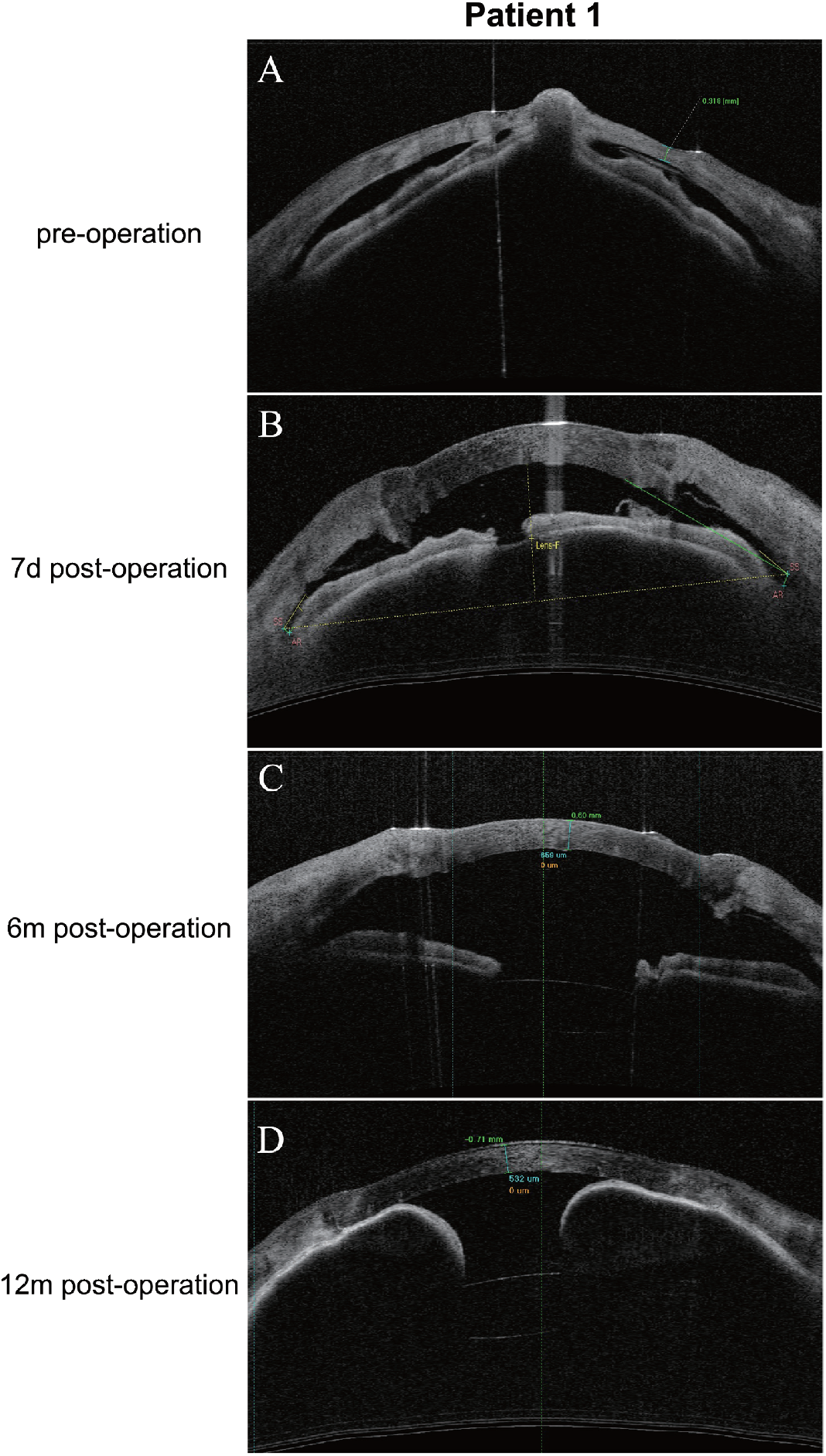
Time-domain OCT of the anterior segment of patient 1. This image shows the time-domain OCT of the anterior segment of patient 1 at different time points (**A-D**). Before the injection, there was iris incarceration in the thinning area of the cornea. OCT showed a slightly shallow anterior chamber after 7 days post-injection and an iris adhesion at the 12-month visit, yet the intraocular pressure of patient 1 was within the normal range. Prophylactic laser therapy was conducted to prevent glaucoma.

**Extended Fig. 5.**
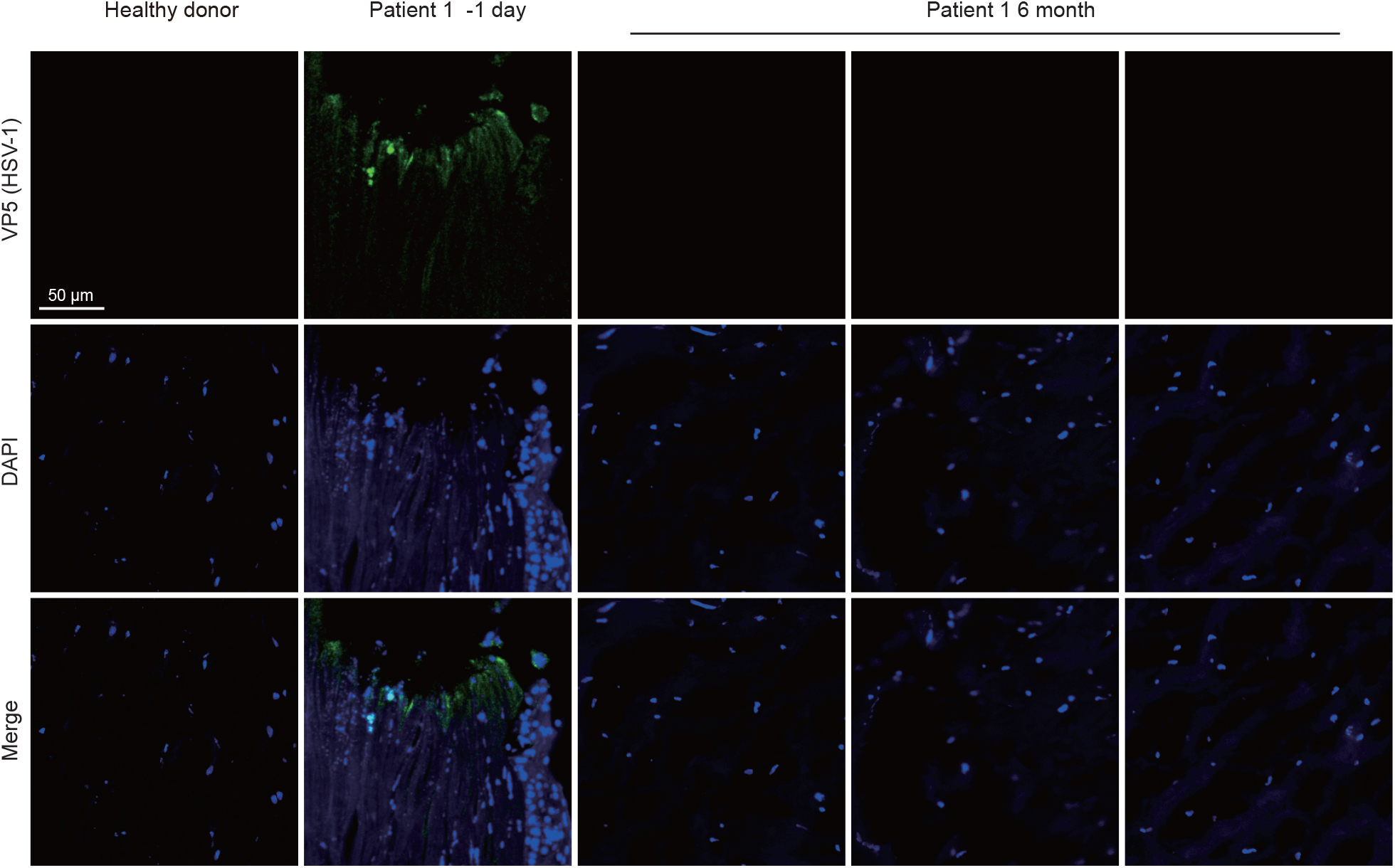
Fluorescence microscopy analysis of HSV-1 in the removed corneal button. After the second corneal transplantation on the right eye of patent 1 at 6 months after PK, the removed corneal button showed no signs of HSV-1 capsid protein VP5 (green) by the fluorescence microscopy. Scale bars, 50 μm.

**Extended Fig. 6.**
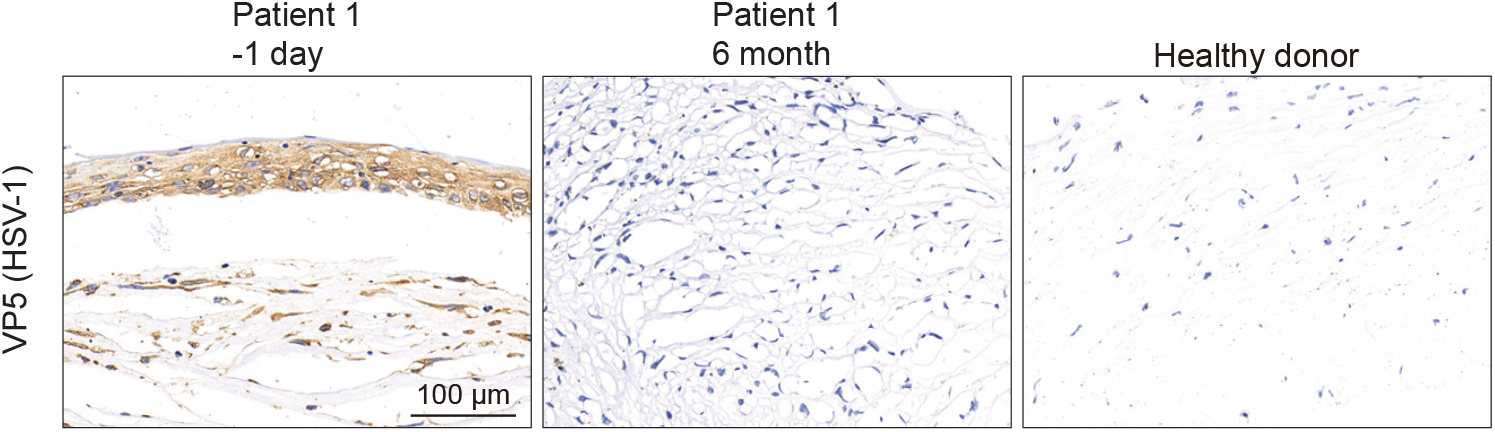
Immunohistochemistry analysis of HSV-1 in the removed corneal button from patient 1. After the second corneal transplantation on his right eye 6 months after PK, the removed corneal button showed no signs of HSV-1 capsid protein VP5 (brown) by immunohistochemistry analysis.

**Extended Fig. 7.**
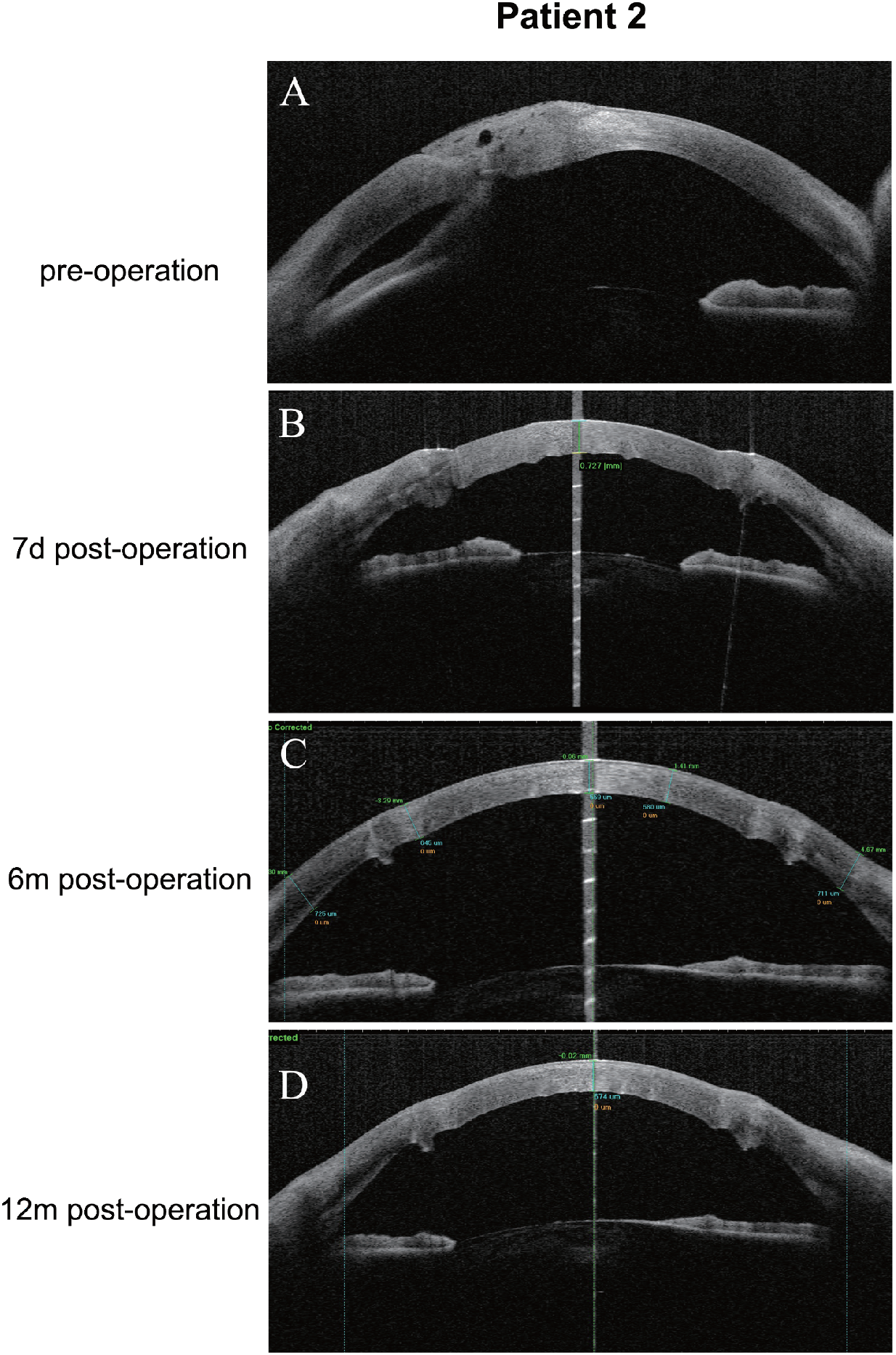
Time-domain OCT of the anterior segment of patient 2. The pre-injection OCT of the anterior segment showed a possible corneal perforation site in patient 2 (**A**). Post-injection OCT images (**B-D**) indicate a deepened anterior chamber, which was maintained for over 12 months after the corneal transplantation.

**Extended Fig. 8.**
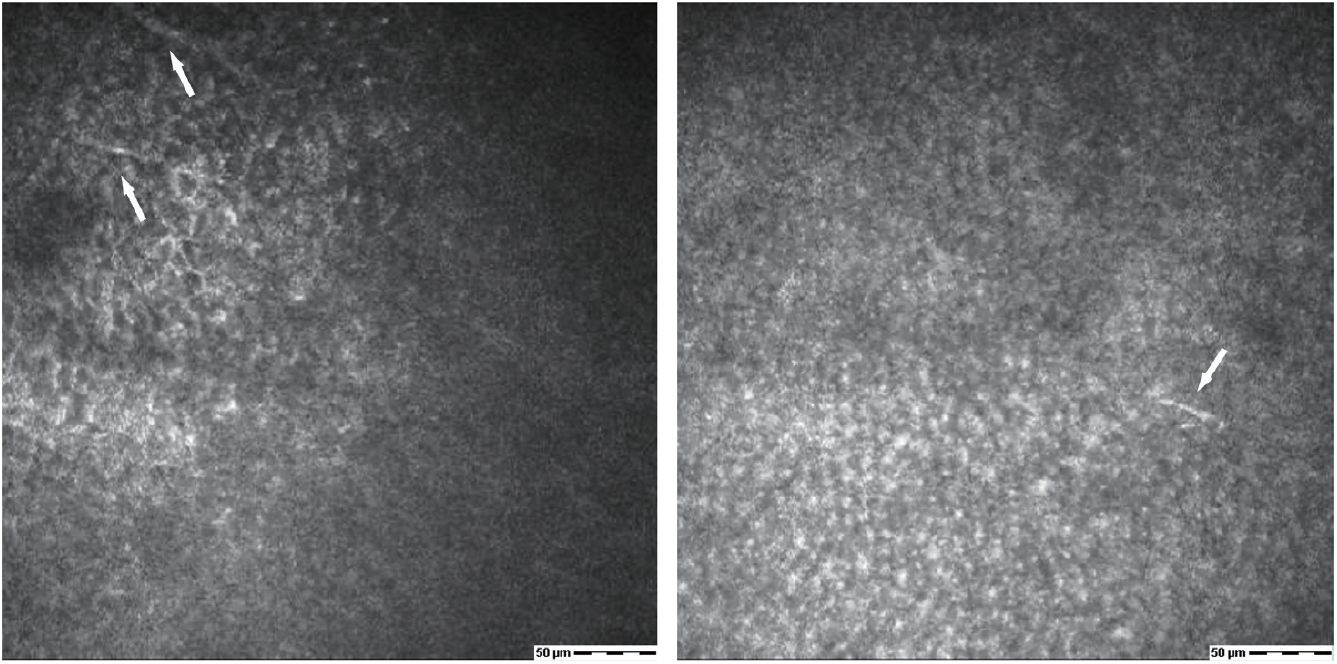
Postoperative IVCM examination of patient 2. IVCM showed slight corneal nerve regeneration (white arrow) in the transplanted corneal graft at the 12 month-follow-up of patient 2.

**Extended Fig. 9.**
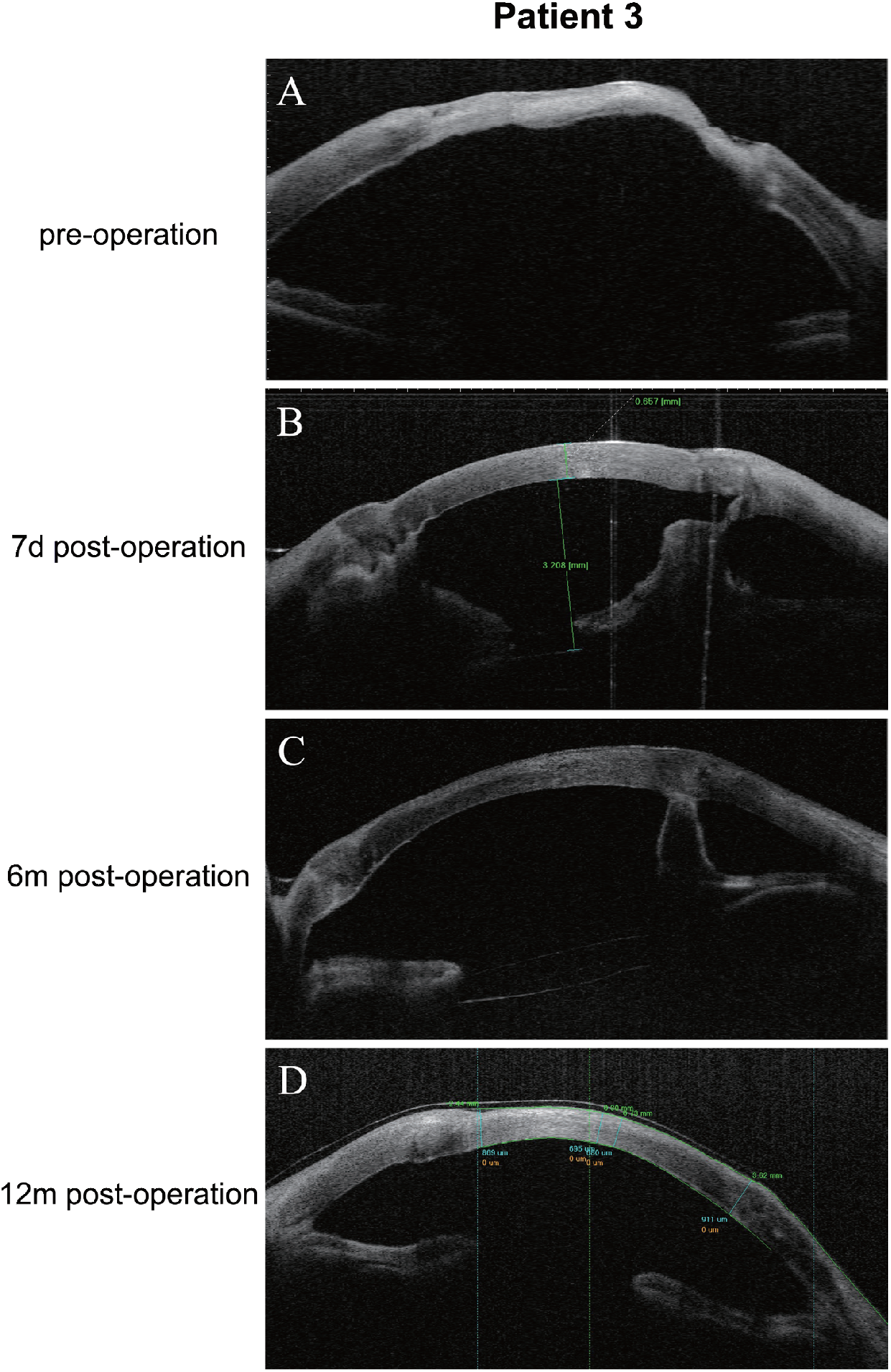
Time-domain OCT of the anterior segment of patient 3. The pre-injection OCT of the anterior segment showed a corneal perforation channel in patient 3 (**A**). Post-injection OCT images (**B-D**) indicate an in-position intraocular lens and a slightly adhered iris to the posterior cornea after PK.

**Extended Fig. 10.**
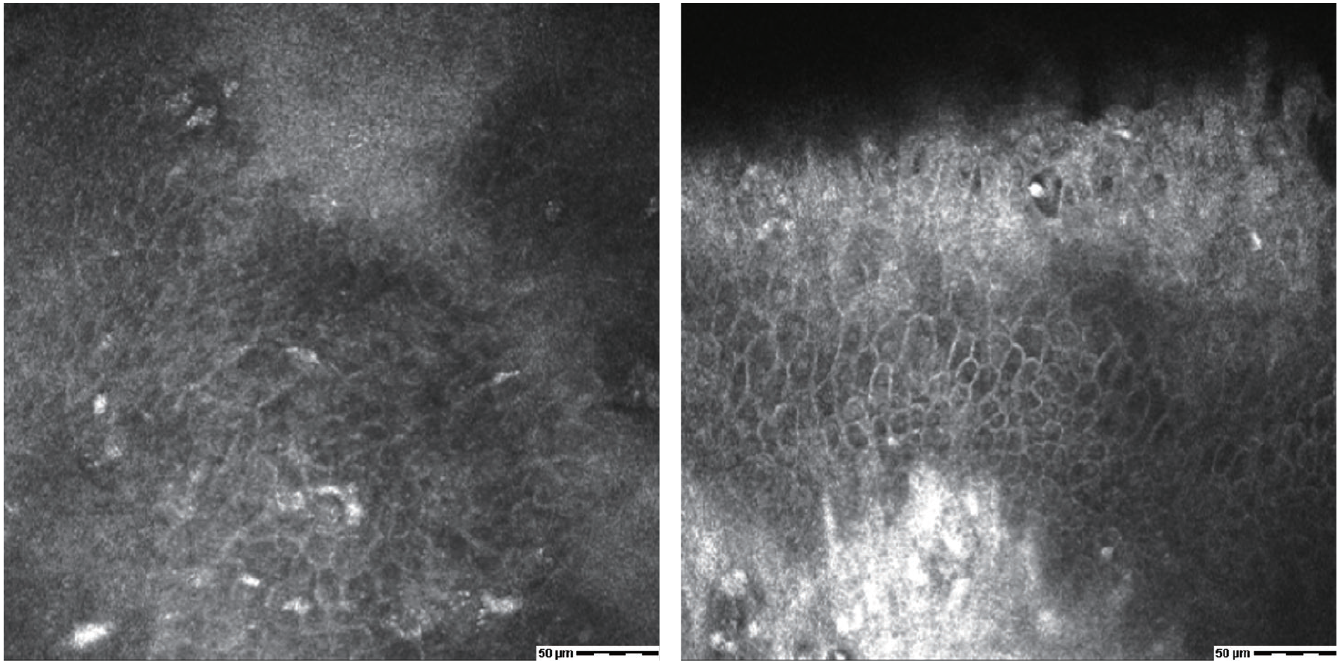
Extended Data Figure10. Postoperative IVCM examination of patient 3. IVCM showed no signs of corneal nerve regeneration at the 12-month follow-up of patient 3. His corneal epithelium density decreased and showed irregular morphologic changes, which indicated the possibility of corneal neurological dystrophy related to HSK.

**Extended Table 1.** P24 antibody test.

| Positive Index* | Positive control | Negative control | -1 day | 1 month | 3 months | 6 months |
| --- | --- | --- | --- | --- | --- | --- |
| **Patient 1** | 15.3 | 0.49 | 0.45 | 0.45 | 0.45 | 0.45 |
| **Patient 2** | 13.4 | 0.51 | 0.50 | 0.44 | 0.46 | 0.44 |
| **Patient 3** | 11.62 | 0.49 | 0.46 | 0.49 | 0.5 | 0.47 |

\* Calculate the mean absorbance values (OD450) for Negative Control using formula:
OD mean (negative control) = (OD (negative control 1) + OD (negative control 2)) / 2.
Calculate the cut-off value: Cut-off = OD mean (negative control) + 0.05
Calculate Positivity Index for each sample:
Positivity Index = mean OD 450 (sample) / Cut-off value.
If Positivity Index value is greater than 1.1, the result is **Positive**.
If Positivity Index value is less than 0.9, the result is **Negative**.

## Supplementary Materials

Other supplementary materials could be found attached to this PDF, including:

Figs. S1 to S20

Tables S1 to S14

Protocol

