## Supplementary Materials for "In Vivo CRISPR Gene Editing in Patients with Herpes Stromal Keratitis"

**Other Supplementary Materials for this manuscript include the following:**

Supplemental Movie

**Table S1. Visual acuity, intraocular pressure and virus tests.**

| | | 1 day pre-injection | 7 days post-injection | 1 month post-injection | 2 months post-injection | 3 months post-injection | 6 months post-injection | 12 months post-injection |
|---|---|---|---|---|---|---|---|---|
| Patient 1 | Best-corrected visual acuity | Light perception | Finger count | Finger count | Finger count | Finger count | 20/100 | 20/100 |
| | Intraocular pressure (mmHg) | Undetectable | 12.0 | 10.4 | 16 | 15.3 | 8 | 21.3 |
| | HSV-1 tests (Ct value) | 21.35 (Cornea)<br>28.37 (Aqueous)<br>30.55*(Tear swab) | -<br>24.09(Tear swab)<br>- | 28.37(Tear swab)<br>31.49(Tear swab)<br>31.08(Tear swab) | -<br>-<br>- | -<br>-<br>- | -<br>-<br>- | -<br>-<br>- |
| Patient 2 | Best-corrected visual acuity | Hand motion | 20/250 | 20/167 | 20/100 | 20/67 | 20/133 | 20/133 |
| | Intraocular pressure (mmHg) | Undetectable | 11.0 | 16.3 | 16.4 | 12.4 | 7.3 | 15.0 |
| | HSV-1 tests (Ct value) | 26.34 (Cornea)<br>36.50 (Aqueous)<br>35.01*(Tear swab) | -<br>-<br>- | -<br>-<br>- | -<br>-<br>- | -<br>-<br>- | -<br>-<br>- | -<br>-<br>- |
| Patient 3 | Best-corrected visual acuity | Hand motion | Hand motion | Hand motion | Hand motion | Hand motion | Hand motion | Finger count |
| | Intraocular pressure (mmHg) | Undetectable | n.a. | 15.0 | 24.1 | 18.5 | 18.1 | 19.3 |
| | HSV-1 tests (Ct value) | 34.9 (Cornea)<br>- (Aqueous)<br>29.5 (Tear swab) | -<br>-<br>- | -<br>-<br>- | -<br>-<br>- | -<br>-<br>- | -<br>-<br>- | -<br>-<br>- |

*Mean HSV-1 titer (Ct value) of three simultaneously sampled tear swabs; n.a., not available; -, Ct value undetectable.

**Table S2. Visual acuity, intraocular pressure and virus tests.**

| Patient | Adverse Event | Start Date | End Date | Placebo Related | Severity | Outcome | Treatment |
|---|---|---|---|---|---|---|---|
| 1 | corneal edema | 11/2020 | 11/2020 | possibly related | mild | resolved | concomitant medication |
| | conjunctival injection | 11/2020 | 11/2020 | unlikely to be related | mild | resolved | concomitant medication |
| | hyphema | 11/2020 | 11/2020 | unrelated | mild | resolved | none |
| | neurotrophic keratitis | 5/2021 | / | unrelated | mild | unresolved | concomitant medication |
| | uncontrolled corneal ulcer | 9/2021 | 11/2021 | unrelated | moderate | resolved | penetrating keratoplasty |
| | cataract | n.a. | 11/2021 | unrelated | mild | resolved | cataract surgery |
| | secondary glaucoma | 10/2021 | 12/2021 | unrelated | mild | resolved | concomitant medication |
| 2 | corneal edema | 1/2021 | 1/2021 | possibly related | mild | resolved | concomitant medication |
| | conjunctival injection | 1/2021 | 1/2021 | unlikely to be related | mild | resolved | concomitant medication |
| | itching | 2/2021 | 3/2021 | unrelated | mild | resolved | concomitant medication |
| | endophthalmitis | 7/2021 | 7/2021 | unrelated | moderate | resolved | vitrectomy and antibiotic injection |
| | cataract | n.a. | / | unrelated | mild | unresolved | none |
| 3 | corneal edema | 5/2021 | 5/2021 | possibly related | mild | resolved | concomitant medication |
| | conjunctival injection | 5/2021 | 5/2021 | unlikely to be related | mild | resolved | concomitant medication |
| | graft rejection | 8/2021 | 10/2021 | unrelated | mild | resolved | concomitant medication |
| | secondary glaucoma | 7/2021 | 8/2021 | unrelated | mild | resolved | concomitant medication |
| | cataract | n.a. | / | unrelated | mild | unresolved | none |

**A**

```
                                       20         40         60
                                       |          |          |
HSV-1 KOS UL23  MASYPCHQHA SAFDQAARSR GHSNRRTALR PRRQQEATEV RLEQKMPTLL RVYIDGPHGM 60
HSV-1 17+ UL23  MASYPCHQHA SAFDQAARSR GHNNRRTALR PRRQQKATEV RLEQKMPTLL RVYIDGPHGM 60
Patient 1 UL23  ---YPCHQHA SAFDQAARSR GHSNRRTALR PRRQQEATEV RLEQKMPTLL RVYIDGPHGM 57
Patient 2 UL23  ---YPCHQHA SAFDQAARSR GHSNRRTALR PRRQQEATEV RLEQKMPTLL RVYIDGPHGM 57
Patient 3 UL23  ---YPCHQHA SAFDQAARSR GHSNRRTALR PRRQQEATEV RLEQKMPTLL RVYIDGPHGM 57

                                       80         100        120
                                       |          |          |
HSV-1 KOS UL23  GKTTTTQLLV ALGSRDDIVY VPEPMTYWQV LGASETIANI YTTQHRLDQG EISAGDAAVV 120
HSV-1 17+ UL23  GKTTTTQLLV ALGSRDDIVY VPEPMTYWRV LGASETIANI YTTQHRLDQG EISAGDAAVV 120
Patient 1 UL23  GKTTTTQLLV ALGSRDDIVY VPEPMTYWQV LGASETIANI YTTQHRLDQG EISAGDAAVV 117
Patient 2 UL23  GKTTTTQLLV ALGSRDDIVY VPEPMTYWQV LGASETIANI YTTQHRLDQG EISAGDAAVV 117
Patient 3 UL23  GKTTTTQLLV ALGSRDDIVY VPEPMTYWQV LGASETIANI YTTQHRLDQG EISAGDAAVV 117

                                       140        160        180
                                       |          |          |
HSV-1 KOS UL23  MTSAQITMGM PYAVTDAVLA PHIGGEAGSS HAPPPALTLI FDRHPIAALL CYPAARYLMG 180
HSV-1 17+ UL23  MTSAQITMGM PYAVTDAVLA PHIGGEAGSS HAPPPALTLI FDRHPIAALL CYPAARYLMG 180
Patient 1 UL23  MTSAQITMGM PYAVTDAVLA PHIGGEAGSS HAPPPALTLI FDRHPIAALL CYPAARYLMG 177
Patient 2 UL23  MTSAQITMGM PYAVTDAVLA PHIGGEAGSS HAPPPALTLI FDRHPIAALL CYPAARYLMG 177
Patient 3 UL23  MTSAQITMGM PYAVTDAVLA PHIGGEAGSS HAPPPALTLI FDRHPIAALL CYPAARYLMG 177

                                       200        220        240
                                       |          |          |
HSV-1 KOS UL23  SMTPQAVLAF VALIPPTLPG TNIVLGALPE DRHIDRLAKR QRPGERLDLA MLAAIRRVYG 240
HSV-1 17+ UL23  SMTPQAVLAF VALIPPTLPG TNIVLGALPE DRHIDRLAKR QRPGERLDLA MLAAIRRVYG 240
Patient 1 UL23  SMTPQAVLAF VALIPPTLPG TNIVLGALPE DRHIDRLAKR QRPGERLDLA MLAAIRRVYG 237
Patient 2 UL23  SMTPQAVLAF VALIPPTLPG TNIVLGALPE DRHIDRLAKR QRPGERLDLA MLAAIRRVYG 237
Patient 3 UL23  SMTPQAVLAF VALIPPTLPG TNIVLGALPE DRHIDRLAKR QRPGERLDLA MLAAIRRVYG 237

                                       260        280        300
                                       |          |          |
HSV-1 KOS UL23  LLANTVRYLQ GGGSWREDWG QLSGTAVPPQ GAEPQSNAGP RPHIGDTLFT LFRAPELLAP 300
HSV-1 17+ UL23  LLANTVRYLQ GGGSWREDWG QLSGAAVPPQ GAEPQSNAGP RPHIGDTLFT LFRAPELLAP 300
Patient 1 UL23  LLANTVRYLQ GGGSWREDWG QLSGTAVPPQ GAEPQSNAGP RPHIGDTLFT LFRAPELLAP 297
Patient 2 UL23  LLANTVRYLQ GGGSWREDWG QLSGTAVPPQ GAEPQSNAGP RPHIGDTLFT LFRAPELLAP 297
Patient 3 UL23  LLANTVRYLQ GGGSWREDWG QLSGTAVPPQ GAEPQSNAGP RPHIGDTLFT LFRAPELLAP 297

                                       320        340        360
                                       |          |          |
HSV-1 KOS UL23  NGDLYNVFAW ALDVLAKRLR PMHVFILDYD QSPAGCRDAL LQLTSGMVQT HVTTPGSIPT 360
HSV-1 17+ UL23  NGDLYNVFAW ALDVLAKRLR PMHVFILDYD QSPAGCRDAL LQLTSGMVQT HVTTPGSIPT 360
Patient 1 UL23  NGDLYNVFAW ALDVLAKRLR PMHVFILDYD QSPAGCRDAL LQLTSGMVQT HVTTPGSIPT 357
Patient 2 UL23  NGDLYNVFAW ALDVLAKRLR PMHVFILDYD QSPAGCRDAL LQLTSGMVQT HVTTPGSIPT 357
Patient 3 UL23  NGDLYNVFAW ALDVLAKRLR PMHVFILDYD QSPAGCRDAL LQLTSGMVQT HVTTPGSIPT 357

HSV-1 KOS UL23  ICDLARTFAR EMGEAN 377
HSV-1 17+ UL23  ICDLARTFAR EMGEAN 377
Patient 1 UL23  ICDLARTFAR EMGEAN 374
Patient 2 UL23  ICDLARTFAR EMGEAN 374
Patient 3 UL23  ICDLARTFAR EMGEAN 374
```

**B**

```
                                        20                            40                            60
HSV-1 KOS UL30  MFSGGGGPLS PGGKSAARAA SGFFAPAGPR GAGRGPPPCL RQNFYNPYLA PVGTQQKPTG 60
HSV-1 17+ UL30  MFSGGGGPLS PGGKSAARAA SGFFAPAGPR GASRGPPPCL RQNFYNPYLA PVGTQQKPTG 60
Patient 1 UL30  ---------- -------RAA SGFFAPAGPR GASRGPPPCL RQNFYNPYLA PVGTQQKPTG 43
Patient 2 UL30  ---------- -------RAA SGFFAPAGPR GAGRGPPPCL RQNFYNPYLA PVGTQQKPTG 43
Patient 3 UL30  ---------- -------RAA SGFFAPAGPR GAGRGPPPCL RQNFYNPYLA PVGTQQKPTG 43
                                        80                           100                           120
HSV-1 KOS UL30  PTQRHTYYSE CDEFRFIAPR VLDEDAPPEK RAGVHDGHLK RAPKVYCGGD ERDVLRVGSG 120
HSV-1 17+ UL30  PTQRHTYYSE CDEFRFIAPR VLDEDAPPEK RAGVHDGHLK RAPKVYCGGD ERDVLRVGSG 120
Patient 1 UL30  PTQRHTYYSE CDEFRFIAPR VLDEDAPPEK RAGVHDGHLK RAPKVYCGGD ERDVLRVGSG 103
Patient 2 UL30  PTQRHTYYSE CDEFRFIAPR VLDEDAPPEK RAGVHDGHLK RAPKVYCGGD ERDVLRVGSG 103
Patient 3 UL30  PTQRHTYYSE CDEFRFIAPR VLDEDAPPEK RAGVHDGHLK RAPKVYCGGD ERDVLRVGSG 103
                                       140                           160                           180
HSV-1 KOS UL30  GFWPRRSRLW GGVDHAPAGF NPTVTVFHVY DILENVEHAY GMRAAQFHAR FMDAITPTGT 180
HSV-1 17+ UL30  GFWPRRSRLW GGVDHAPAGF NPTVTVFHVY DILENVEHAY GMRAAQFHAR FMDAITPTGT 180
Patient 1 UL30  GFWPRRSRLW GGVDHAPAGF NPTVTVFHVY DILENVEHAY GMRAAQFHAR FMDAITPTGT 163
Patient 2 UL30  GFWPRRSRLW GGVDHAPAGF NPTVTVFHVY DILENVEHAY GMRAAQFHAR FMDAITPTGT 163
Patient 3 UL30  GFWPRRSRLW GGVDHAPAGF NPTVTVFHVY DILENVEHAY GMRAAQFHAR FMDAITPTGT 163
                                       200                           220                           240
HSV-1 KOS UL30  VITLLGLTPE GHRVAVHVYG TRQYFYMNKE EVDRHLQCRA PRDLCERMAA ALRESPGASF 240
HSV-1 17+ UL30  VITLLGLTPE GHRVAVHVYG TRQYFYMNKE EVDRHLQCRA PRDLCERMAA ALRESPGASF 240
Patient 1 UL30  VITLLGLTPE GHRVAVHVYG TRQYFYMNKE EVDRHLQCRA PRDLCERMAA ALRESPGASF 223
Patient 2 UL30  VITLLGLTPE GHRVAVHVYG TRQYFYMNKE EVDRHLQCRA PRDLCERMAA ALRESPGASF 223
Patient 3 UL30  VITLLGLTPE GHRVAVHVYG TRQYFYMNKE EVDRHLQCRA PRDLCERMAA ALRESPGASF 223
                                       260                           280                           300
HSV-1 KOS UL30  RGISADHFEA EVVERTDVYY YETRPALFYR VYVRSGRVLS YLCDNFCPAI KKYEGGVDAT 300
HSV-1 17+ UL30  RGISADHFEA EVVERTDVYY YETRPALFYR VYVRSGRVLS YLCDNFCPAI KKYEGGVDAT 300
Patient 1 UL30  RGISADHFEA EVVERTDVYY YETRPALFYR VYVRSGRVLS YLCDNFCPAI KKYEGGVDAT 283
Patient 2 UL30  RGISADHFEA EVVERTDVYY YETRPALFYR VYVRSGRVLS YLCDNFCPAI KKYEGGVDAT 283
Patient 3 UL30  RGISADHFEA EVVERTDVYY YETRPALFYR VYVRSGRVLS YLCDNFCPAI KKYEGGVDAT 283
                                       320                           340                           360
HSV-1 KOS UL30  TRFILDNPGF VTFGWYRLKP GRNNTLAQPR APMAFGTSSD VEFNCTADNL AIEGGMSDLP 360
HSV-1 17+ UL30  TRFILDNPGF VTFGWYRLKP GRNNTLAQPR APMAFGTSSD VEFNCTADNL AIEGGMSDLP 360
Patient 1 UL30  TRFILDNPGF VTFGWYRLKP GRNNTLAQPR APMAFGTSSD VEFNCTADNL AIEGGMSDLP 343
Patient 2 UL30  TRFILDNPGF VTFGWYRLKP GRNNTLAQPR APMAFGTSSD VEFNCTADNL AIEGGMSDLP 343
Patient 3 UL30  TRFILDNPGF VTFGWYRLKP GRNNTLAQPR APMAFGTSSD VEFNCTADNL AIEGGMSDLP 343
                                       380                           400                           420
HSV-1 KOS UL30  AYKLMCFDIE CKAGGEDELA FPVAGHPEDL VIQISCLLYD LSTTALEHVL LFSLGSCDLP 420
HSV-1 17+ UL30  AYKLMCFDIE CKAGGEDELA FPVAGHPEDL VIQISCLLYD LSTTALEHVL LFSLGSCDLP 420
Patient 1 UL30  AYKLMCFDIE CKAGGEDELA FPVAGHPEDL VIQISCLLYD LSTTALEHVL LFSLGSCDLP 403
Patient 2 UL30  AYKLMCFDIE CKAGGEDELA FPVAGHPEDL VIQISCLLYD LSTTALEHVL LFSLGSCDLP 403
Patient 3 UL30  AYKLMCFDIE CKAGGEDELA FPVAGHPEDL VIQISCLLYD LSTTALEHVL LFSLGSCDLP 403
                                       440                           460                           480
HSV-1 KOS UL30  ESHLNELAAR GLPTPVVLEF DSEFEMLLAF MTLVKQYGPE FVTGYNIINF DWPFLLAKLT 480
HSV-1 17+ UL30  ESHLNELAAR GLPTPVVLEF DSEFEMLLAF MTLVKQYGPE FVTGYNIINF DWPFLLAKLT 480
Patient 1 UL30  ESHLNELAAR GLPTPVVLEF DSEFEMLLAF MTLVKQYGPE FVTGYNIINF DWPFLLAKLT 463
Patient 2 UL30  ESHLNELAAR GLPTPVVLEF DSEFEMLLAF MTLVKQYGPE FVTGYNIINF DWPFLLAKLT 463
Patient 3 UL30  ESHLNELAAR GLPTPVVLEF DSEFEMLLAF MTLVKQYGPE FVTGYNIINF DWPFLLAKLT 463
                                       500                           520                           540
HSV-1 KOS UL30  DIYKVPLDGY GRMNGRGVFR VWDIGQSHFQ KRSKIKVNGM VNIDMYGIIT DKIKLSSYKL 540
HSV-1 17+ UL30  DIYKVPLDGY GRMNGRGVFR VWDIGQSHFQ KRSKIKVNGM VNIDMYGIIT DKIKLSSYKL 540
Patient 1 UL30  DIYKVPLDGY GRMNGRGVFR VWDIGQSHFQ KRSKIKVNGM VNIDMYGIIT DKIKLSSYKL 523
Patient 2 UL30  DIYKVPLDGY GRMNGRGVFR VWDIGQSHFQ KRSKIKVNGM VNIDMYGIIT DKIKLSSYKL 523
Patient 3 UL30  DIYKVPLDGY GRMNGRGVFR VWDIGQSHFQ KRSKIKVNGM VNIDMYGIIT DKIKLSSYKL 523
                                       560                           580                           600
HSV-1 KOS UL30  NAVAEAVLKD KKKDLSYRDI PAYYATGPAQ RGVIGEYCIQ DSLLVGQLFF KFLPHLELSA 600
HSV-1 17+ UL30  NAVAEAVLKD KKKDLSYRDI PAYYAAGPAQ RGVIGEYCIQ DSLLVGQLFF KFLPHLELSA 600
Patient 1 UL30  NAVAEAVLKD KKKDLSYRDI PAYYAAGPAQ RGVIGEYCIQ DSLLVGQLFF KFLPHLELSA 583
Patient 2 UL30  NAVAEAVLKD KKKDLSYRDI PAYYATGPAQ RGVIGEYCIQ DSLLVGQLFF KFLPHLELSA 583
Patient 3 UL30  NAVAEAVLKD KKKDLSYRDI PAYYATGPAQ RGVIGEYCIQ DSLLVGQLFF KFLPHLELSA 583
                                       620                           640                           660
HSV-1 KOS UL30  VARLAGINIT RTIYDGQQIR VFTCLLRLAD QKGFILPDTQ GRFRGAGGEA PKRPAAARED 660
HSV-1 17+ UL30  VARLAGINIT RTIYDGQQIR VFTCLLRLAD QKGFILPDTQ GRFRGAGGEA PKRPAAARED 660
Patient 1 UL30  VARLAGINIT RTIYDGQQIR VFTCLLRLAD QKGFILPDTQ GRFRGAGGEA PKRPAAARED 643
Patient 2 UL30  VARLAGINIT RTIYDGQQIR VFTCLLRLAD QKGFILPDTQ GRFRGAGGEA PKRPAAARED 643
Patient 3 UL30  VARLAGINIT RTIYDGQQIR VFTCLLRLAD QKGFILPDTQ GRFRGAGGEA PKRPAAARED 643
                                       680                           700                           720
HSV-1 KOS UL30  EERPEEEGED EDEREEGGGE REPEGARETA GRHVGYQGAR VLDPTSGFHV NPVVVFDFAS 720
HSV-1 17+ UL30  EERPEEEGED EDEREEGGGE REPEGARETA GRHVGYQGAR VLDPTSGFHV NPVVVFDFAS 720
Patient 1 UL30  EERPEEEGED EDEREEGGGE REPEGARETA GRHVGYQGAR VLDPTSGFHV NPVVVFDFAS 703
Patient 2 UL30  EERPEEEGED EDEREEGGGE REPEGARETA GRHVGYQGAR VLDPTSGFHV NPVVVFDFAS 703
Patient 3 UL30  EERPEEEGED EDEREEGGGE REPEGARETA GRHVGYQGAR VLDPTSGFHV NPVVVFDFAS 703
```

```
                                       740                            760                            780
HSV-1 KOS UL30  LYPSIIQAHN LCFSTLSLRA DAVAHLEAGK DYLEIEVGGR RLFFVKAHVR ESLLSILLRD 780
HSV-1 17+ UL30  LYPSIIQAHN LCFSTLSLRA DAVAHLEAGK DYLEIEVGGR RLFFVKAHVR ESLLSILLRD 780
Patient 1 UL30  LYPSIIQAHN LCFSTLSLRA DAVAHLEAGK DYLEIEVGGR RLFFVKAHVR ESLLSILLRD 763
Patient 2 UL30  LYPSIIQAHN LCFSTLSLRA DAVAHLEAGK DYLEIEVGGR RLFFVKAHVR ESLLSILLRD 763
Patient 3 UL30  LYPSIIQAHN LCFSTLSLRA DAVAHLEAGK DYLEIEVGGR RLFFVKAHVR ESLLSILLRD 763
                                       800                            820                            840
HSV-1 KOS UL30  WLAMRKQIRS RIPQSSPEEA VLLDKQQAAI KVVCNSVYGF TGVQHGLLPC LHVAATVTTI 840
HSV-1 17+ UL30  WLAMRKQIRS RIPQSSPEEA VLLDKQQAAI KVVCNSVYGF TGVQHGLLPC LHVAATVTTI 840
Patient 1 UL30  WLAMRKQIRS RIPQSSPEEA VLLDKQQAAI KVVCNSVYGF TGVQHGLLPC LHVAATVTTI 823
Patient 2 UL30  WLAMRKQIRS RIPQSSPEEA VLLDKQQAAI KVVCNSVYGF TGVQHGLLPC LHVAATVTTI 823
Patient 3 UL30  WLAMRKQIRS RIPQSSPEEA VLLDKQQAAI KVVCNSVYGF TGVQHGLLPC LHVAATVTTI 823
                                       860                            880                            900
HSV-1 KOS UL30  GREMLLATRE YVHARWAAFE QLLADFPEAA DMRAPGPYSM RIIYGDTDSI FVLCRGLTAA 900
HSV-1 17+ UL30  GREMLLATRE YVHARWAAFE QLLADFPEAA DMRAPGPYSM RIIYGDTDSI FVLCRGLTAA 900
Patient 1 UL30  GREMLLATRE YVHARWAAFE QLLADFPEAA DMRAPGPYSM RIIYGDTDSI FVLCRGLTAA 883
Patient 2 UL30  GREMLLATRE YVHARWAAFE QLLADFPEAA DMRAPGPYSM RIIYGDTDSI FVLCRGLTAA 883
Patient 3 UL30  GREMLLATRE YVHARWAAFE QLLADFPEAA DMRAPGPYSM RIIYGDTDSI FVLCRGLTAA 883
                                       920                            940                            960
HSV-1 KOS UL30  GLTAMGDKMA SHISRALFLP PIKLECEKTF TKLLLIAKKK YIGVIYGGKM LIKGVDLVRK 960
HSV-1 17+ UL30  GLTAVGDKMA SHISRALFLP PIKLECEKTF TKLLLIAKKK YIGVIYGGKM LIKGVDLVRK 960
Patient 1 UL30  GLTAVGDKMA SHISRALFLP PIKLECEKTF TKLLLIAKKK YIGVIYGGKM LIKGVDLVRK 943
Patient 2 UL30  GLTAMGDKMA SHISRALFLP PIKLECEKTF TKLLLIAKKK YIGVIYGGKM LIKGVDLVRK 943
Patient 3 UL30  GLTAMGDKMA SHISRALFLP PIKLECEKTF TKLLLIAKKK YIGVIYGGKM LIKGVDLVRK 943
                                       980                          1,000                          1,020
HSV-1 KOS UL30  NNCAFINRTS RALVDLLFYD DTVSGAAAAL AERPAEEWLA RPLPEGLQAF GAVLVDAHRR 1020
HSV-1 17+ UL30  NNCAFINRTS RALVDLLFYD DTVSGAAAAL AERPAEEWLA RPLPEGLQAF GAVLVDAHRR 1020
Patient 1 UL30  NNCAFINRTS RALVDLLFYD DTVSGAAAAL AERPAEEWLA RPLPEGLQAF GAVLVDAHRR 1003
Patient 2 UL30  NNCAFINRTS RALVDLLFYD DTVSGAAAAL AERPAEEWLA RPLPEGLQAF GAVLVDAHRR 1003
Patient 3 UL30  NNCAFINRTS RALVDLLFYD DTVSGAAAAL AERPAEEWLA RPLPEGLQAF GAVLVDAHRR 1003
                                     1,040                          1,060                          1,080
HSV-1 KOS UL30  ITDPERDIQD FVLTAELSRH PRAYTNKRLA HLTVYYKLMA RRAQVPSIKD RIPYVIVAQT 1080
HSV-1 17+ UL30  ITDPERDIQD FVLTAELSRH PRAYTNKRLA HLTVYYKLMA RRAQVPSIKD RIPYVIVAQT 1080
Patient 1 UL30  ITDPERDIQD FVLTAELSRH PRAYTNKRLA HLTVYYKLMA RRAQVPSIKD RIPYVIVAQT 1063
Patient 2 UL30  ITDPERDIQD FVLTAELSRH PRAYTNKRLA HLTVYYKLMA RRAQVPSIKD RIPYVIVAQT 1063
Patient 3 UL30  ITDPERDIQD FVLTAELSRH PRAYTNKRLA HLTVYYKLMA RRAQVPSIKD RIPYVIVAQT 1063
                                     1,100                          1,120                          1,140
HSV-1 KOS UL30  REVEETVARL AALRELDAAA PGDEPAPPAA LPSPAKRPRE TPSHADPPGG ASKPRKLLVS 1140
HSV-1 17+ UL30  REVEETVARL AALRELDAAA PGDEPAPPAA LPSPAKRPRE TPSPADPPGG ASKPRKLLVS 1140
Patient 1 UL30  REVEETVARL AALRELDAAA PGDEPAPPAA LPSPAKRPRE TPSPADPPGG ASKPRKLLVS 1123
Patient 2 UL30  REVEETVARL AALRELDAAA PGDEPAPPAA LPSPAKRPRE TPSHADPPGG ASKPRKLLVS 1123
Patient 3 UL30  REVEETVARL AALRELDAAA PGDEPAPPAA LPSPAKRPRE TPSHADPPGG ASKPRKLLVS 1123
                                     1,160                          1,180                          1,200
HSV-1 KOS UL30  ELAEDPAYAI AHGVALNTDY YFSHLLGAAC VTFKALFGNN AKITESLLKR FIPEVWHPPD 1200
HSV-1 17+ UL30  ELAEDPAYAI AHGVALNTDY YFSHLLGAAC VTFKALFGNN AKITESLLKR FIPEVWHPPD 1200
Patient 1 UL30  ELAEDPAYAI AHGVALNTDY YFSHL----- ---------- ---------- ---------- 1152
Patient 2 UL30  ELAEDPAYAI AHGVALNTDY YFSHL----- ---------- ---------- ---------- 1152
Patient 3 UL30  ELAEDPAYAI AHGVALNTDY YFSHL----- ---------- ---------- ---------- 1152
                                     1,220
HSV-1 KOS UL30  DVAARLRAAG FGAVGAGATA EETRRMLHRA FDTLA 1235
HSV-1 17+ UL30  DVAARLRTAG FGAVGAGATA EETRRMLHRA FDTLA 1235
Patient 1 UL30  ---------- ---------- ---------- ----- 1152
Patient 2 UL30  ---------- ---------- ---------- ----- 1152
Patient 3 UL30  ---------- ---------- ---------- ----- 1152
```

**Fig. S1. Amino acids alignment for TK and DNA pol.** HSV-1 strains were isolated from the three participants and sequenced for genes encoding thymidine kinase (TK, UL23) and DNA polymerase (DNA pol, UL30). Letter in red color indicated changed amino acids in TK (**A**) and DNA pol (**B**). HSV-1 KOS and 17syn+ strains were used as references. Only amino-acid changes different from both KOS and 17syn+ simultaneously are considered as ACV resistant mutations.

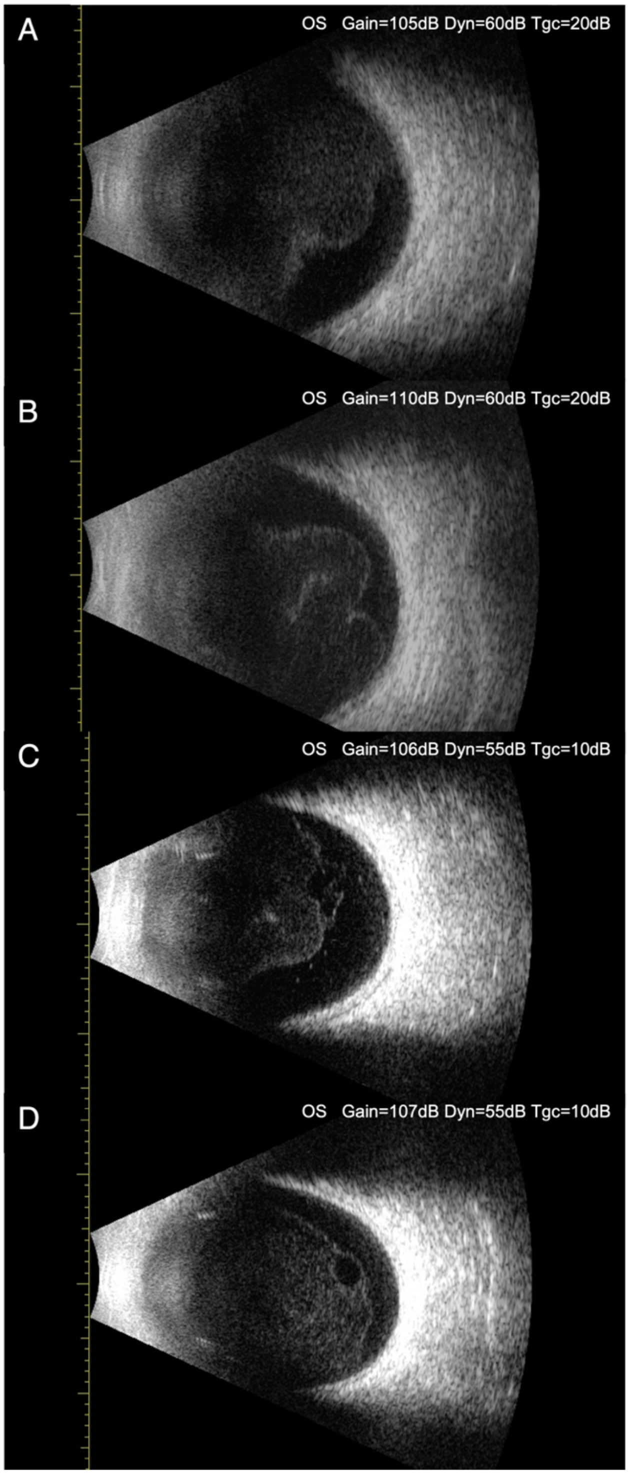


**Fig. S2. Postoperative B-scan ultrasonography of patient 3.** B-scan ultrasonography of the patient 3 on 7 days (**A**), 1 month (**B**), 3 months (**C**) and 6 months (**D**) post-injection. Continuous ribbon-like echo was found in the posterior vitreous body, indicating vitreous opacity with posterior detachment.

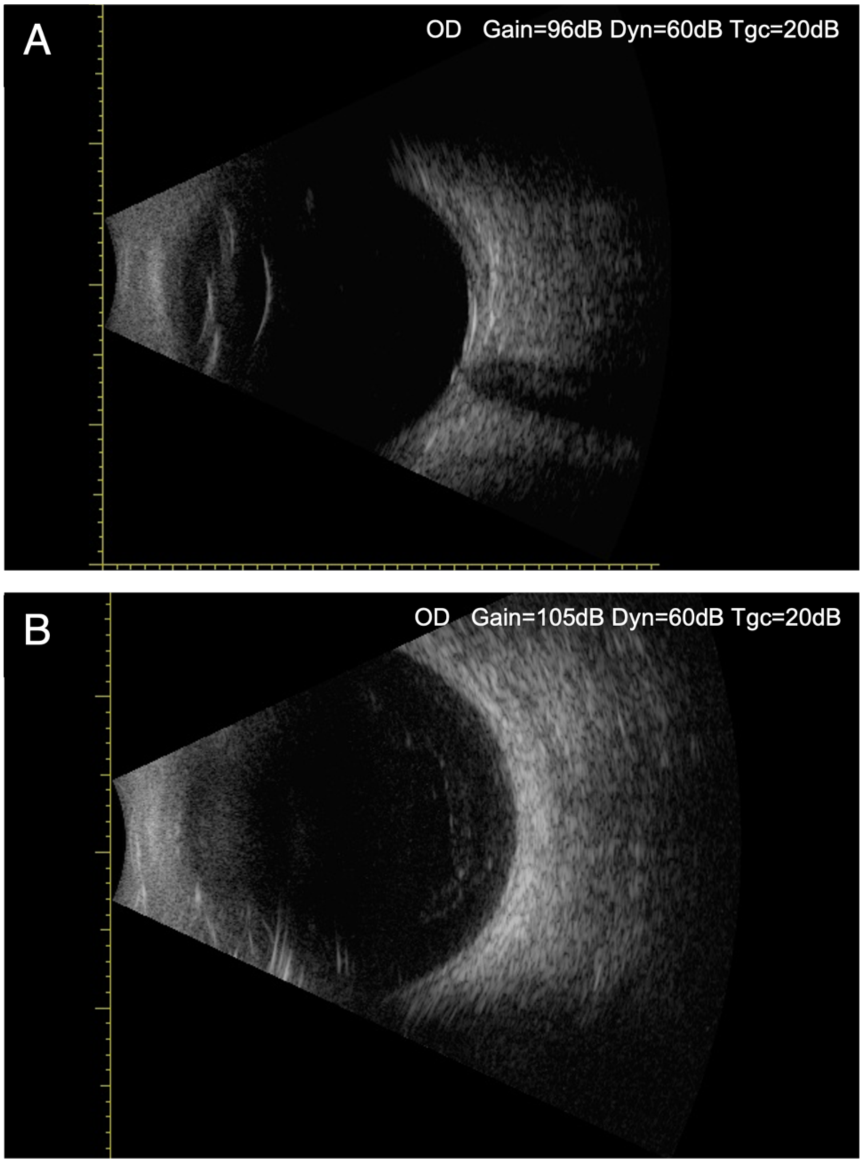


**Fig. S3. Postoperative B-scan ultrasonography of patient 2.** B-scan ultrasonography results of patient 2 on 6 months (**A**) and 9 months (**B**) post-injection showed no remarkable changes in the vitreous body.

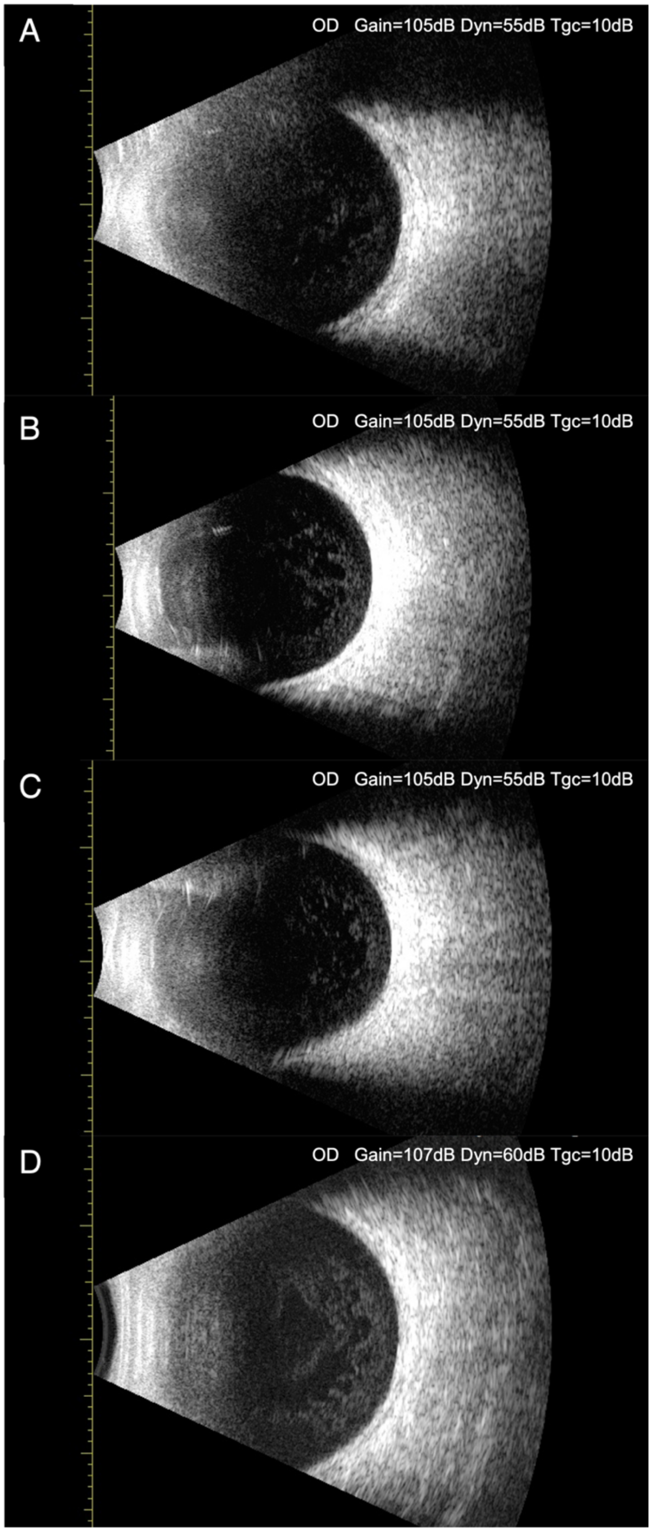


**Fig. S4. Postoperative B-scan ultrasonography of patient 1.** B-scan ultrasonography of patient 1 on 7 days (**A**), 1 month (**B**), 6 months (**C**) and 12 months (**D**) post-injection revealed no remarkable changes in the vitreous body.

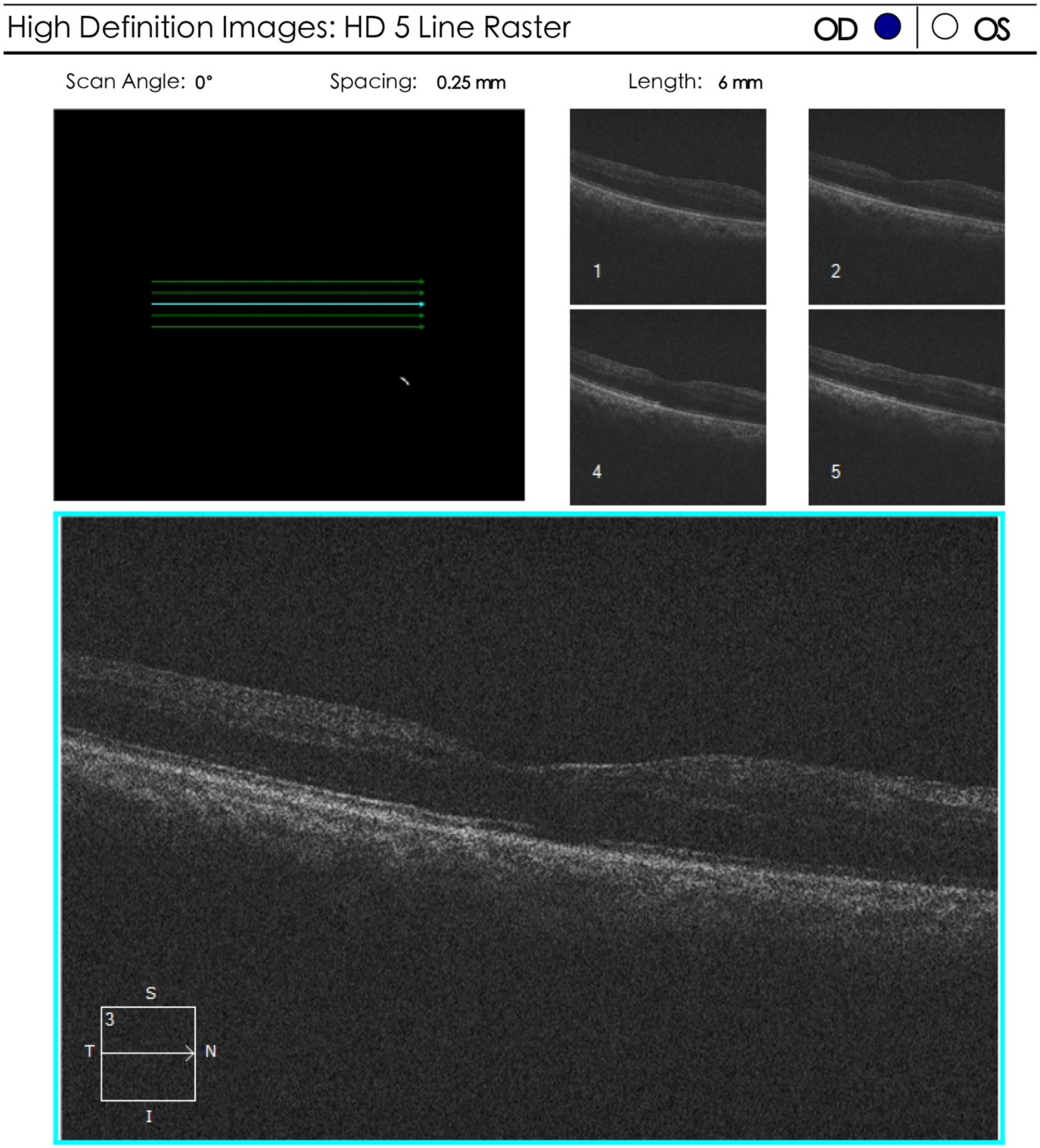


**Fig. S5. Spectral-domain OCT of the retina of patient 2.** The spectral-domain OCT of patient 2’s retina was performed at 6 months post-injection using Cirrus OCT 5000 (Zeiss). The structure of his fundus showed no obvious exception after the administration of HELP.

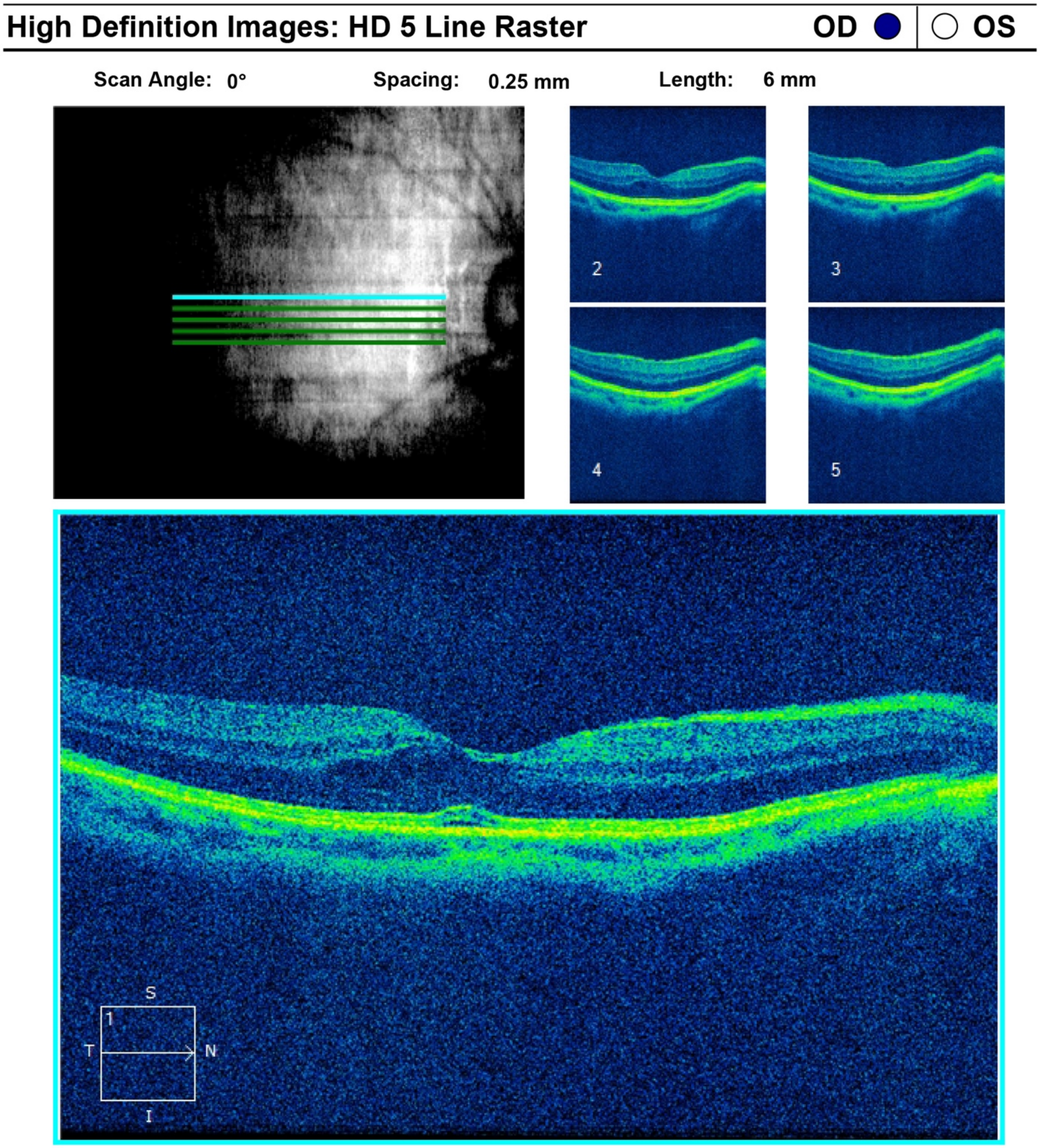


**Fig. S6. Spectral-domain OCT of the retina of patient 1.** The optical coherence tomography revealed mild center-involved intraretinal fluid and subfoveal fluid in the right eye of patient 1 at six months after the treatment. The ellipsoid zone was intact. We continue to follow up on this situation without any intervention.

A

**Diagnosis:**

### 1_Pattern-VEP 2.0&1.0 deg ( Monitor )

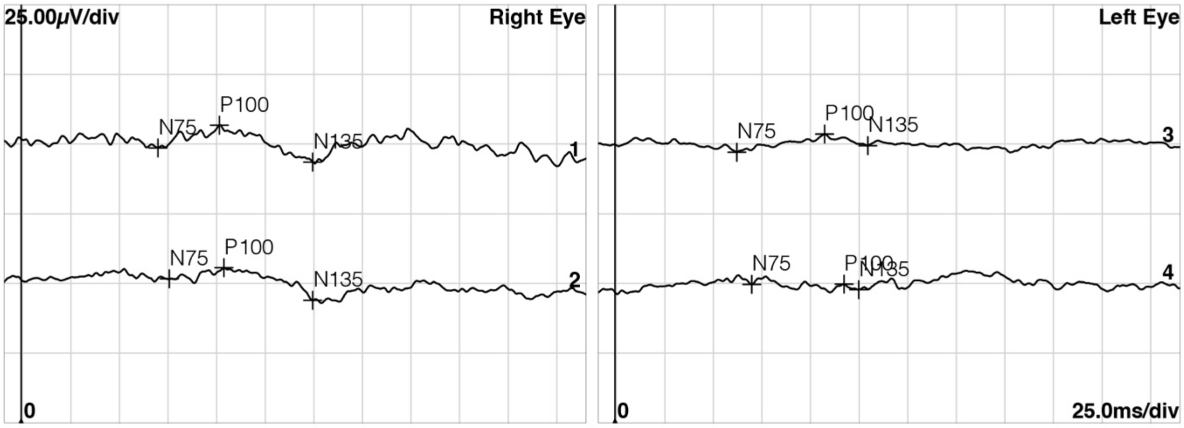


| Normals | - | 96-109 | - | 7.08µV-17.7µV | |
|---|---|---|---|---|---|
| **Channel** | **N75 [ms]** | **P100 [ms]** | **N135 [ms]** | **N75-P100** | **P100-N135** |
| **1 R1 2,0 deg** | 70.5 | 102.2 | 150.3 | 8.13µV | 13.3µV |
| **2 R1 1,0 deg** | 76.3 | 104.5 | 150.3 | 3.94µV (!) | 11.7µV |
| **3 L1 2,0 deg** | 62.8 | 108.0 | 130.3 | 6.51µV (!) | 4.14µV |
| **4 L1 1,0 deg** | 70.5 | 118.0 (!) | 125.6 | 83.5nV (!) | 2.01µV |

### 2_Pattern-VEP 30 min&15 min ( Monitor )

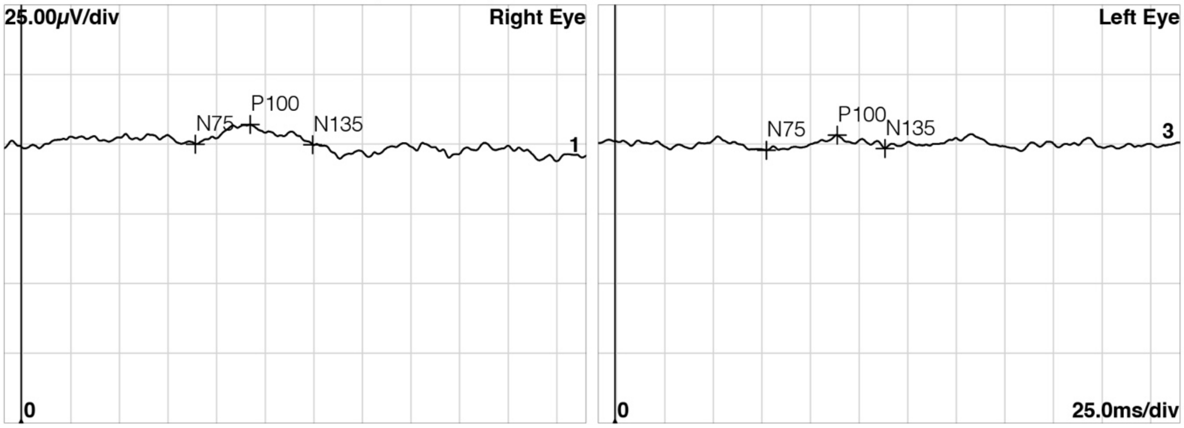


| Normals | - | 105-126 | - | 7.00µV-42.5µV | |
|---|---|---|---|---|---|
| **Channel** | **N75 [ms]** | **P100 [ms]** | **N135 [ms]** | **N75-P100** | **P100-N135** |
| **1 R1 30 min** | 89.8 | 118.0 | 150.3 | 7.13µV | 7.29µV |
| **2 R1 15 min** | | | | | |
| **3 L1 30 min** | 78.1 | 114.5 | 139.1 | 5.40µV (!) | 4.80µV |
| **4 L1 15 min** | | | | | |

B

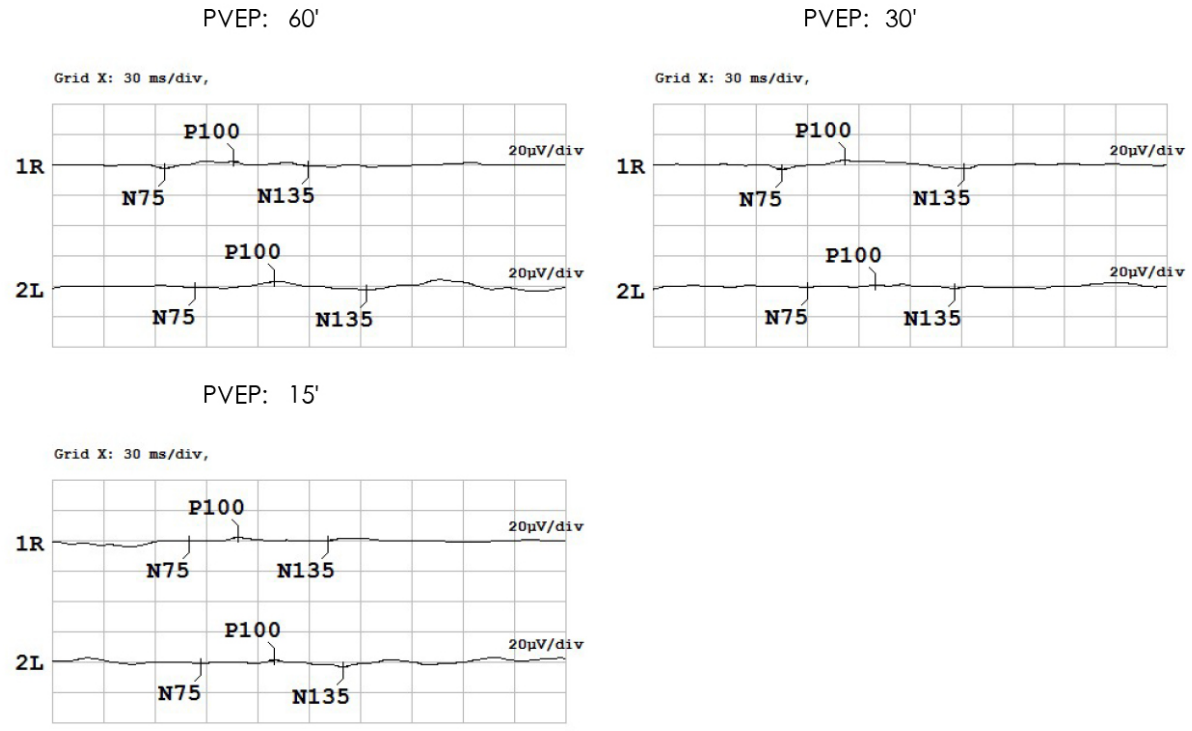


PVEP: 图形翻转,60'

| No. | N75 [ms] | P100 [ms] | N135 [ms] | N75-P100 [μV] | P100-N135 [μV] |
|---|---|---|---|---|---|
| 1R | 66.0 | 106.0 | 149.5 | 4.9 | 3.4 |
| 2L | 83.5 | 130.0 | 183.5 | 4.8 | 5.8 |

PVEP: 图形翻转,30'

| No. | N75 [ms] | P100 [ms] | N135 [ms] | N75-P100 [μV] | P100-N135 [μV] |
|---|---|---|---|---|---|
| 1R | 75.5 | 112.0 | 182.0 | 6.3 | 6.0 |
| 2L | 90.5 | 130.0 | 176.0 | 2.8 | 3.1 |

PVEP: 图形翻转,15'

| No. | N75 [ms] | P100 [ms] | N135 [ms] | N75-P100 [μV] | P100-N135 [μV] |
|---|---|---|---|---|---|
| 1R | 80.0 | 109.0 | 161.5 | 3.2 | 1.9 |
| 2L | 87.0 | 130.0 | 170.0 | 3.0 | 4.9 |

**Fig. S7. Retina ERG of patient 3.** The light-stimulated electrical activity of the retina indicates no remarkable change in terms of retinal function in patient 3 (**A**: pre-injection; **B**: six months post-injection).

PVEP: 60'

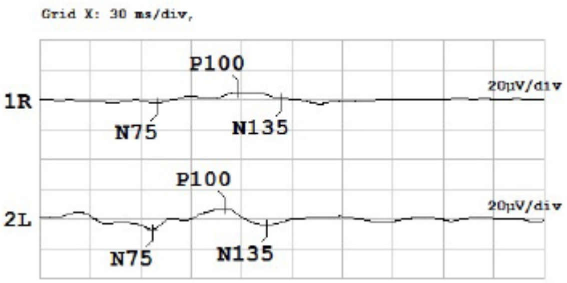


PVEP: 30'

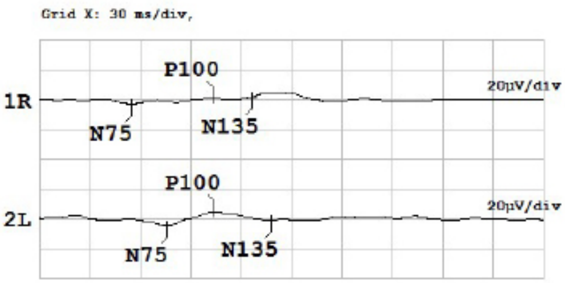


PVEP: 15'

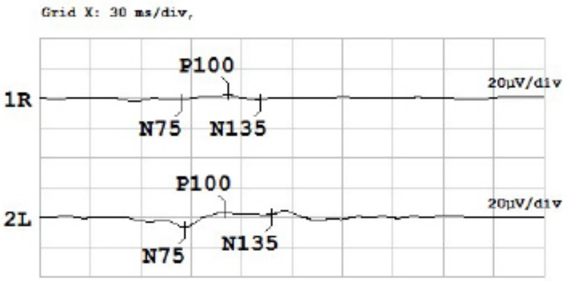


PVEP: 图形翻转.60'

| No. | N75 [ms] | P100 [ms] | N135 [ms] | N75-P100 [µV] | P100-N135 [µV] |
|---|---|---|---|---|---|
| 1R | 70.0 | 118.5 | 144.5 | 7.9 | 4.2 |
| 2L | 67.0 | 110.0 | 135.0 | 14.6 | 11.8 |

PVEP: 图形翻转.30'

| No. | N75 [ms] | P100 [ms] | N135 [ms] | N75-P100 [µV] | P100-N135 [µV] |
|---|---|---|---|---|---|
| 1R | 55.0 | 103.5 | 126.5 | 4.7 | -0.6 |
| 2L | 76.0 | 104.0 | 138.0 | 9.8 | 6.4 |

PVEP: 图形翻转.15'

| No. | N75 [ms] | P100 [ms] | N135 [ms] | N75-P100 [µV] | P100-N135 [µV] |
|---|---|---|---|---|---|
| 1R | 84.0 | 112.0 | 131.5 | 3.9 | 4.1 |
| 2L | 86.0 | 110.0 | 138.0 | 10.5 | 1.2 |

**Fig. S8. Retina ERG of patient 2.** The light-stimulated electrical activity of retina indicates no significant change in terms of retinal function in patient 2 twelve months after the treatment.

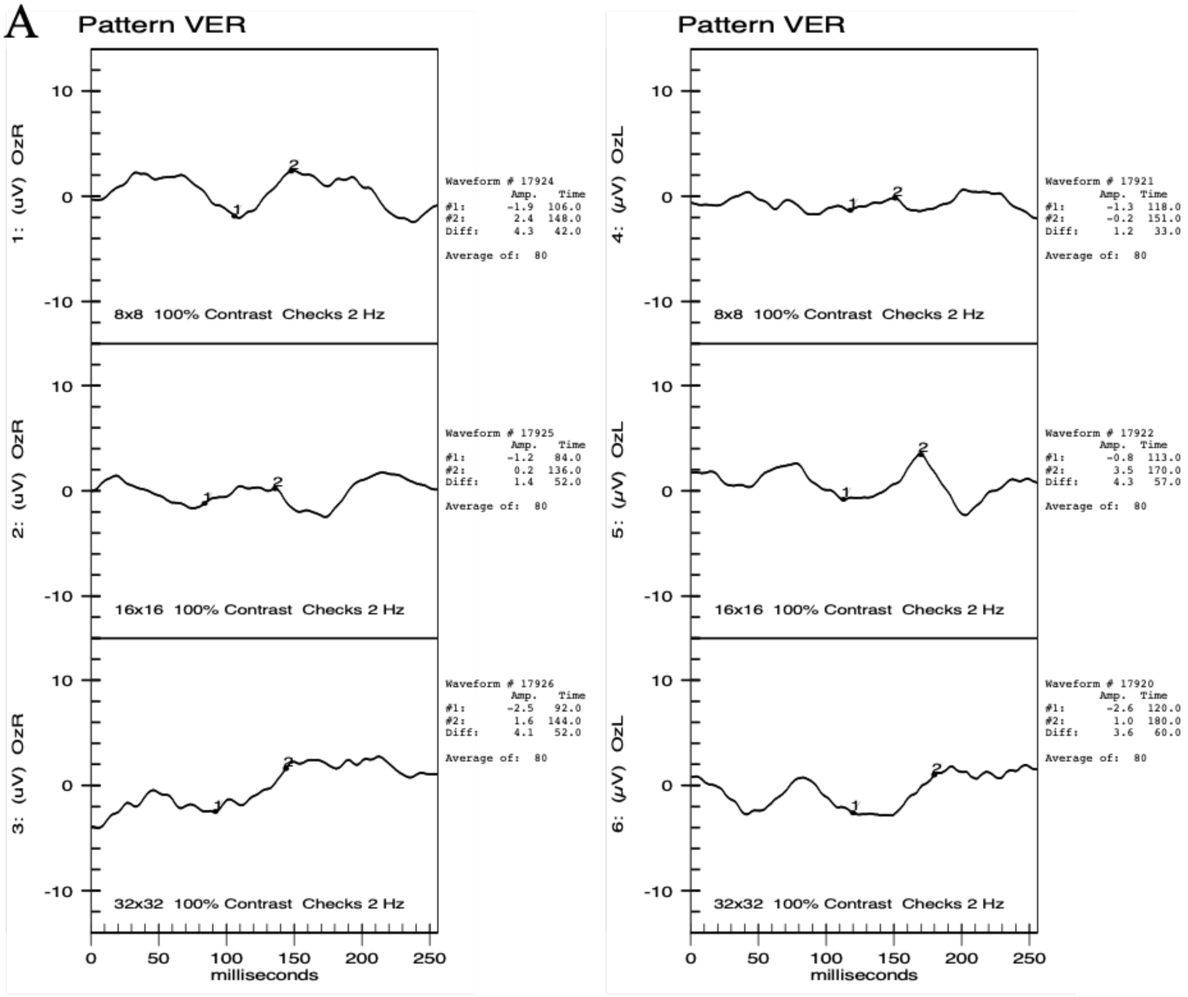
A
Pattern VER
1: (uV) OzR
8x8 100% Contrast Checks 2 Hz
Waveform # 17924
Amp. Time
#1: -1.9 106.0
#2: 2.4 148.0
Diff: 4.3 42.0
Average of: 80
2: (uV) OzR
16x16 100% Contrast Checks 2 Hz
Waveform # 17925
Amp. Time
#1: -1.2 84.0
#2: 0.2 136.0
Diff: 1.4 52.0
Average of: 80
3: (uV) OzR
32x32 100% Contrast Checks 2 Hz
Waveform # 17926
Amp. Time
#1: -2.5 92.0
#2: 1.6 144.0
Diff: 4.1 52.0
Average of: 80
milliseconds
Pattern VER
4: (μV) OzL
8x8 100% Contrast Checks 2 Hz
Waveform # 17921
Amp. Time
#1: -1.3 118.0
#2: -0.2 151.0
Diff: 1.2 33.0
Average of: 80
5: (μV) OzL
16x16 100% Contrast Checks 2 Hz
Waveform # 17922
Amp. Time
#1: -0.8 113.0
#2: 3.5 170.0
Diff: 4.3 57.0
Average of: 80
6: (μV) OzL
32x32 100% Contrast Checks 2 Hz
Waveform # 17920
Amp. Time
#1: -2.6 120.0
#2: 1.0 180.0
Diff: 3.6 60.0
Average of: 80
milliseconds

**B**

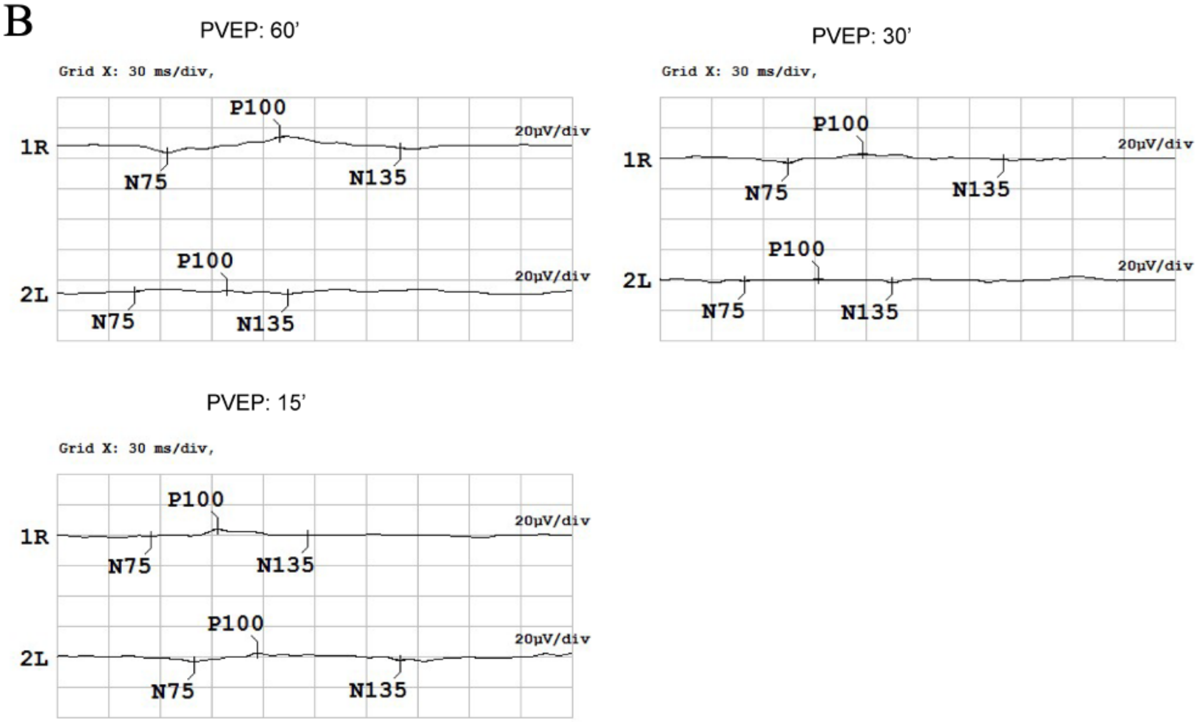


PVEP: 图形翻转,60'

| No. | N75 [ms] | P100 [ms] | N135 [ms] | N75-P100 [µV] | P100-N135 [µV] |
|---|---|---|---|---|---|
| 1R | 64.5 | 130.0 | 200.0 | 10.9 | 7.9 |
| 2L | 45.0 | 99.5 | 134.5 | 0.7 | 2.7 |

PVEP: 图形翻转,30'

| No. | N75 [ms] | P100 [ms] | N135 [ms] | N75-P100 [µV] | P100-N135 [µV] |
|---|---|---|---|---|---|
| 1R | 74.5 | 118.0 | 200.0 | 6.2 | 4.2 |
| 2L | 49.5 | 92.5 | 135.5 | 1.6 | 2.7 |

PVEP: 图形翻转,15'

| No. | N75 [ms] | P100 [ms] | N135 [ms] | N75-P100 [µV] | P100-N135 [µV] |
|---|---|---|---|---|---|
| 1R | 55.0 | 94.0 | 146.0 | 5.1 | 4.7 |
| 2L | 80.0 | 117.0 | 200.0 | 6.0 | 5.0 |

**Fig. S9. Retinal ERG examination of patient 1.** ERG detected no obvious changes of rod or cone responses to light stimulus in patient 1. (**A**: pre-injection; **B**: twelve months post-injection)

**Table S3. Intraocular bacterial and fungal culture results of patient 2.** Gram+ cocci were found in the right eye of patient 2 during the vitrectomy 6 months after PK.

| Culture | Presence | Species |
|---|---|---|
| **Bacteria** | + | Gram-positive cocci |
| **Fungus** | - | - |

### *Patient 1*

A

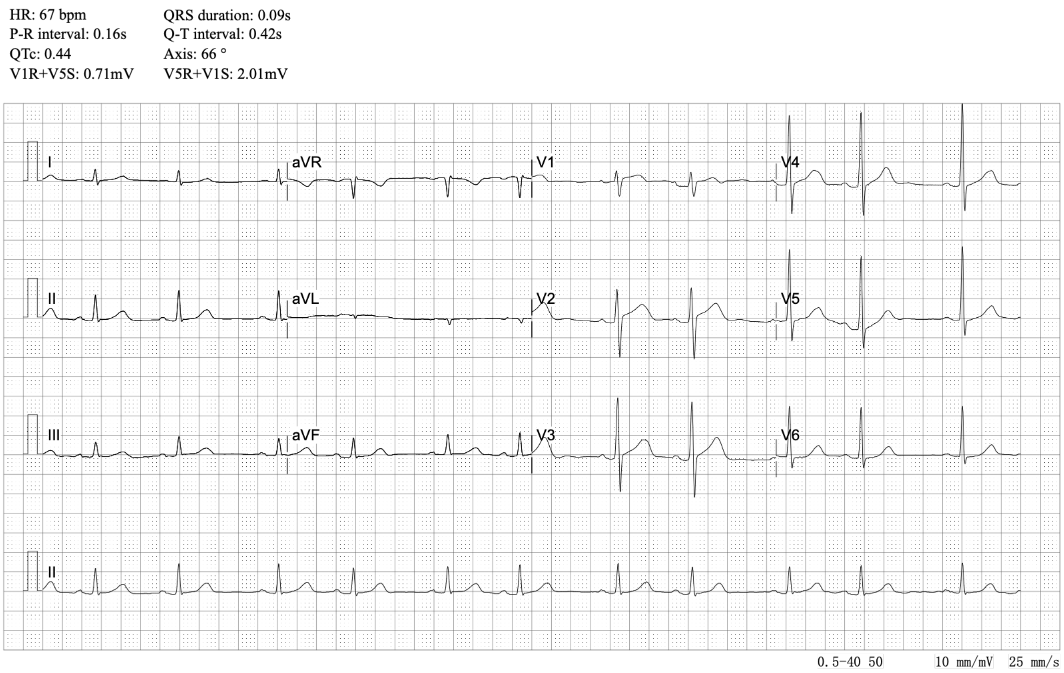


B

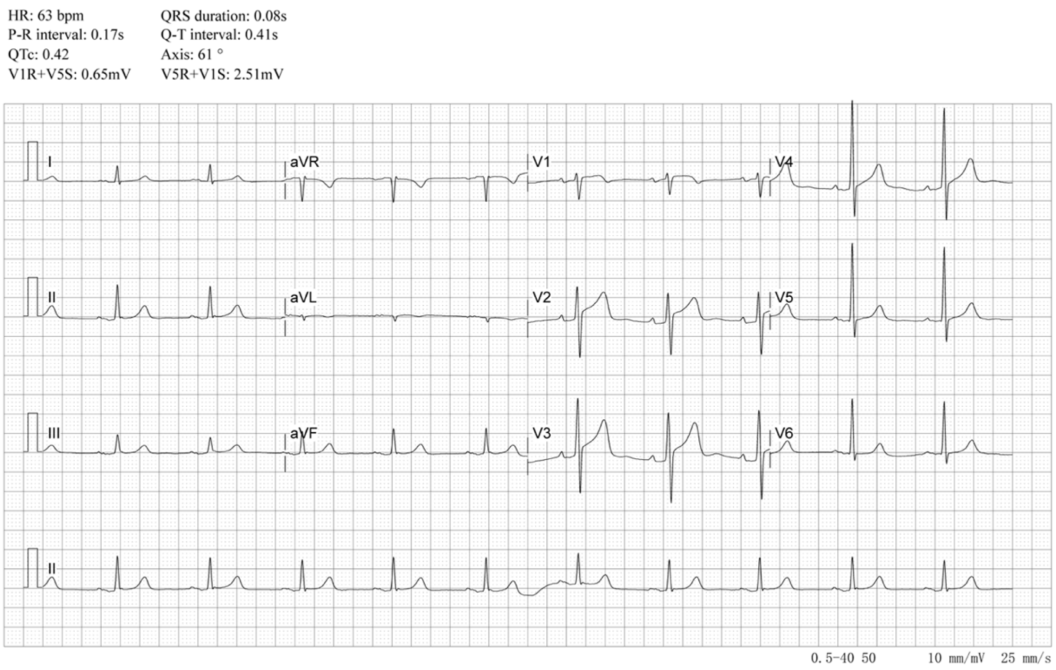


**Fig. S10. The ECG examination.** The pre- and postoperative ECG of patient 1 was unremarkable. (**A**: pre-injection; **B**: twelve months post-injection)

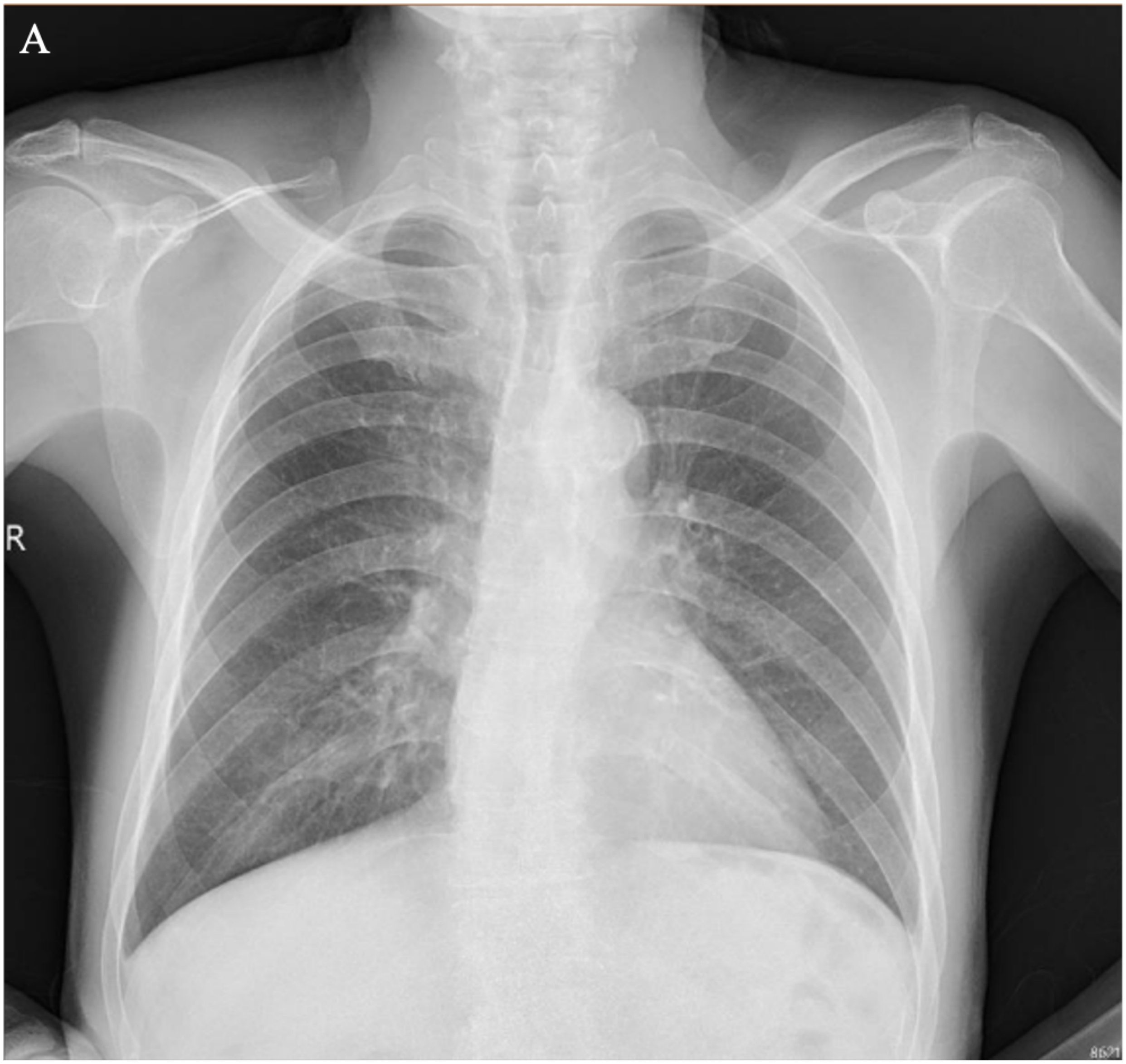
A
R

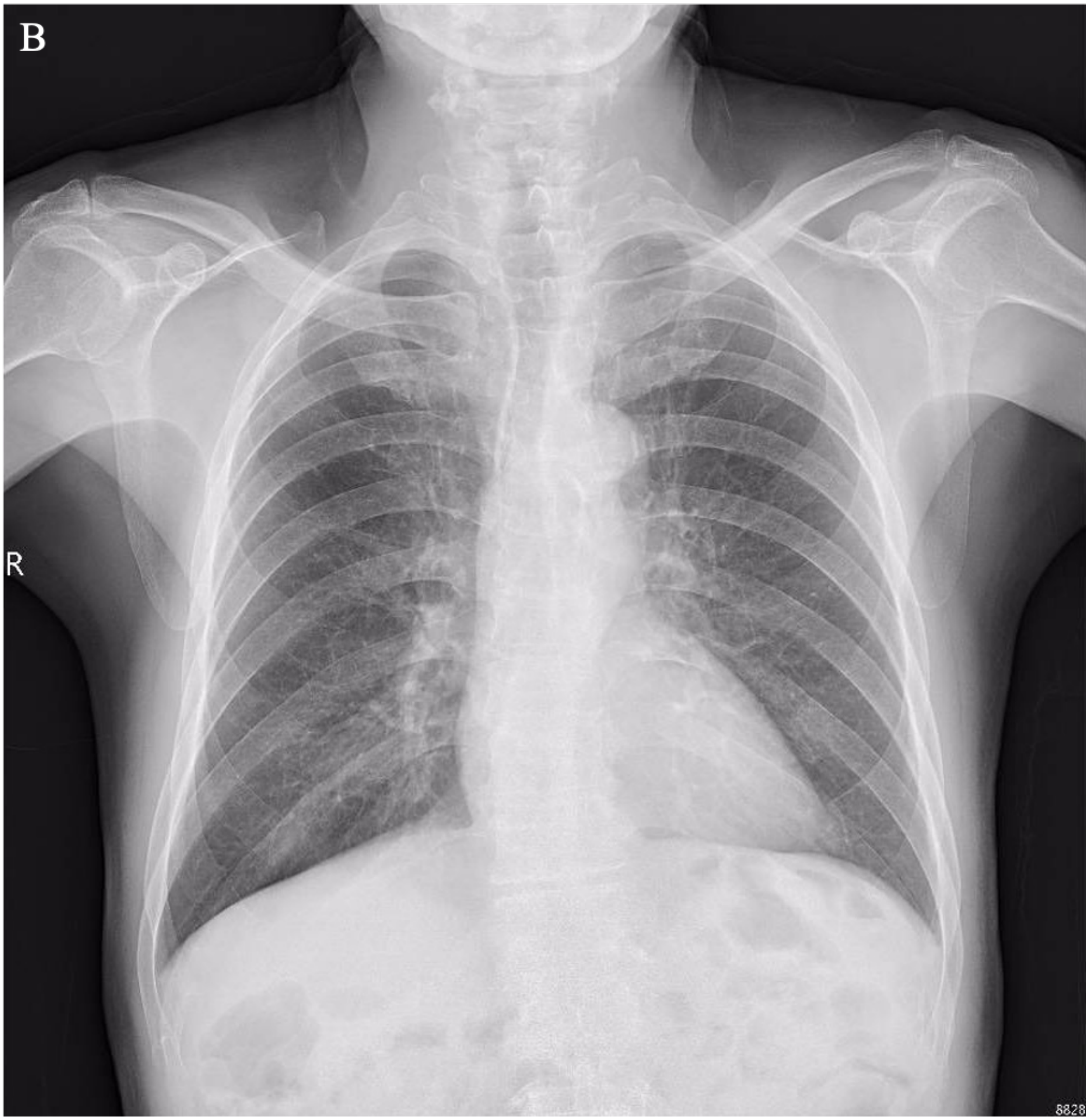


**Fig. S11. Chest X-ray examination.** The Chest X-ray of patient 1 showed no remarkable changes before (**A**) and twelve months (**B**) after the injection.

**Table S4. Blood tests.** The blood tests of patient 1 showed slight anemia, hypoproteinemia, and diabetes before the injection. We observed no abnormal changes which were related to the injection in these blood tests at 12-month follow-up.

| Code | Pre-Injection | Twelve Months Post-Injection | Normal Range | Unit |
|---|---|---|---|---|
| White blood cell count | 5.24 | 4.39 | 4.0~10.0 | 10^9/L |
| Red blood cell count | 2.71 ↓ | 3.47 ↓ | 4.3~5.8 | 10^12/L |
| Hemoglobin | 106 ↓ | 122 ↓ | 130~175 | g/l |
| Packed red blood cell volume | 30.7 ↓ | 36.9 ↓ | 40~50 | % |
| Red blood cell volume distribution width-coefficient of variation | 15.1 ↑ | 15.2 ↑ | 10.0~15.0 | % |
| Red blood cell volume distribution width-standard deviation | 62.3 ↑ | 59.8 ↑ | 35.0~50.0 | fL |
| Mean corpuscular volume | 113.3 ↑ | 106.3 ↑ | 80.0~100.0 | fL |
| Mean corpuscular hemoglobin | 39.1 ↑ | 35.2 ↑ | 27.0~33.0 | Pg |
| Mean corpuscular hemoglobin concentration | 345 | 331 | 320~360 | g/l |
| Platelet count | 175 | 213 | 100~400 | 10^9/L |
| Platelet distribution width | 9.7 | 11.1 | 9.0~17.00 | |
| Thrombocytocrit | 0.17 | 0.22 | 0.16~0.22 | % |
| Mean platelet volume | 9.7 | 10.2 | 9.0~16.0 | fL |
| Platelet-larger cell ratio | 21.6 | 25.0 | 14.0~46.0 | % |
| Percentage of neutrophil | 61.6 | 56.2 | 50.0~70.0 | % |
| Percentage of lymphocyte | 32.1 | 30.8 | 20.0~40.0 | % |
| Percentage of eosinophil | 1.5 | 3.0 | 0.5~5.0 | % |
| Percentage of monocyte | 4.4 | 8.9 ↑ | 3.0~8.0 | % |
| Percentage of basophil | 0.4 | 1.1 ↑ | 0.0~1.0 | % |
| Neutrophil count | 3.23 | 2.47 | 2.0~7.0 | 10^9/L |
| Lymphocyte count | 1.68 | 1.35 | 0.8~4.0 | 10^9/L |
| Monocyte count | 0.23 | 0.39 | 0.10~0.80 | 10^9/L |
| Eosinophil count | 0.08 | 0.13 | 0.00~0.50 | 10^9/L |
| Basophil count | 0.02 | 0.05 | 0.00~0.10 | 10^9/L |

| Code | Pre-Injection | Twelve Months Post-Injection | Normal Range | Unit |
|---|---|---|---|---|
| Alanine aminotransferase | 22 | 31 | 0~65 | u/l |
| Aspartate aminotransferase | 25 | 30 | 15~37 | u/l |
| Total protein | 67 | 71 | 64~82 | g/l |

| | | | | | | |
|---|---|---|---|---|---|---|
| **Albumin** | 47 | | 43 | | 35~54 | g/l |
| **Globulin** | 20 | | 28 | | 20~40 | g/l |
| **Albumin/globulin** | 2.4 | | 1.5 | | 1.2~2.5 | |
| **γ-Glutamyltransferase** | 55 | | 37 | | 15~85 | u/l |
| **Prealbumin** | 150 | ↓ | 200 | | 200~400 | mg/l |
| **Alkaline phosphatase** | 134 | | 137 | ↑ | 50~136 | u/l |
| **Blood urea nitrogen** | 6.8 | ↑ | 9.2 | ↑ | 2.5~6.4 | mmol/l |
| **Creatinine** | 78 | | 90 | | 53~115 | umol/l |
| **Uric acid** | 0.42 | ↑ | 0.38 | ↑ | 0.202~0.417 | mmol/l |
| **Total bilirubin** | 10 | | 6 | | 0~17 | umol/l |
| **Connect bilirubin** | 5 | | 2 | | 1~5 | umol/l |
| **Total bile acid** | 14.5 | ↑ | 14.4 | ↑ | 0.0~10.0 | umol/l |
| **High density lipoprotein** | 1.73 | | 1.86 | | 0.910~2.060 | mmol/l |
| **Low density lipoprotein** | 1.33 | | 2.21 | | 0.00~3.36 | mmol/l |
| **Apoprotein A** | 1.53 | | 1.28 | | 1.100~1.700 | g/l |
| **Apoprotein B** | 0.56 | ↓ | 0.76 | ↓ | 0.800~1.550 | g/l |
| **Apoprotein E** | 35 | | 38 | | 27~45 | mg/l |
| **Lipoprotein small a** | 25 | | 42 | | 0~300 | mg/l |
| **Small dense low density lipoprotein** | 0.74 | | 1.20 | | 0.26~1.36 | mmol/l |
| **Calcium** | 2.21 | | 2.21 | | 2.04~2.74 | mmol/l |
| **Phosphorus** | 1.24 | | 1.15 | | 0.80~1.60 | mmol/l |
| **Potassium** | 4.6 | | 4.8 | | 3.5~5.4 | mmol/l |
| **Sodium** | 145 | | 143 | | 135~147 | mmol/l |
| **Chloride** | 109 | ↑ | 103 | | 96~108 | mmol/l |
| **Carbon dioxide binding capacity** | 22 | | 24 | | 21~32 | mmol/l |
| **Lactate dehydrogenase** | 172 | | 153 | | 81~234 | u/l |
| **Creatine kinase** | 75 | | 95 | | 39~308 | u/l |
| **Complement 3c** | 0.73 | ↓ | 0.71 | ↓ | 0.9~1.8 | g/l |
| **Complement 4** | 0.20 | | 0.16 | | 0.1~0.4 | g/l |
| **Complement 1q** | 108.7 | ↓ | 117.7 | ↓ | 159~233 | mg/l |
| **Total complement** | 68.7 | ↑ | 62.7 | ↑ | 32.5~58.3 | u/ml |
| **Haptoglobin** | 56.50 | | 92.00 | | 32~205 | mg/dl |
| **Blood glucose** | 6.1 | | 6.9 | ↑ | 3.9~6.1 | mmol/l |
| **Total cholesterol** | 3.04 | | 4.54 | | 2.80~5.20 | mmol/l |
| **Triacylglycerol** | 0.77 | | 0.70 | | 0.34~2.26 | mmol/l |

| Code | Pre-Injection | Twelve Months Post-Injection | Normal Range | Unit |
|---|---|---|---|---|
| **Prothrombin time** | 13.7 | 12.7 | 11.0~14.5 | sec |
| **International normalized ratio** | 1.03 | 0.98 | 0.80~1.20 | INR |
| **Activated partial thromboplastin time** | 35.9 | 32.6 | 28~45 | s |
| **Thrombin time** | 16.3 | 17.2 | 14.0~21.0 | s |
| **Fibrinogen** | 2.70 | 3.01 | 2.00~4.00 | g/l |
| **D-Dimer** | 0.67 ↑ | 0.75 ↑ | 0.00~0.50 | ug/ml |
| **Prothrombin time ratio** | 95 | 105 | 70~150 | % |
| **Code** | **Pre-Injection** | **Twelve Months Post-Injection** | **Normal Range** | **Unit** |
| **Glycated hemoglobin** | 6.2 ↑ | 6.2 ↑ | 4.0~6.0 | % |

*Red font denotes an elevated clinical index than the normal range; pink font denotes a decreased clinical index than the normal range. Similarly hereinafter.

**Table S5. Urine tests.** The pre-injection and twelve months post-injection routine urinalysis of patient 1 was unremarkable.

| Code | Pre-Injection | Twelve Months Post-Injection | Normal Range | Unit |
|---|---|---|---|---|
| **Urine glucose** | - | - | - | |
| **Ketone body** | - | - | - | |
| **Occult blood** | - | - | - | |
| **Protein** | - | - | - | |
| **Nitrite** | - | - | - | |
| **Bilirubin** | - | - | - | |
| **Specific gravity** | >=1.030 ↑ | 1.020 | 1.003~1.030 | |
| **Urine PH value** | 6.0 | 6.0 | 5.0~6.5 | |
| **Urobilinogen** | 16 | 16 | 3.0~16.0 | umol/l |
| **Leukocyte** | - | - | - | |
| **White blood cell count** | 2 | 2 | 0~28 | /ul |
| **Red blood cell count** | 1 | 6 | 0~17 | /ul |
| **Squamous epithelial cells** | - | 2 | 0~28 | /ul |
| **Non-squamous epithelial cells** | - | 1 | 0~6 | /ul |
| **Trichomonas** | - | - | 0~1 | /ul |
| **Kidney epithelial cells** | - | - | 0~6 | /ul |
| **Transparent tube** | - | - | 0~2 | /ul |
| **Particle tube** | - | - | 0~1 | /LPF |
| **Cell tube** | - | - | 0~1 | /LPF |
| **Triphosphate crystal** | - | - | | |
| **Calcium oxalate crystal** | - | - | | |
| **Leucine crystal** | - | - | | |
| **Cystine crystals** | - | - | | |

**Table S6. Infectious diseases tests.** The four transfusion-associated contagion tests (hepatitis B, hepatitis C, syphilis, AIDS) of patient 1 were unremarkable.

| Code | Pre-Injection | Twelve Months Post-Injection |
|---|---|---|
| **Hepatitis B virus surface antigen** | Negative | Negative |
| **Hepatitis B virus surface antibody** | Negative | Negative |
| **Hepatitis B virus e antigen** | Negative | Negative |
| **Hepatitis B virus e antibody** | Negative | Negative |
| **Hepatitis B virus core antibody** | Negative | Negative |
| **Hepatitis B virus antibody-immunoglobin M** | Negative | Negative |
| **Hepatitis B virus pre-S1 antigen** | Negative | Negative |
| **Hepatitis C virus antibody** | Negative | Negative |
| **Treponema pallidum particle agglutination test** | Negative | Negative |
| **Rapid plasma regain test** | Negative | Negative |
| **Human immunodeficiency virus antibody** | Negative | Negative |

### *Patient 2*

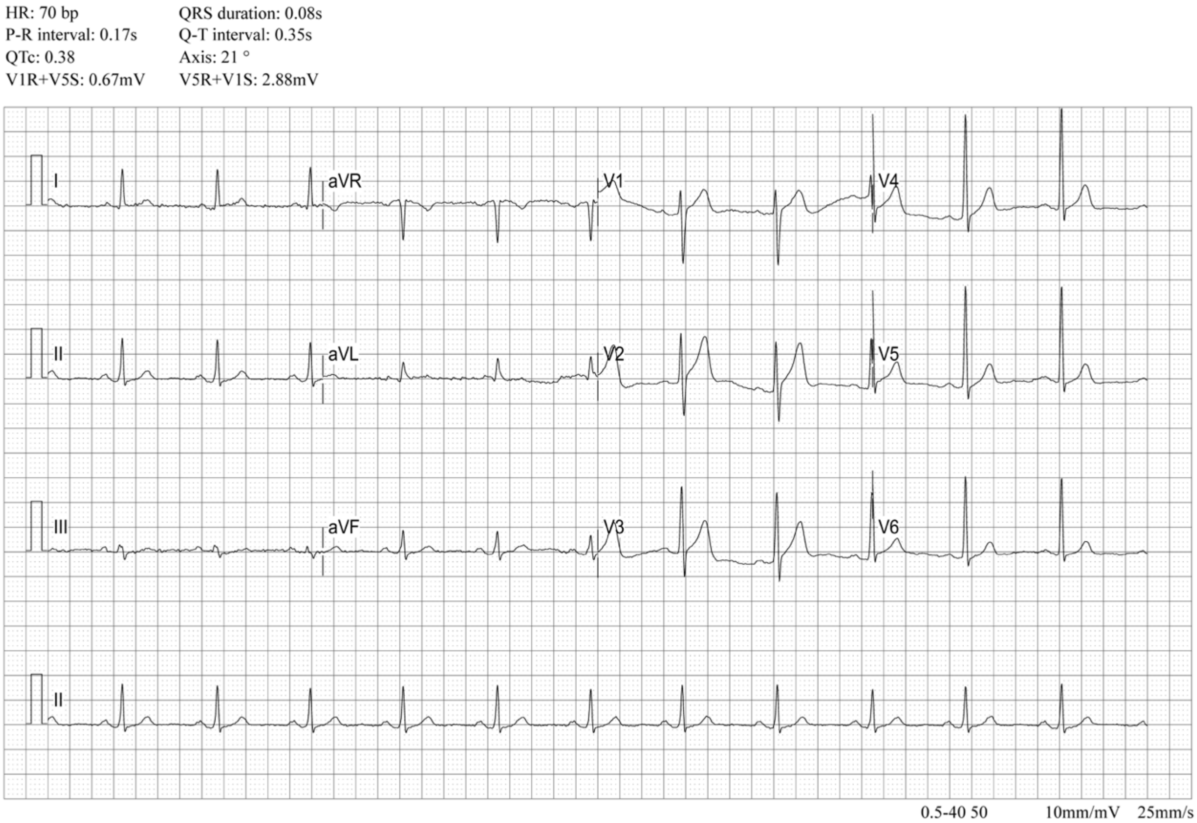


**Fig. S12. The ECG.** The ECG of patient 2 showed counterclockwise transposition, indicating a possibly hypertrophic in the left ventricle. Slightly hypertrophy of the ventricle is not reckoned as a contraindication for corneal transplant surgery.

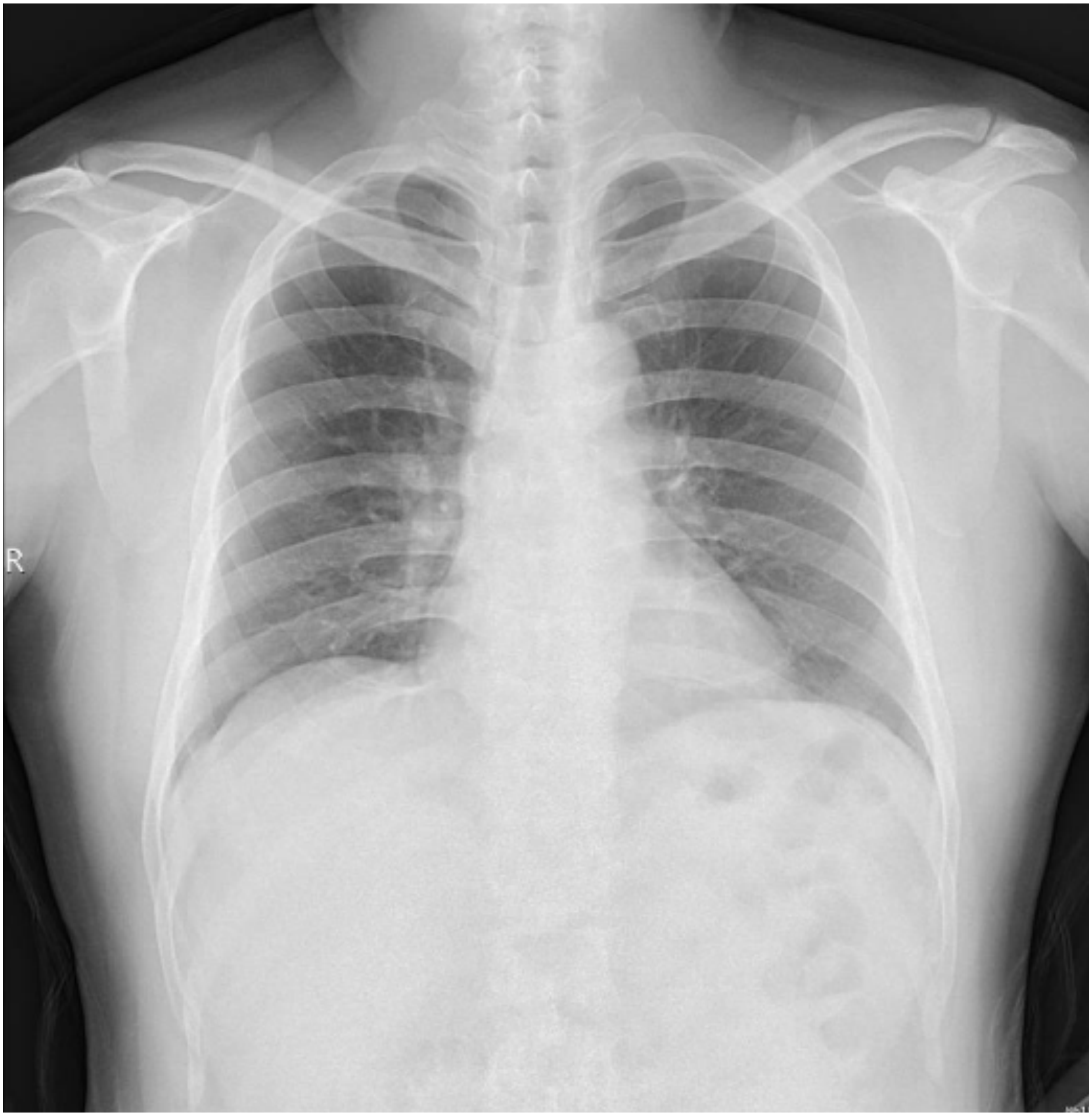


**Fig. S13. Chest X-ray film.** The chest X-ray result of patient 2 was unremarkable.

**Table S7. Blood tests.** The blood tests of patient 2 indicated mild hyperlipidemia, which had no obvious influence on the implementation of the surgery. We observed no notable changes related to the injection in these blood tests at 12-month follow-up.

| Code | Pre-Injection | Twelve Months Post-Injection | Normal Range | Unit |
|---|---|---|---|---|
| **White blood cell count** | 7.17 | 10.16 ↑ | 4.0~10.0 | 10^9/L |
| **Red blood cell count** | 4.73 | 4.69 | 4.3~5.8 | 10^12/L |
| **Hemoglobin** | 148 | 144 | 130~175 | g/l |
| **Packed red blood cell volume** | 44.0 | 43.1 | 40~50 | % |
| **Red blood cell volume distribution width-coefficient of variation** | 12.1 | 12.9 | 10.0~15.0 | % |
| **Red blood cell volume distribution width-standard deviation** | 41.6 | 43.1 | 35.0~50.0 | fL |
| **Mean corpuscular volume** | 93.0 | 91.9 | 80.0~100.0 | fL |
| **Mean corpuscular hemoglobin** | 31.3 | 30.7 | 27.0~33.0 | Pg |
| **Mean corpuscular hemoglobin concentration** | 336 | 334 | 320~360 | g/l |
| **Platelet count** | 229 | 236 | 100~400 | 10^9/L |
| **Platelet distribution width** | 9.3 | 9.8 | 9.0~17.00 | |
| **Thrombocytocrit** | 0.21 | 0.22 | 0.16~0.22 | % |
| **Mean platelet volume** | 9.1 | 9.2 | 9.0~16.0 | fL |
| **Platelet-larger cell ratio** | 17.7 | 18.4 | 14.0~46.0 | % |
| **Percentage of neutrophil** | 64.3 | 77.8 ↑ | 50.0~70.0 | % |
| **Percentage of lymphocyte** | 28.7 | 18.3 ↓ | 20.0~40.0 | % |
| **Percentage of eosinophil** | 1.0 | 0.3 ↓ | 0.5~5.0 | % |
| **Percentage of monocyte** | 5.6 | 3.1 | 3.0~8.0 | % |
| **Percentage of basophil** | 0.4 | 0.5 | 0.0~1.0 | % |
| **Neutrophil count** | 4.61 | 7.9 ↑ | 2.0~7.0 | 10^9/L |
| **Lymphocyte count** | 2.06 | 1.86 | 0.8~4.0 | 10^9/L |
| **Monocyte count** | 0.40 | 0.32 | 0.10~0.80 | 10^9/L |
| **Eosinophil count** | 0.07 | 0.03 | 0.00~0.50 | 10^9/L |
| **Basophil count** | 0.03 | 0.05 | 0.00~0.10 | 10^9/L |
| **Code** | **Pre-Injection** | **Twelve Months Post-Injection** | **Normal Range** | **Unit** |
| **Alanine aminotransferase** | 17 | 19 | 0~65 | u/l |
| **Aspartate aminotransferase** | 17 | 15 | 15~37 | u/l |
| **Total protein** | 70 | 68 | 64~82 | g/l |

| | | | | | | |
|---|---|---|---|---|---|---|
| **Albumin** | 46 | | 42 | | 35~54 | g/l |
| **Globulin** | 24 | | 26 | | 20~40 | g/l |
| **Albumin/globulin** | 1.9 | | 1.6 | | 1.2~2.5 | |
| **γ-Glutamyltransferase** | 33 | | 37 | | 15~85 | u/l |
| **Prealbumin** | 320 | | 350 | | 200~400 | mg/l |
| **Alkaline phosphatase** | 71 | | 80 | | 50~136 | u/l |
| **Blood urea nitrogen** | 5.5 | | 7.5 | ↑ | 2.5~6.4 | mmol/l |
| **Creatinine** | 69 | | 62 | | 53~115 | umol/l |
| **Uric acid** | 0.25 | | 0.32 | | 0.202~0.417 | mmol/l |
| **Total bilirubin** | 16 | | 10 | | 0~17 | umol/l |
| **Connect bilirubin** | 5 | | 2 | | 1~5 | umol/l |
| **Total bile acid** | 2.5 | | 11.2 | ↑ | 0.0~10.0 | umol/l |
| **High density lipoprotein** | 1.24 | | 1.61 | | 0.910~2.060 | mmol/l |
| **Low density lipoprotein** | 4.36 | ↑ | 4.72 | ↑ | 0.00~3.36 | mmol/l |
| **Apoprotein A** | 1.46 | | 1.24 | | 1.100~1.700 | g/l |
| **Apoprotein B** | 1.51 | | 1.55 | | 0.800~1.550 | g/l |
| **Apoprotein E** | 52 | ↑ | 46 | ↑ | 27~45 | mg/l |
| **Lipoprotein small a** | 41 | | 47 | | 0~75 | nmol/l |
| **Small dense low density lipoprotein** | 3.11 | ↑ | 2.70 | ↑ | 0.26~1.36 | mmol/l |
| **Calcium** | 2.34 | | 2.25 | | 2.04~2.74 | mmol/l |
| **Phosphorus** | 1.18 | | 0.99 | | 0.80~1.60 | mmol/l |
| **Potassium** | 4.2 | | 4.0 | | 3.5~5.4 | mmol/l |
| **Sodium** | 140 | | 142 | | 135~147 | mmol/l |
| **Chloride** | 104 | | 101 | | 96~108 | mmol/l |
| **Carbon dioxide binding capacity** | 23 | | 21 | | 21~32 | mmol/l |
| **Lactate dehydrogenase** | 168 | | 182 | | 81~234 | u/l |
| **Creatine kinase** | 122 | | 128 | | 39~308 | u/l |
| **Complement 3c** | 1.03 | | 1.04 | | 0.9~1.8 | g/l |
| **Complement 4** | 0.20 | | 0.32 | | 0.1~0.4 | g/l |
| **Complement 1q** | 154.5 | ↓ | 169.4 | | 159~233 | mg/l |
| **Total complement** | 67.9 | ↑ | 63.5 | ↑ | 32.5~58.3 | u/ml |
| **Haptoglobin** | 93.30 | | 110.50 | | 32~205 | mg/dl |
| **Blood glucose** | 5.6 | | 6.1 | | 3.9~6.1 | mmol/l |
| **Total cholesterol** | 6.12 | ↑ | 7.04 | ↑ | 2.80~5.20 | mmol/l |
| **Triacylglycerol** | 2.33 | ↑ | 1.81 | | 0.34~2.26 | mmol/l |

| Code | Pre-Injection | Twelve Months Post-Injection | Normal Range | Unit |
|---|---|---|---|---|
| **Prothrombin time** | 13.3 | 12.4 | 11.0~14.5 | sec |
| **International normalized ratio** | 0.99 | 0.95 | 0.80~1.20 | INR |
| **Activated partial thromboplastin time** | 35.3 | 33.0 | 28~45 | s |
| **Thrombin time** | 18.7 | 19.7 | 14.0~21.0 | s |
| **Fibrinogen** | 2.98 | 2.74 | 2.00~4.00 | g/l |
| **D-Dimer** | 0.24 | 0.23 | 0.00~0.50 | ug/ml |
| **Prothrombin time ratio** | 102 | 110 | 70~150 | % |

| Code | Pre-Injection | Twelve Months Post-Injection | Normal Range | Unit |
|---|---|---|---|---|
| **Glycated hemoglobin** | 5.9 | 5.9 | 4.0~6.0 | % |

**Table S8. Routine urine test.** The routine urinalysis of patient 2 was unremarkable.

| Code | Pre-Injection | Twelve Months Post-Injection | Normal Range | Unit |
|---|---|---|---|---|
| **Urine glucose** | - | - | - | |
| **Ketone body** | - | - | - | |
| **Occult blood** | - | - | - | |
| **Protein** | - | - | - | |
| **Nitrite** | - | - | - | |
| **Bilirubin** | - | - | - | |
| **Specific gravity** | >=1.030 ↑ | 1.025 | 1.003~1.030 | |
| **Urine PH value** | 5.5 | 6.0 | 5.0~6.5 | |
| **Urobilinogen** | 3.2 | 16 | 3.0~16.0 | umol/l |
| **Leukocyte** | - | - | - | |
| **White blood cell count** | 6 | 6 | 0~28 | /ul |
| **Red blood cell count** | 2 | 2 | 0~17 | /ul |
| **Squamous epithelial cells** | 2 | 1 | 0~28 | /ul |
| **Non-squamous epithelial cells** | - | 1 | 0~6 | /ul |
| **Trichomonas** | - | - | 0~1 | /ul |
| **Kidney epithelial cells** | - | - | 0~6 | /ul |
| **Transparent tube** | - | - | 0~2 | /ul |
| **Particle tube** | - | - | 0~1 | /LPF |
| **Cell tube** | - | - | 0~1 | /LPF |
| **Triphosphate crystal** | - | - | | |
| **Calcium oxalate crystal** | - | - | | |
| **Leucine crystal** | - | - | | |
| **Cystine crystals** | - | - | | |

**Table S9. Transfusion-associated contagion tests.** The contagion tests of patient 2 showed specific antibodies to the hepatitis B virus. The other three transfusion-associated contagion tests (hepatitis C, syphilis, AIDS) were negative.

| **Code** | **Pre-Injection** | **Twelve Months Post-Injection** |
|---|---|---|
| **Hepatitis B virus surface antigen** | Negative | Negative |
| **Hepatitis B virus surface antibody** | Weak Positive | Weak Positive |
| **Hepatitis B virus e antigen** | Negative | Negative |
| **Hepatitis B virus e antibody** | Negative | Negative |
| **Hepatitis B virus core antibody** | Positive | Positive |
| **Hepatitis B virus antibody-immunoglobin M** | Negative | Negative |
| **Hepatitis B virus pre-S1 antigen** | Negative | Negative |
| **Hepatitis C virus antibody** | Negative | Negative |
| **Treponema pallidum particle agglutination test** | Negative | Negative |
| **Rapid plasma regain test** | Negative | Negative |
| **Human immunodeficiency virus antibody** | Negative | Negative |

### *Patient 3*

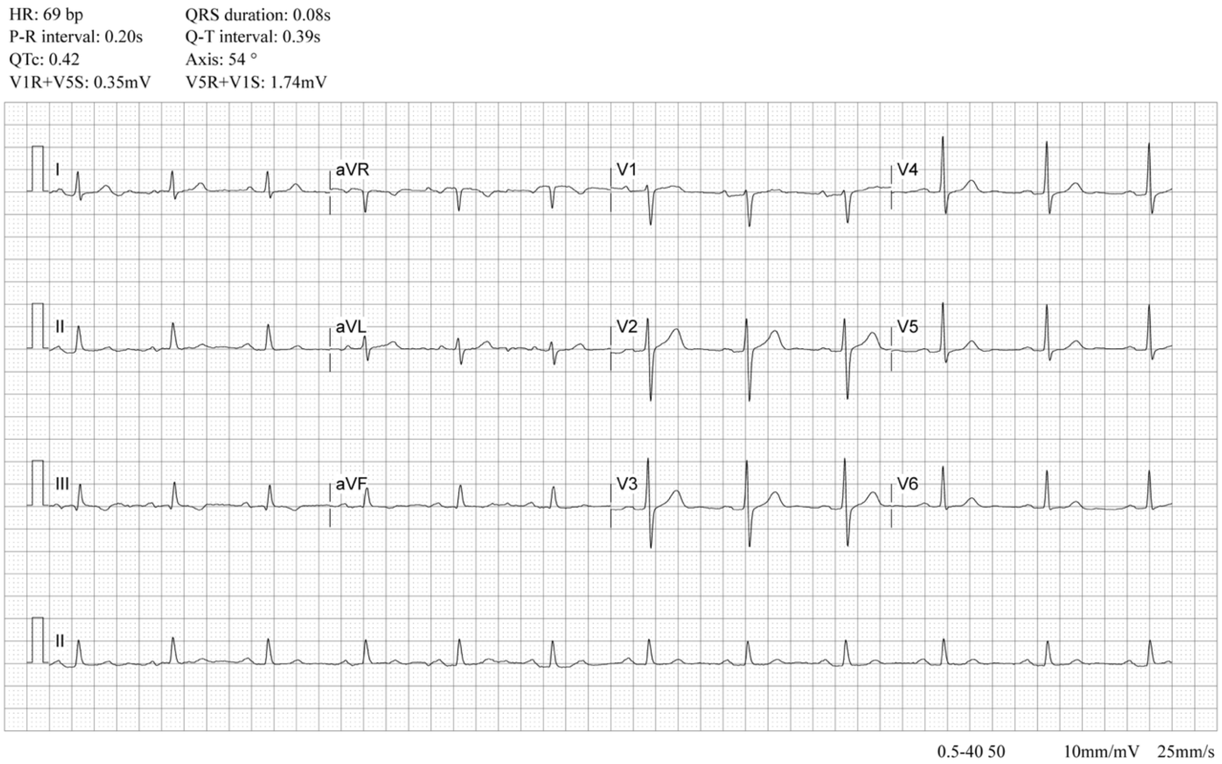


**Fig. S14. The ECG examination.** The pre-operative ECG of patient 3 was unremarkable.

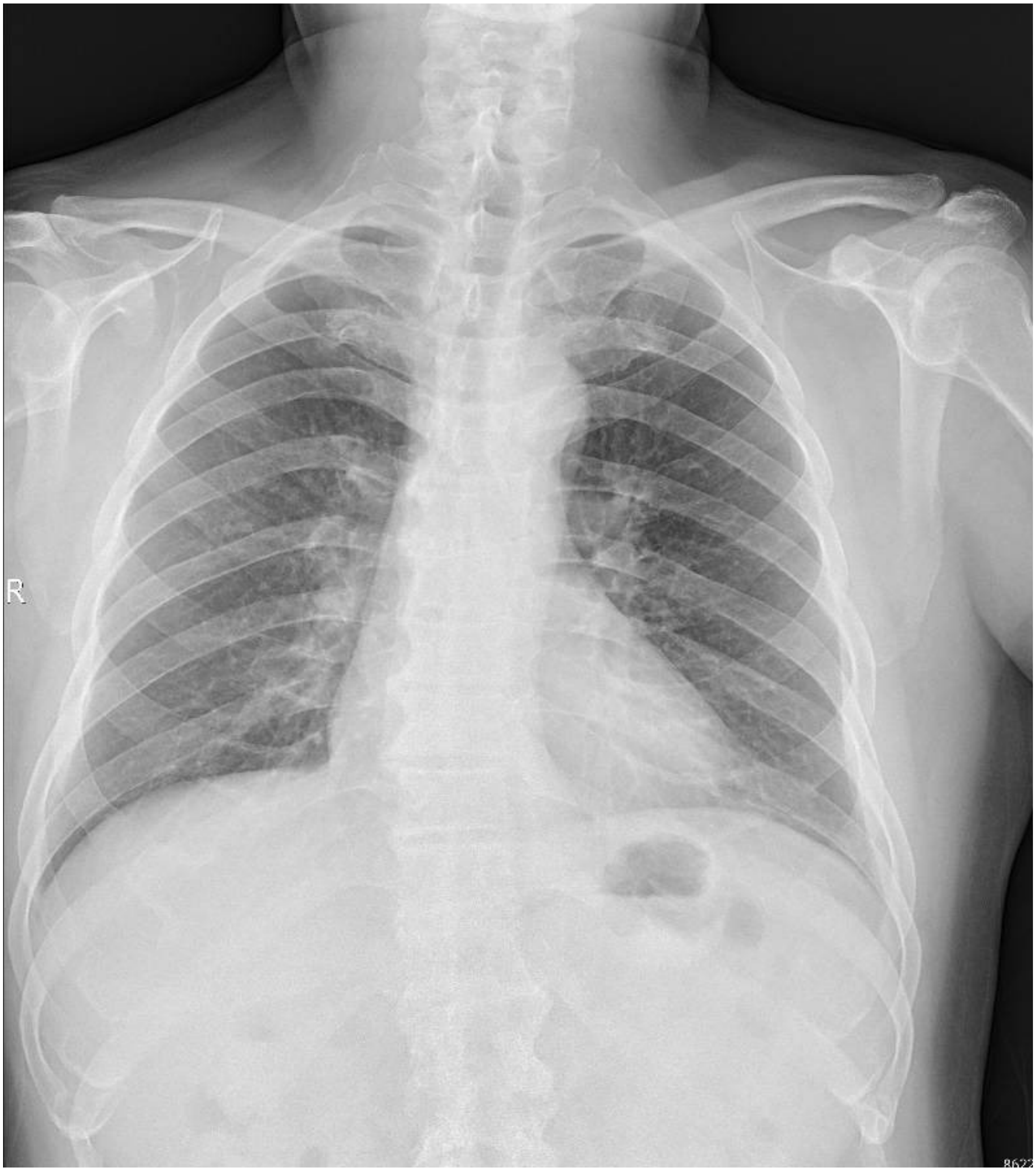


**Fig. S15. Chest X-ray film.** The chest X-ray result of patient 3 was unremarkable.

**Table S10. Blood tests.** The blood tests of patient 3 indicated mild hyperlipidemia, which had no obvious influence on the implementation of the surgery.

| Code | Pre-Injection | Six Months Post-Injection | Normal Range | Unit |
|---|---|---|---|---|
| **White blood cell count** | 5.45 | 4.83 | 4.0~10.0 | 10^9/L |
| **Red blood cell count** | 5.28 | 4.89 | 4.3~5.8 | 10^12/L |
| **Hemoglobin** | 159 | 152 | 130~175 | g/l |
| **Packed red blood cell volume** | 46.1 | 42.8 | 40~50 | % |
| **Red blood cell volume distribution width-coefficient of variation** | 11.9 | 12.4 | 10.0~15.0 | % |
| **Red blood cell volume distribution width-standard deviation** | 37.6 | 39.0 | 35.0~50.0 | fL |
| **Mean corpuscular volume** | 87.3 | 87.5 | 80.0~100.0 | fL |
| **Mean corpuscular hemoglobin** | 30.1 | 31.1 | 27.0~33.0 | Pg |
| **Mean corpuscular hemoglobin concentration** | 345 | 355 | 320~360 | g/l |
| **Platelet count** | 151 | 105 | 100~400 | 10^9/L |
| **Platelet distribution width** | 10.8 | 12.2 | 9.0~17.00 | |
| **Thrombocytocrit** | 0.14 ↓ | 0.11 ↓ | 0.16~0.22 | % |
| **Mean platelet volume** | 9.5 | 10.7 | 9.0~16.0 | fL |
| **Platelet-larger cell ratio** | 21.5 | 30.8 | 14.0~46.0 | % |
| **Percentage of neutrophil** | 62.3 | 57.7 | 50.0~70.0 | % |
| **Percentage of lymphocyte** | 29.7 | 32.4 | 20.0~40.0 | % |
| **Percentage of eosinophil** | 1.7 | 2.3 | 0.5~5.0 | % |
| **Percentage of monocyte** | 5.9 | 7.3 | 3.0~8.0 | % |
| **Percentage of basophil** | 0.4 | 0.3 | 0.0~1.0 | % |
| **Neutrophil count** | 3.40 | 2.21 | 2.0~7.0 | 10^9/L |
| **Lymphocyte count** | 1.62 | 1.24 | 0.8~4.0 | 10^9/L |
| **Monocyte count** | 0.32 | 0.28 | 0.10~0.80 | 10^9/L |
| **Eosinophil count** | 0.09 | 0.09 | 0.00~0.50 | 10^9/L |
| **Basophil count** | 0.02 | 0.01 | 0.00~0.10 | 10^9/L |

| Code | Pre-Injection | Six Months Post-Injection | Normal Range | Unit |
|---|---|---|---|---|
| **Alanine aminotransferase** | 29 | 53 | 0~65 | u/l |
| **Aspartate aminotransferase** | 18 | 26 | 15~37 | u/l |
| **Total protein** | 76 | 65 | 64~82 | g/l |
| **Albumin** | 49 | 40 | 35~54 | g/l |

| | | | | | | |
|---|---|---|---|---|---|---|
| **Globulin** | 27 | | 24 | | 20~40 | g/l |
| **Albumin/globulin** | 1.8 | | 1.66 | | 1.2~2.5 | |
| **γ-Glutamyltransferase** | 37 | | 49 | | 15~85 | u/l |
| **Prealbumin** | 350 | | 310 | | 200~400 | mg/l |
| **Alkaline phosphatase** | 95 | | 103 | | 50~136 | u/l |
| **Blood urea nitrogen** | 6.6 | ↑ | 5.6 | | 2.5~6.4 | mmol/l |
| **Creatinine** | 78 | | 77 | | 53~115 | umol/l |
| **Uric acid** | 0.32 | | 0.37 | | 0.202~0.417 | mmol/l |
| **Total bilirubin** | 16 | | 16 | | 0~17 | umol/l |
| **Connect bilirubin** | 7 | ↑ | 3 | | 1~5 | umol/l |
| **Total bile acid** | 6 | | 2 | | 0.0~10.0 | umol/l |
| **High density lipoprotein** | 1.16 | | 0.84 | ↓ | 0.910~2.060 | mmol/l |
| **Low density lipoprotein** | 2.79 | | 2.24 | | 0.00~3.36 | mmol/l |
| **Apoprotein A** | 1.29 | | 1.00 | ↓ | 1.100~1.700 | g/l |
| **Apoprotein B** | 1.000 | | 0.800 | | 0.800~1.550 | g/l |
| **Apoprotein E** | 39 | | 35 | | 27~45 | mg/l |
| **Lipoprotein small a** | 38 | | 42 | | 0~75 | nmol/l |
| **Small dense low density lipoprotein** | 1.90 | ↑ | 1.87 | ↑ | 0.26~1.36 | mmol/l |
| **Calcium** | 2.32 | | 2.32 | | 2.04~2.74 | mmol/l |
| **Phosphorus** | 0.97 | | 1.23 | | 0.80~1.60 | mmol/l |
| **Potassium** | 4.3 | | 4.4 | | 3.5~5.4 | mmol/l |
| **Sodium** | 140 | | 139 | | 135~147 | mmol/l |
| **Chloride** | 102 | | 105 | | 96~108 | mmol/l |
| **Carbon dioxide binding capacity** | 26 | | 23 | | 21~32 | mmol/l |
| **Lactate dehydrogenase** | 149 | | 175 | | 81~234 | u/l |
| **Creatine kinase** | 75 | | 98 | | 39~308 | u/l |
| **Complement 3c** | 1.07 | | 1.12 | | 0.9~1.8 | g/l |
| **Complement 4** | 0.31 | | 0.29 | | 0.1~0.4 | g/l |
| **Complement 1q** | 195.5 | | 210.4 | | 159~233 | mg/l |
| **Total complement** | 82.5 | ↑ | 84.7 | ↑ | 32.5~58.3 | u/ml |
| **Haptoglobin** | 86.50 | | 98.20 | | 32~205 | mg/dl |
| **Blood glucose** | 5.7 | | 4.7 | | 3.9~6.1 | mmol/l |
| **Total cholesterol** | 4.36 | | 3.68 | | 2.80~5.20 | mmol/l |
| **Triacylglycerol** | 1.23 | | 2.24 | | 0.34~2.26 | mmol/l |

| Code | Pre-Injection | Six Months Post-Injection | Normal Range | Unit |
|---|---|---|---|---|
| **Prothrombin time** | 13.3 | 11.1 | 11.0~14.5 | sec |
| **International normalized ratio** | 1.00 | 0.94 | 0.80~1.20 | INR |
| **Activated partial thromboplastin time** | 37.7 | 32.8 | 28~45 | s |
| **Thrombin time** | 18.0 | 16.8 | 14.0~21.0 | s |
| **Fibrinogen** | 3.43 | 2.72 | 2.00~4.00 | g/l |
| **D-Dimer** | 0.28 | 0.31 | 0.00~0.50 | ug/ml |
| **Prothrombin time ratio** | 99 | 97 | 70~150 | % |

| Code | Pre-Injection | Six Months Post-Injection | Normal Range | Unit |
|---|---|---|---|---|
| **Glycated hemoglobin** | 5.3 | 4.89 | 4.0~6.0 | % |

**Table S11. Routine urine test.** The routine urinalysis of patient 3 was unremarkable.

| Code | Pre-Injection Results | | Normal Range | Unit |
|---|---|---|---|---|
| **Urine glucose** | - | | - | |
| **Ketone body** | - | | - | |
| **Occult blood** | - | | - | |
| **Protein** | - | | - | |
| **Nitrite** | - | | - | |
| **Bilirubin** | - | | - | |
| **Specific gravity** | >=1.030 | ↑ | 1.003~1.030 | |
| **Urine PH value** | 7.0 | ↑ | 5.0~6.5 | |
| **Urobilinogen** | 3.2 | | 3.0~16.0 | umol/l |
| **Leukocyte** | - | | - | |
| **White blood cell count** | 1 | | 0~28 | /ul |
| **Red blood cell count** | 1 | | 0~17 | /ul |
| **Squamous epithelial cells** | - | | 0~28 | /ul |
| **Non-squamous epithelial cells** | - | | 0~6 | /ul |
| **Trichomonas** | - | | 0~1 | /ul |
| **Kidney epithelial cells** | - | | 0~6 | /ul |
| **Transparent tube** | - | | 0~2 | /ul |
| **Particle tube** | - | | 0~1 | /LPF |
| **Cell tube** | - | | 0~1 | /LPF |
| **Triphosphate crystal** | - | | | |
| **Calcium oxalate crystal** | - | | | |
| **Leucine crystal** | - | | | |
| **Cystine crystals** | - | | | |

**Table S12. Transfusion-associated contagion tests.** The contagion tests of patient 3 showed specific antibodies to the hepatitis B virus. The other three transfusion-associated contagion tests (hepatitis C, syphilis, AIDS) were negative.

| Code | Pre-Injection Results |
|---|---|
| **Hepatitis B virus surface antigen** | Negative |
| **Hepatitis B virus surface antibody** | Positive |
| **Hepatitis B virus e antigen** | Negative |
| **Hepatitis B virus e antibody** | Positive |
| **Hepatitis B virus core antibody** | Positive |
| **Hepatitis B virus antibody-immunoglobin M** | Negative |
| **Hepatitis B virus pre-S1 antigen** | Negative |
| **Hepatitis C virus antibody** | Negative |
| **Treponema pallidum particle agglutination test** | Negative |
| **Rapid plasma regain test** | Negative |
| **Human immunodeficiency virus antibody** | Negative |

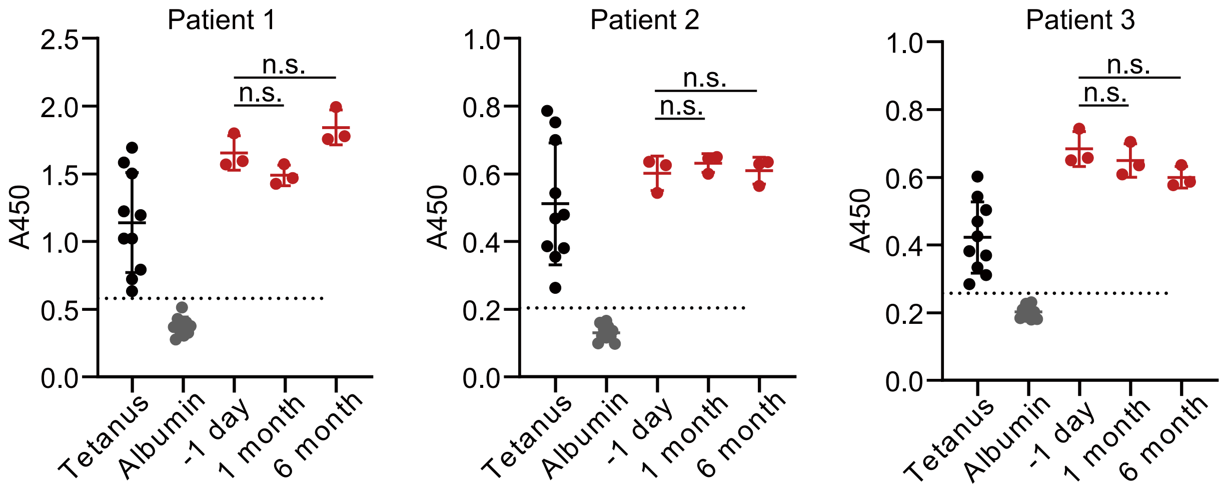


**Fig. S16. ELISA results detect antibodies against SpCas9.** Tetanus toxoid and human albumin in the sera from different donors served as a positive and negative control, respectively. The Cas9-specific antibodies were determined at different time points. The change in Cas9 antibody levels before and after HELP administration was insignificant. All samples above the dotted line were considered antibody-positive. The dotted line represents the mean absorbance of the negative control, human albumin, plus three s.d. from the mean. Data and error bars represent mean ± s.e.m.; n.s., non-significant; unpaired two-tailed Student's t-tests.

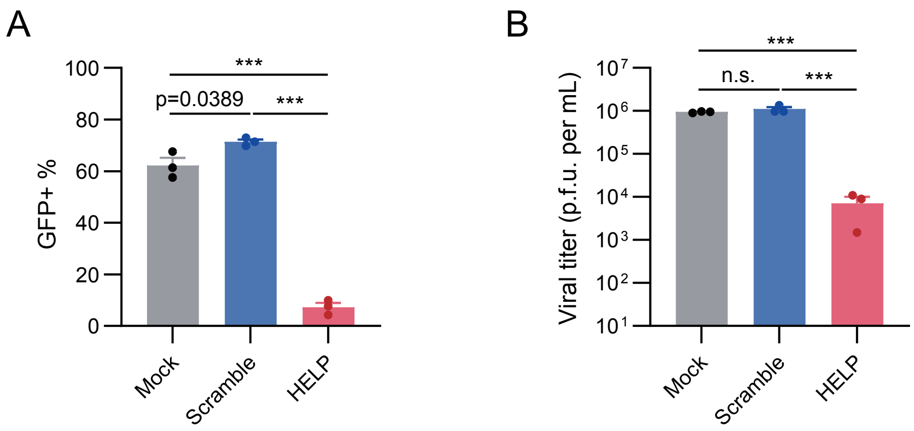


**Fig. S17. In vitro antiviral activity of the clinical grade HELP.** 4 × 104 293T cells were seeded in a 48-well plate and transduced with 400 ng of HELP or scramble control on the following day. The medium was refreshed 12 h post-infection (h.p.i.). 24 h after transduction, cells were infected with HSV-1-GFP at an MOI of 1. The cells and supernatants were harvested at 24 and 48 h.p.i. for flow cytometry (**A**) and plaque assay (**B**), respectively. 293T cells were transduced with HELP for 24 h and then infected with HSV-1-GFP. n=3 biologically independent samples. Data and error bars represent mean ± s.e.m.; n.s., non-significant; ***P<0.001; unpaired two-tailed Student's t-tests.

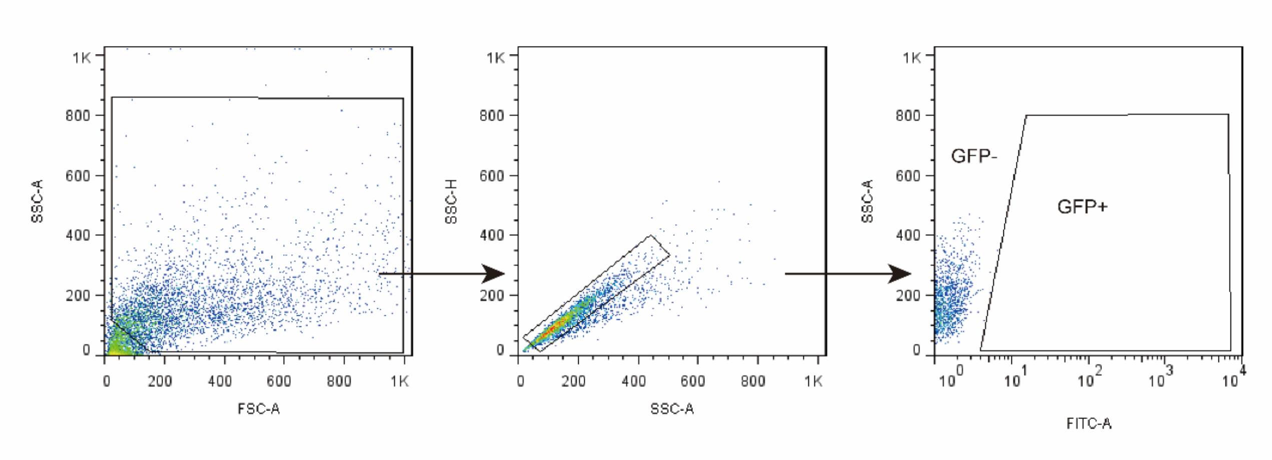


**Fig. S18. Gating strategies used for cell sorting analysis.** Gate strategy to sort GFP positive cells from HSV1-GFP infected 293T cells on supplementary figure S17a.

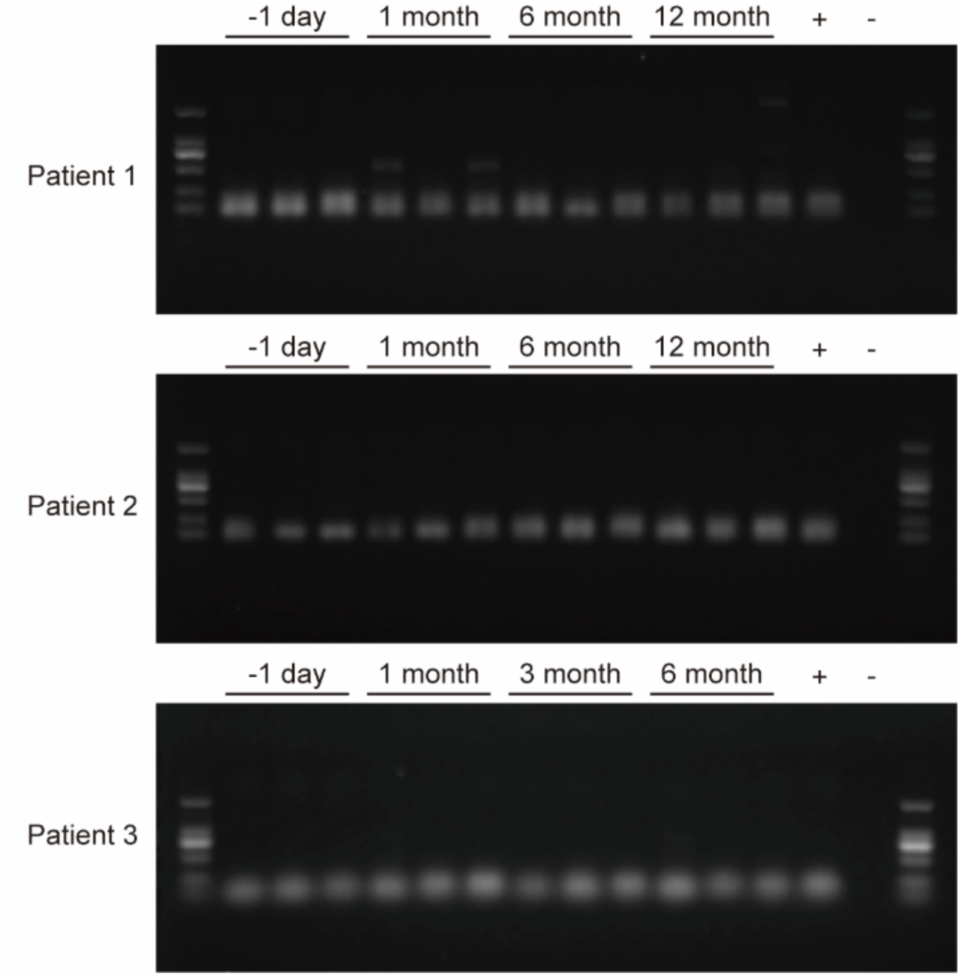


**Fig. S19. Validity of patient samples.** GAPDH gene in the tear swab of patients was tested by PCR to verify the validity of patient samples. Eye swab samples of 1 day pre-injection, 1 month, 6 months, 12 months post-injection of patient 1 & 2, and samples of 1 day pre-injection, 1 month, 3 months, 6 months post-injection of patient 3 were used as templates for PCR followed by nucleic acid gel electrophoresis. The results showed that GAPDH was positive in all samples, indicating that the swabs were valid.

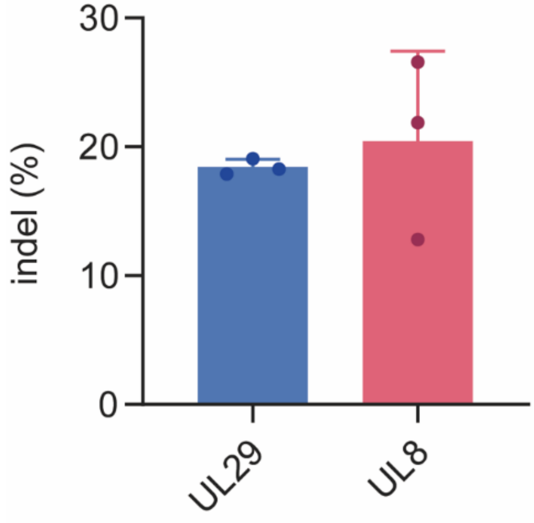


**Fig. S20. Gene editing efficiency of the clinical grade HELP.** TIDE analysis of indels in the HSV-1 genome. 2×104 293T cells were incubated with 200 ng P24 HELP, then infected with HSV-1 with MOI=1. Virus DNA was collected at 2 days post-infection for Sanger sequencing. n=3 biologically independent samples.

**Table S13. The primers for UL8 and UL29 amplification.**

| Target sites | gRNA Sequence (5’-3’) | Primer names | Sequence (5’-3’) |
|---|---|---|---|
| UL8 | GGGGCAGCCATACCGCGTAA | Y1-F | gagccgtagaaatcccgcag |
| | | Y2-R | aaacctcaccaaacagacaa |
| UL29 | GCGAGCGTACACGTATCCC | Y3-F | gggtgtagtccgaaaagccaa |
| | | Y4-R | cacgcccccaggtaaagtgta |

**Table 14. The quality report of HELP.**

| **Source** | OBiO Technology (Shanghai) Corp., Ltd. | **Specification** | 0.25 ml/vial |
|---|---|---|---|
| **Test Category** | **Test** | **Result** | |
| **Quantification** | p24 protein content | 1.19E+04 ng p24/ml | |
| **Safety** | Bacteria | Negative | |
| | Mycoplasma | Negative | |
| **Conclusion** | Qualified | | |

**Table S15. The list of protocol changes.**

| Section of Current protocol | Rationale | Current Version |
|---|---|---|
| Outcomes Measures | Adjust the contents and priorities of outcomes | Primary outcome: HSV-1 testing outcome of the intervention eye; adverse effects.<br>Secondary outcome: patient graft survival; best-corrected visual acuity. |
| 2.1 | Adjust the inclusion criteria | Patients with refractory keratitis caused by herpes virus type I who has acute corneal perforation or had at least one time failed corneal transplant due to the virus relapse. |
