## Supplementary material for "In Vivo CRISPR Gene Editing in Patients with Herpes Stromal Keratitis": Protocol

This trial protocol has been provided by the authors to give readers additional information about their work.

### PROTOCOL

This supplement contains the following items:

1. Original protocol, final protocol, summary of changes.

2. Original and final statistical analysis plan.

### Original Study Protocol

**Protocol ID: 2020023-2-v2**

### Safety and Efficacy of CRISPR/Cas9 mRNA Instantaneous Gene Editing Therapy to Treat Refractory Viral Keratitis

Investigator: Jiaxu Hong, M.D., Ph.D, M.PA.
Deputy Chief Physician
Department of Ophthalmology and Visual Science

Research Institute: Eye, and ENT Hospital,
Shanghai Medical College, Fudan University, Shanghai, China

Founding: National Natural Science Foundation of China,
81970766, 8217040684 and 31971364
the Program for Professor of Special Appointment
(Eastern Scholar) at Shanghai Institutions of Higher Learning,
TP2019046
The Shanghai Innovation Development Program,
2020-RGZN-02033
the Shanghai Key Clinical Research Program,
SHDC2020CR3052B
Startup funding from Shanghai Center for Systems Biomedicine
Shanghai Jiao Tong University, WF220441504

Collaborators: Key Laboratory of Systems Biomedicine,
Shanghai Jiao Tong University

Version 2.0

Version Date: April 12, 2020

**Brief Summary:**

The purpose of this study is to evaluate the safety, tolerability and efficacy of a single dose of CRISPR/Cas9 mRNA Instantaneous Gene Editing Therapy administered via corneal injection in participants with refractory herpetic viral keratitis.

| Condition or disease | Intervention/treatment |
|---|---|
| Herpes Simplex Virus Infection<br>Refractory Viral Keratitis | Drug: HELP (BD111) Adult single group Dose<br>(Administered by corneal injection surgery. Dosage form: injection solution. Dose: 250μL. Frequency of administration: one time injection.) |

**Description:**

This is an open-label, single dose study of HELP in adult (ages 18 to 70) participants with refractory herpetic viral keratitis. Six participants will be enrolled. HELP is a novel gene editing product designed to clear Herpes simplex virus type I (HSV-1) that results in herpetic stromal keratitis in both acute and recurrent infection models which is the leading factor for infectious blindness.

The follow-up period was 360 days, and the patients will be followed up 3±1 days, 7±2 days, 30±7 days, 90±14 days, 180±21 days, and 360+31 days after treatment.

**Outcomes Measures:**

- **<u>Primary Outcome Measure</u>**

1. Concentration of dose limiting toxicities

Observe and record AE, SAE incidence of dose limiting toxicities related with HELP administration.

2. Concentration of maximum tolerated dose

Observe and record AE, SAE at maximum tolerated dose when occurs dose limiting toxicities.

3. Clearance effective of HSV-1 genome

Judge HSV-1 genome clearance effective according to DNA sequencing results by methods of Plaque assay, Elisa, PCR etc.

4. Grafted corneal survival time

Observe the survival time of grafted corneal in subjects, whether the grafted corneal is transparent or opacity.

5. Visual improvement compared with baseline

Judge the visual recovery progress according to visual examination results on day 3±1，7±2，30±7，90±14，180±21，360±31.

- **Secondary Outcome Measure**

1. Changes of herpes virus load compared with baseline

Herpes virus content before and after treatment were determined by methods of plaque assay, ELISA, PCR etc. Compare the viral content changes with baseline.

5.1 Quality Control and Quality Assurance………………………………………...16

5.2 Monitoring…………………………………………………………………….16

**6. Data Collection and Evaluation**………………………………………………………..17

**7. Statistical Considerations**……………………………………………………………...18

10.1 Ethical Conduct of the Study……………………………………………….……...20

10.2 Patient Information and Consent…………………………………………….….20

**11. Reference**…………………………………………..…………………..………..………22

### 1. Introduction

#### 1.1 Study Objectives

The primary objective of this study is to determine the safety of CRISPR/Cas9 mRNA Instantaneous Gene Editing Therapy in the form of intracorneal injected HELP. The secondary objective is to evaluate the HSV-1 erasing efficiency of HELP with the assistance of corneal transplantation for the treatment of refractory HSK patients. In addition, potential long-term off-target effect of HELP injection should also be determined during the 360 days of follow-ups.

#### 1.2 Backgrounds

HELP, a mRNA-carrying lentiviral particles that include SpCas9 mRNA and HSV-1-gene-targeting guide RNAs (designated HSV-1-erasing lentiviral particles, termed HELP). The guide RNA expression cassette simultaneously were targeted two genes of HSV-1, UL8 and UL29, and co-packaged it with SpCas9 mRNA in an mRNA-carrying lentiviral particle. The gRNA expression cassette is reverse transcribed and maintained as circular episomal DNA, corresponding to an integration-defective lentiviral vector. Lentiviral transporters have been reported increasingly in terms of gene editing. As reported in other studies,[1-5] only transient, mild ocular inflammation following surgery was noted and the presence of vector was limited to the injected eye after treatment.

### 2. Eligibility Criteria

#### 2.1 Inclusion Criteria

Patients with refractory keratitis caused by herpes virus type I who has acute corneal perforation or had at least one time failed corneal transplant due to the virus relapse.

- Age between 18 to 75 years
- No systemic immune eye disease
- Good eyelid structure and blink function
- Exists the potential of visual recovery by evaluation of ocular structure and function

- Identify patients who are repeatedly infected with HSV-1 virus(only under the condition that the patients are obviously suffering relapse infections, corneal perforation and need corneal transplantation)
- No retinal detachment and normal visual function
- No history of corneal trauma
- Subjects or their legal guardians voluntarily participate in this study, sign informed consent, good compliance and cooperation with follow-up visits

**2.2 Exclusion Criteria**

- Lacrimal coating and blink function loss
- Schirmer's test result is less than 2mm for severe dry eye disease
- Pregnant and lactating women (pregnancy defined in this study as positive urine pregnancy test)
- Currently is involved in clinical trials of other drugs or medical devices
- Active eye infection (including but not limited to: blepharitis, infectious conjunctivitis, keratitis, sclerotitis, endophthalmitis) in target eye or contralateral eye within 30 days prior to enrollment
- Ocular surface malignant tumor
- A history of allergic reaction or allergy to sodium luciferin, allergy to protein products used for treatment or diagnosis, allergy to 2 drugs or non-drug factors, or current allergic disease
- current in an infectious disease requiring oral, intramuscular or intravenous administration
- Patients with systemic immune diseases
- Any uncontrolled clinical problems (such as severe mental, neurological, cardiovascular, respiratory and other systemic diseases and malignant neoplasms)
- Not effective contraception
- In uncontrolled hypertension, systolic is no less than 160 mmHg, diastolic is no less than 100 mmHg
- In uncontrolled diabetes, fasting glucose is no less than 10.0umol/L

- Renal insufficiency, serum creatinine is more than 133umol/L
- Arrhythmia, myocardial ischemia, myocardial infarction (diagnosed by electrocardiogram)
- Liver dysfunction, al ANINE aminotransferase and aspartate aminotransferase levels are higher than 80 IU/L
- Platelet level is below 100,000 /uL or above 450,000 /uL
- Hemoglobin level is below 10.0g/dL (male) or 9.0g/dL (female)
- No anticoagulant was used, prothrombin time is higher than 16s, and thrombin time of activated part is higher than 50s
- HIV infection (HIV-positive)
- Subjects lack compliance with the study or the ability to sign informed consent
- There are currently signs of systemic infection, including fever and ongoing antibiotic treatment (in this study, systemic infection was defined as deviation from normal values of white blood cells, lymphocytes, and neutrophils on routine blood tests)
- Administration of Glucocorticoids and other systemic immunosuppressive drugs
- The investigator judges other conditions unsuitable for the trial

### 3. Study Implementation

#### 3.1 Study Design

The enrollment of participants initiates relevant processes immediately. After the completion of preoperative examinations, participants with corneal perforation will be arranged for transplantation as soon as possible. During the usual procedure of the surgery, CRISPR/Cas9 mRNA formulation in the form of HELP will be administrated for the treatment of refractory HSK. The hospital observations of participants will persist for 7 days. The postoperative recovery condition, HSV-1 testing outcome, adverse effects of the participants will be examined at every follow-up time. After their hospital discharge on 7th day, follow-up will be conducted on 1 month, 2 months, 3 months, 6 months, and 12 months after the surgery. The basic information is shown in the Table 1. below.

**Table 1. Basic Information of Our Clinical Study**

| Item | Content |
|---|---|
| Study Type | Interventional (Clinical Trial) |
| Estimated Enrollment | 6 participants |
| Allocation | N/A |
| Intervention Model | Single Group Assignment |
| Intervention Model Description | Single group target value: Since there is no similar products approved on the market or similar comparative treatment methods, the single group target value method is adopted |
| Masking | None (Open Label) |
| Primary Purpose | Treatment |
| Actual Study Start Date | November 4, 2020 |
| Estimated Primary Completion Date | November 2021 |
| Estimated Study Completion Date | May 2022 |

In this study, each participant diagnosed with terminal refractory HSK will undergo HELP injection during the normal procedure of penetrating keratoplasty (PK). Before the surgery, they will receive conventional topical or systemic antiviral treatment. During the operation, single dose CRISPR/Cas9 mRNA formulation in the form of HELP will be injected into the recipient graft bed. Then the participants will no longer receive any forms of antiviral treatment unless occurs an uncontrollable virus recurrence, when the conventional antiviral therapy continues. At follow-up visits, participants will undergo a series of examinations, including best-corrected visual acuity, intraocular pressure, photograph of anterior segment, postoperative B-scan ultrasonography, time-domain optical coherence tomography (OCT), spectral-domain OCT, electroretinography (ERG), ELISA of blood samples, deep sequencing of corneal epithelial cells, HSV-1 assessment and potential adverse events.

##### 3.2 Preparation of HELP Formulation

HELP (BD111) was produced following Good Manufacturing Practice (GMP) guidelines. Briefly, 293T cells were seeded in 10-tray stacks cell factory (Thermo Fisher Scientific) and transfected with 6 packaging plasmids using PEI. Cell culture medium was refreshed 6 h post-transfection.

Supernatants were harvested 48 h later and pooled before filtration through 0.45 μm. The downstream purification process included a benzonase treatment overnight at 4°C, followed by ultrafiltration and purification using ion-exchange chromatography. The resulting BD111 was dissolved in PBS, passed through 0.22-μm filters, and stored at -80℃.

#### 3.3 Corneal Transplant and HELP Administration

##### 3.3.1 Preoperative Medications

The preoperative medications for participants having a PK are listed below:

- Topical antiviral drugs: Acyclovir eye drops 6 times daily and oral acyclovir tablets 3 times daily till the day before surgery
- Miotics: 1% Pilocarpine drops for phakic patients undergoing PK, instilled every 10 minutes for 30 minutes before the operation
- Intravenous Mannitol 20%: Dosage of 1 g/kg given over 20–30 minutes before the operation to lower the intraocular pressure

##### 3.3.2 Anesthesia

PK will be performed under retrobulbar block for patients with good cooperation or those who are at risk for undergoing general endotracheal anesthesia in our hospital. The procedure is performed under monitored anesthesia care, including the control of blood pressure, heart rate, oxygen saturation and anxiety. Patients should supine and look straight forward, with their heads maintained in a neutral position. After sterilization of the surgical area, a needle (27 Gauge, 3 cm long) is inserted at the inferolateral border of the bony orbit and directed straight back until its tip has passed the equator of the globe. The needle is then directed medially and cephalad toward the apex of the orbit. Upon a negative aspiration for blood, 3-4mls of local anesthetic solution is injected and the needle is then withdrawn. 2% Lidocaine (Xylocaine) is used as anesthetic agent. Patients are then draped in sterile fashion using a surgical film, which isolates eyelashes from the sterile field.

##### 3.3.3 Procedural Technique

After the placement of a lid speculum, an appropriately sized trephine (usually 6.5-7.5mm) marks the epithelium on the host cornea prior to trephination. Once centered, trephination to approximately 90% depth is performed. Through the base of the trephination groove enters the anterior chamber using a

75 blade. The cornea is then carefully removed using corneal scissors, avoiding any inadvertently cutting or straying outside of the groove. After removing the cornea, viscoelastic is instilled to maintain the anterior chamber and provide protection to the intraocular tissues. The removed host tissue will be sent to pathology for pathogen detection.

The donor corneal button is cut endothelial side up using a trephine-loaded punch instrument, which should be completed prior to trephining the host. In globe perforation participants, donor grafts should be oversized by 0.5-0.75 mm compared with the host bed. Once the donor button is cut, the tissue is carefully protected for later use. Before transferring to the recipient bed, the donor graft is irrigated to remove storage solution. 10-0 nylon suture and interrupted sutures are preferred for securing the donor tissue to the host bed. Initially 4 interrupted cardinal sutures in sequence at 12, 6, 3, 9 o'clock position are performed, and 8-12 more sutures may be necessary before the graft is secure. Loose ends of the sutures should be cut short and the knots be buried in the host tissue. Finally, additional viscoelastic is applied in the angle and to the anterior chamber.

##### 3.3.4 HELP Administration

The HELP formulation stored in -80°C will be transferred in 4 °C from the lab to the surgery room 30 min earlier. Then it will be recovered to the room temperature 10 min before its usage, where in total of 0.25 ml formulation is drawn into a 27g ophthalmic syringe for the later injection. Penetrating keratoplasty will be performed following the routine procedures described above. After sewing up the donor cornea, the HELP formulation in the syringe is injected in evenly divided 6-8 locations of the recipient graft bed. The indication for its successful entrance into the stroma will be the visible edema and whitening of the corneal bed in the injection sites, which should cover the entire recipient bed area in the end.

#### 3.4 Post-operation Management

- Antimicrobials: for an infectious cause, topical antimicrobials should be selected based on cultures after PK. Initially, the antibiotic should be broad spectrum with low toxicity (e.g. fluoroquinolones), typically four times daily during the first week
- Immune Modulators: topical corticosteroid drops (such as difluprednate 0.05% ophthalmic suspension or prednisolone acetate 1.0% ophthalmic suspension four times daily (average). Steroids are tapered depending on the patient

- Intraocular Pressure Lowering Agents: intraocular Pressure should be monitored at each postoperative visit, agents for lowering pressure should be used if significantly elevated
- Ocular Lubricants: non-preserved artificial tears can be started early in the postoperative period to maintain and preserve the corneal epithelium

#### 4. CRISPR/Cas9 Therapy Interruption

A CRISPR/Cas9 therapy interruption is a restart of discontinued conventional anti-viral therapy, since the CRISPR gene editing therapy is single dose applied. The CRISPR/Cas9 therapy interruption will be performed under certain conditions. As HSV-1 testing result is the main outcome of this study, the CRISPR/Cas9 therapy interruption will only be performed when HSV-1 testing result is at a certain range.

##### 4.1 Criteria for CRISPR/Cas9 Therapy Interruption

- A written informed consent was provided by the patient
- HSV-1 testing results rises to excessive range (Ct value <25)
- Noticeable corneal or systemic adverse events related to CRISPR/Cas9 therapy

#### 5. Quality Control

##### 5.1 Quality Control and Quality Assurance

During this study, the Emergency Response Group will conduct regular visits to ensure the protocol and Gond Clinical Practices (GCPs) being followed. The monitors may inspect the accuracy of the recorded data through source documents. The investigators and their relevant personnel are available and cooperate during the monitoring visits and possible audits or inspections.

##### 5.2 Monitoring

The potential audits and inspections, including source data verification, must be accepted by representatives designated by the Protocol Chair. Through these audits and inspections, monitors will examine whether the medical activities and documents related to this study were following the protocol, institutional regulation, and any applicable policy requirements. Monitoring will start at the first time of participant registration and will continue during the entire protocol performance.

#### 6. Data Collection and Evaluation

Unless noted, the procedures for all patients must be identical. Any AE/SAE will be documented and reported throughout the study. The period of data collection will be divided into three sections, including pre-study screening, study procedure and post-study follow-up. Screening procedures for baseline data will be taken 3~14 days prior to CRISPR/Cas9 gene editing treatment. An, and institutional Review Board (IRB) approved informed consent document (ICD), must be signed and dated before any specific procedures are done. Results from standard clinical assessments or laboratory tests, prior to the date of informed consent but within the allowed time restraint for screening procedures, can be used for determining the patient's eligibility. The patient's source documents must clearly document the use of pre-consent results. A screening will contain the following items:

- Sign form of informed consent
- Disease history and prior treatment
- Complete ophthalmic examinations
- Vital signs
- Laboratory evaluation
- Other necessary items depending on specific situations

During the study period, each patient will receive corresponding treatment according to the protocol, which was elaborated in the “Study Implementation” section. After the procedure, all the patients will be scheduled a follow-up for 360 days or more, the patients will be followed up 7±2 days, 30±7 days, 60±10 days 90±14 days, 180±21 days and 360±31 days after treatment, where their post-treatment status will be carefully evaluated.

### 7. Statistical Considerations

This is an open-label, single-arm clinical study. The investigators conducted this research to evaluate the safety and efficacy of HSV-1-targeting CRISPR formulation HELP (BD111) for in vivo treatment of refractory HSK patients. In this study, participants will receive a single dose of HELP formulation by intrastromal injection during the penetrating keratoplasty for acute corneal perforation. At each follow-up point, a series of safety and effectiveness evaluations will be performed.

The primary endpoint, HSV-1 in aqueous humour, corneal button, tear swabs will be evaluated at various test points according to the data collection plan, as summarized by simple descriptive summary. For safety evaluations, clinical examinations (slit-lamp examination, measurement of best-corrected visual acuity, intraocular pressure, B-scan ultrasonography, the time-domain OCT, the spectral-domain OCT, and the ERG) will be presented as simple descriptive statistics in the form of pictures, line charts or summarized tables. Deep sequencing of the corneal epithelial cells (acquired by corneal wipes) will also be reported as descriptively summarized tables. Blood ELISA will be reported graphically with the aid of the Student's test. Other laboratory and imaging correlative studies will also be reported descriptively and graphically.

### 8. Adverse Event Reporting

#### 8.1 Adverse Event

AE is defined as an undesirable experience or medical condition that occurs after the administration of the procedures or operations specified in the protocol. AE includes all events that may or may not be related to the study. All observed or volunteered AEs regardless of suspected causal relationship to the CRISPR/Cas9 gene editing treatment will be reported as described in the following sections. For all AEs, the investigator must obtain adequate information both to evaluate the outcome of AEs and to assess whether it fits the criteria as a SAE, which requires immediate notification to designated representatives. Follow-ups by the investigator are also required until the event stabilizes at an acceptable level to the investigator.

#### 8.2 Serious Adverse Event Reporting

SAE is an AE with a potential outcome of death, prolonged hospitalization, disability or permanent damage, congenital anomaly/birth defect, or other serious important medical events. Specifically, SAE is any untoward medical occurrence that:

- Results in death
- Life-threatening
- Requires extra hospitalization or prolongation of existing hospitalization
- Results in persistent or significant disability/incapacity
- Results in congenital anomaly/birth defect

All suspected SAEs related to the study should be followed up according to the institution's standard of care, preferably in consultation with leading specialists.

##### 8.3 Recording and Reporting

An emergency contact will be assigned to each participant in case of any unexpected circumstances. In occurrence of an AE, it will be graded according to the DAIDS severity grading system ICH 6:

- Mild: participant was able to perform all normal activities
- Moderate: the participant had to discontinue some activities due to the AE
- Severe: the participant was incapacitated by the AE and unable to perform normal activities
- Life-threatening: participant experienced extreme limitation in activity, significant assistance required; significant medical intervention/therapy required, hospitalization probable

Investigators will assess the occurrence of AEs and SAEs at all follow up time points during the study. All AE/SAE will be recorded and reported in detail, with further investigation into the relationship between each AE and the clinical procedures.

#### 9. Record Form & Retention

All data related to this study in both paper and electronic form will be available only to the research team and the legal supervisors. The original data are the property of the investigation team and are inaccessible to any other third parties. The researchers from the team are ultimately responsible for the collection and analyze of the data. Any alterations to the original data must be documented. All the original data will be retained securely by the investigators both during and after the closure of the study in order to ensure evaluations or audits from regulatory authorities. If the investigator is unable to continue retaining the study records for the required period for any reason, the Emergency Response Group should be prospectively notified.

#### 10.Ethics

The conduct of this study will abide by the laws and principles proclaimed in the Declaration of Helsinki. All managements and operations will be carried out under the supervision of The Institutional Review Board (IRB) & Hospital Ethics Committee (HEC).

##### 10.1 Ethical Conduct of the Study

The study will be conducted in accordance with the protocol and the requirements according to the concerned laws. The project will be submitted to the supervision parties for censorship. The study will be initiated only with the approval of the IRB & HEC. All the data and information collected from the study will be presented to the supervision parties, especially the information related to AE & SAE. Alteration of the project plan will not be made without the consideration and authorization of the supervision parties. A full report will be provided after the closure of the study.

##### 10.2 Patient Information and Consent

All participants and/or their legal representatives will be fully informed of the objectives and details of this study. All possible outcomes and risks will be acknowledged. A consent of written form will be obtained from each patient or their representatives. The consent process will be witnessed by an impartial supervisor. A copy of the consent will be provided if requested by the participants. The consent documents will be stored by the investigators along with other data collected during the study for a period required by the protocol. All the personal information and data from the patients will only be available to the investigators and supervision parties to guarantee privacy. The data will not be used for purposes other than this study.

### Current Study Protocol

**Protocol ID: 2020023-2-v3**

### In Vivo CRISPR Gene Editing in a Patient with Herpes Stromal Keratitis

Investigator: Jiaxu Hong, M.D., Ph.D, M.PA.
Deputy Chief Physician
Department of Ophthalmology and Visual Science

Research Institute: Eye, and ENT Hospital,
Shanghai Medical College, Fudan University, Shanghai, China

Founding: National Natural Science Foundation of China,
81970766, 8217040684 and 31971364
the Program for Professor of Special Appointment
(Eastern Scholar) at Shanghai Institutions of Higher Learning,
TP2019046
The Shanghai Innovation Development Program,
2020-RGZN-02033
the Shanghai Key Clinical Research Program,
SHDC2020CR3052B
Startup funding from Shanghai Center for Systems Biomedicine
Shanghai Jiao Tong University, WF220441504

Collaborators: Key Laboratory of Systems Biomedicine,
Shanghai Jiao Tong University

Version 3.0

Version Date: July 13, 2021

**Brief Summary:**

The purpose of this study is to evaluate the safety, tolerability and efficacy of a single dose of CRISPR/Cas9 mRNA Instantaneous Gene Editing Therapy administered via corneal injection in participants with refractory herpetic viral keratitis.

| Condition or disease | Intervention/treatment |
|---|---|
| Herpes Simplex Virus Infection<br>Refractory Viral Keratitis | Drug: HELP (BD111) Adult single group Dose (Administered by corneal injection surgery. Dosage form: injection solution. Dose: 200μL. Frequency of administration: one time injection.) |

**Description:**

This is a single-arm, single dose study of mRNA-carrying HELP (HSV-1-erasing lentiviral particles) in adult (ages 18 to 70) participants with refractory herpetic viral keratitis. Approximately 6 participants will be enrolled and receive HELP injection during the usual procedure of corneal transplantation. HELP is a novel gene editing formulation designed to clear Herpes simplex virus type I (HSV-1) that results in herpetic stromal keratitis in both acute and recurrent infection models which is the leading factor for infectious blindness. The main outcomes of this study include HSV-1 testing outcome, graft survival, visual acuity and adverse effects of the participants.

**Outcomes Measures:**

1. Primary Outcome Measure

- HSV-1 testing results of the intervention eye. Tear swab of 1 week, 1 month, 2 months, 3 months, 6 months and 12 months post-operation will be collected and quantified for HSV-1 genome by real-time polymerase chain reaction (RT-PCR). The virus testing outcome was indicated as positive (+) or negative (-) by Ct values. Ct value of above 40 will be considered negative, 36-40 be considered weakly positive, and below 35 will be considered a strong positive.

- Adverse effects, including possible alteration of ocular structure and function, Cas9-specific immunoreaction or off-target editing of CRISPR in the bloodstream. Postoperative B-scan ultrasonography, time-domain OCT, spectral-domain OCT, and electroretinography (ERG) will be used to evaluate ocular status. ELISA and Whole-genome sequencing (WGS) will be conducted on the blood samples collected at 3-month and 6-month visit, respectively.

2. Secondary Outcome Measure

- Patient graft survival. The corneal grafts of participants will be monitored under a complete ophthalmic examination, including slit-lamp examination and anterior segment optical coherence tomography (OCT), at every follow up visit.
- Best-corrected visual acuity. Changes of visual acuity of participants along the clinical trial.

10.1 Ethical Conduct of the Study……………………………………………….……...39

10.2 Patient Information and Consent……………………………………………….….39

**11. Reference**…………………………………………….…………………….……….……….………41

### 1. Introduction

#### 1.1 Study Objectives

The primary objective of this study is to evaluate the HSV-1 erasing efficiency of HELP with the assistance of corneal transplantation for the treatment of refractory HSK patients. The secondary objective is to determine the safety of CRISPR/Cas9 mRNA Instantaneous Gene Editing Therapy in the form of intracorneal injected HELP. In addition, potential long-term off-target effect of HELP injection should also be determined during the 360 days of follow-ups.

#### 1.2 Backgrounds

HELP, a mRNA-carrying lentiviral particles that include SpCas9 mRNA and HSV-1-gene-targeting guide RNAs (designated HSV-1-erasing lentiviral particles, termed HELP). The guide RNA expression cassette simultaneously were targeted two genes of HSV-1, UL8 and UL29, and co-packaged it with SpCas9 mRNA in an mRNA-carrying lentiviral particle. The gRNA expression cassette is reverse transcribed and maintained as circular episomal DNA, corresponding to an integration-defective lentiviral vector. Lentiviral transporters have been reported increasingly in terms of gene editing. As reported in other studies,[1-5] only transient, mild ocular inflammation following surgery was noted and the presence of vector was limited to the injected eye after treatment.

### 2. Eligibility Criteria

#### 2.1 Inclusion Criteria

- Age between 18 to 70 years old;
- No systemic immune eye disease;
- Good eyelid structure and blink function;
- Exists the potential of visual recovery by evaluation of ocular structure and function;
- Patients with refractory keratitis who are repeatedly infected with HSV-1 virus (more than three times per year) and resistant to topical or systemic anti-viral agents, with no response to regular immunosuppressive agents;

- Patients who are obviously suffering from relapse HSV infections with corneal perforation, requiring corneal transplantation;
- No history of corneal trauma;
- Subjects or their legal guardians voluntarily participate in this study, sign informed consent, good compliance and cooperation with follow-up visits.

**2.2 Exclusion Criteria**

- Lacrimal coating and blink function loss;
- Schirmer's test result is less than 2mm for severe dry eye disease;
- Pregnant and lactating women (pregnancy defined in this study as positive urine pregnancy test);
- Currently is involved in clinical trials of other drugs or medical devices;
- Active eye infection (including but not limited to: blepharitis, infectious conjunctivitis, keratitis, sclerotitis, endophthalmitis) in target eye or contralateral eye within 30 days prior to enrollment;
- Ocular surface malignant tumor;
- A history of allergic reaction or allergy to sodium luciferin, allergy to protein products used for treatment or diagnosis, allergy to ≥ 2 drugs or non-drug factors, or current allergic disease;
- current in an infectious disease requiring oral, intramuscular or intravenous administration;
- Patients with systemic immune diseases;
- Any uncontrolled clinical problems (such as severe mental, neurological, cardiovascular, respiratory and other systemic diseases and malignant neoplasms);
- Not effective contraception;
- In uncontrolled hypertension, systolic is no less than 160 mmHg, diastolic is no less than 100 mmHg;
- In uncontrolled diabetes, fasting glucose is no less than 10.0umol/L;
- Renal insufficiency, serum creatinine is more than 133umol/L;
- Arrhythmia, myocardial ischemia, myocardial infarction (diagnosed by electrocardiogram);

- Liver dysfunction, al ANINE aminotransferase and aspartate aminotransferase levels are higher than 80 IU/L;
- Platelet level is below 100,000 /μL or above 450,000 /μL;
- Hemoglobin level is below 10.0g/dL (male) or 9.0g/dL (female);
- No anticoagulant was used, prothrombin time is higher than 16s, and thrombin time of activated part is higher than 50s;
- HIV infection (HIV-positive);
- Subjects lack compliance with the study or the ability to sign informed consent;
- There are currently signs of systemic infection, including fever and ongoing antibiotic treatment (in this study, systemic infection was defined as deviation from normal values of white blood cells, lymphocytes, and neutrophils on routine blood tests);
- Administration of Glucocorticoids and other systemic immunosuppressive drugs;
- The investigator judges other conditions unsuitable for the trial.

#### 3. Study Implementation

##### 3.1 Study Design

The enrollment of participants initiates relevant processes immediately. After the completion of preoperative examinations, participants with corneal perforation will be arranged for transplantation as soon as possible. During the usual procedure of the surgery, CRISPR/Cas9 mRNA formulation in the form of HELP will be administrated for the treatment of refractory HSK. The hospital observations of participants will persist for 7 days. The postoperative recovery condition, HSV-1 testing outcome, adverse effects of the participants will be examined at every follow-up time. After their hospital discharge on 7th day, follow-up will be conducted on 1 month, 2 months, 3 months, 6 months and 12 months after the surgery. The basic information is shown in the Table 1. below.

**Table 1. Basic Information of Our Clinical Study**

| Item | Content |
|---|---|
| Study Type | Interventional (Clinical Trial) |
| Estimated Enrollment | Six participants |
| Allocation | N/A |
| Intervention Model | Single Group Assignment |
| Intervention Model Description | Single group target value: Since there is no similar products approved on the market or similar comparative treatment methods, the single group target value method is adopted |
| Masking | None (Open Label) |
| Primary Purpose | Treatment |
| Actual Study Start Date | November 4, 2020 |
| Estimated Primary Completion Date | November 2021 |
| Estimated Study Completion Date | May 2022 |

In this study, each participant diagnosed with terminal refractory HSK will undergo HELP injection during the normal procedure of penetrating keratoplasty (PK). Before the surgery, they will receive conventional topical or systemic antiviral treatment. During the operation, single dose CRISPR/Cas9 mRNA formulation in the form of HELP will be injected into the recipient graft bed. Then the participants will no longer receive any forms of antiviral treatment unless occurs an uncontrollable virus recurrence, when the conventional antiviral therapy continues. At follow-up visits, participants will undergo a series of examinations, including best-corrected visual acuity, intraocular pressure, photograph of anterior segment, postoperative B-scan ultrasonography, time-domain optical coherence tomography (OCT), spectral-domain OCT, electroretinography (ERG), ELISA of blood samples, deep sequencing of corneal epithelial cells, HSV-1 testing and potential adverse events.

##### 3.2 Preparation of HELP Formulation

HELP (BD111) was produced following Good Manufacturing Practice (GMP) guidelines. Briefly, 293T cells were seeded in 10-tray stacks cell factory (Thermo Fisher Scientific) and transfected with 6 packaging plasmids using PEI. Cell culture medium was refreshed 6 h post-transfection.

Supernatants were harvested 48 h later and pooled before filtration through 0.45 μm. The downstream purification process included a benzonase treatment overnight at 4°C, followed by ultrafiltration and purification using ion-exchange chromatography. The resulting BD111 was dissolved in PBS, passed through 0.22-μm filters, and stored at -80℃.

#### 3.3 Corneal Transplant and HELP Administration

##### 3.3.1 Preoperative Medications

The preoperative medications for participants having a PK are listed below:

- Topical antiviral drugs: Acyclovir eye drops 6 times daily and oral acyclovir tablets 3 times daily till the day before surgery
- Miotics: 1% Pilocarpine drops for phakic patients undergoing PK, instilled every 10 minutes for 30 minutes before the operation
- Intravenous Mannitol 20%: Dosage of 1 g/kg given over 20–30 minutes before the operation to lower the intraocular pressure

##### 3.3.2 Anesthesia

PK will be performed under retrobulbar block for patients with good cooperation or those who are at risk for undergoing general endotracheal anesthesia in our hospital. The procedure is performed under monitored anesthesia care, including the control of blood pressure, heart rate, oxygen saturation and anxiety. Patients should supine and look straight forward, with their heads maintained in a neutral position. After sterilization of the surgical area, a needle (27 Gauge, 3 cm long) is inserted at the inferolateral border of the bony orbit and directed straight back until its tip has passed the equator of the globe. The needle is then directed medially and cephalad toward the apex of the orbit. Upon a negative aspiration for blood, 3-4mls of local anesthetic solution is injected and the needle is then withdrawn. 2% Lidocaine (Xylocaine) is used as anesthetic agent. Patients are then draped in sterile fashion using a surgical film, which isolates eyelashes from the sterile field.

##### 3.3.3 Procedural Technique

After the placement of a lid speculum, an appropriately sized trephine (usually 6.5-7.5mm) marks the epithelium on the host cornea prior to trephination. Once centered, trephination to approximately 90% depth is performed. Through the base of the trephination groove enters the anterior chamber using a

75 blade. The cornea is then carefully removed using corneal scissors, avoiding any inadvertently cutting or straying outside of the groove. After removing the cornea, viscoelastic is instilled to maintain the anterior chamber and provide protection to the intraocular tissues. The removed host tissue will be sent to pathology for pathogen detection.

The donor corneal button is cut endothelial side up using a trephine-loaded punch instrument, which should be completed prior to trephining the host. In globe perforation participants, donor grafts should be oversized by 0.5-0.75 mm compared with the host bed. Once the donor button is cut, the tissue is carefully protected for later use. Before transferring to the recipient bed, the donor graft is irrigated to remove storage solution. 10-0 nylon suture and interrupted sutures are preferred for securing the donor tissue to the host bed. Initially 4 interrupted cardinal sutures in sequence at 12, 6, 3, 9 o'clock position are performed, and 8-12 more sutures may be necessary before the graft is secure. Loose ends of the sutures should be cut short and the knots be buried in the host tissue. Finally, additional viscoelastic is applied in the angle and to the anterior chamber.

##### 3.3.4 HELP Administration

The HELP formulation stored in -80°C will be transferred in 4 °C from the lab to the surgery room 30 min earlier. Then it will be recovered to the room temperature 10 min before its usage, where in total of 0.25 ml formulation is drawn into a 27g ophthalmic syringe for the later injection. Penetrating keratoplasty will be performed following the routine procedures described above. After sewing up the donor cornea, the HELP formulation in the syringe is injected in evenly divided 6-8 locations of the recipient graft bed. The indication for its successful entrance into the stroma will be the visible edema and whitening of the corneal bed in the injection sites, which should cover the entire recipient bed area in the end.

#### 3.4 Post-operation Management

- Antimicrobials: for an infectious cause, topical antimicrobials should be selected based on cultures after PK. Initially, the antibiotic should be broad spectrum with low toxicity (e.g. fluoroquinolones), typically four times daily during the first week
- Immune Modulators: topical corticosteroid drops (such as difluprednate 0.05% ophthalmic suspension or prednisolone acetate 1.0% ophthalmic suspension four times daily (average). Steroids are tapered depending on the patient

- Intraocular Pressure Lowering Agents: intraocular Pressure should be monitored at each postoperative visit, agents for lowering pressure should be used if significantly elevated
- Ocular Lubricants: non-preserved artificial tears can be started early in the postoperative period to maintain and preserve the corneal epithelium

#### 4. CRISPR/Cas9 Therapy Interruption

A CRISPR/Cas9 therapy interruption is a restart of discontinued conventional anti-viral therapy, since the CRISPR gene editing therapy is single dose applied. The CRISPR/Cas9 therapy interruption will be performed under certain conditions. As HSV-1 testing results is the main outcome of this study, the CRISPR/Cas9 therapy interruption will only be performed when virus testing outcome is at a certain range.

##### 4.1 Criteria for CRISPR/Cas9 Therapy Interruption

- A written informed consent was provided by the patient
- HSV-1 testing outcome rises to excessive range (Ct value <25)
- Noticeable corneal or systemic adverse events related to CRISPR/Cas9 therapy

#### 5. Quality Control

##### 5.1 Quality Control and Quality Assurance

During this study, the Emergency Response Group will conduct regular visits to ensure the protocol and Gond Clinical Practices (GCPs) being followed. The monitors may inspect the accuracy of the recorded data through source documents. The investigators and their relevant personnel are available and cooperate during the monitoring visits and possible audits or inspections.

##### 5.2 Monitoring

The potential audits and inspections, including source data verification, must be accepted by representatives designated by the Protocol Chair. Through these audits and inspections, monitors will examine whether the medical activities and documents related to this study were following the protocol, institutional regulation, and any applicable policy requirements. Monitoring will start at the first time of participant registration and will continue during the entire protocol performance.

#### 6. Data Collection and Evaluation

Unless noted, the procedures for all patients must be identical. Any AE/SAE will be documented and reported throughout the study. The period of data collection will be divided into three sections, including pre-study screening, study procedure and post-study follow-up. Screening procedures for baseline data will be taken 3~14 days prior to CRISPR/Cas9 gene editing treatment. An, and institutional Review Board (IRB) approved informed consent document (ICD), must be signed and dated before any specific procedures are done. Results from standard clinical assessments or laboratory tests, prior to the date of informed consent but within the allowed time restraint for screening procedures, can be used for determining the patient's eligibility. The patient's source documents must clearly document the use of pre-consent results. A screening will contain the following items:

- Sign form of informed consent
- Disease history and prior treatment
- Complete ophthalmic examinations
- Vital signs
- Laboratory evaluation
- Other necessary items depending on specific situations

During the study period, each patient will receive corresponding treatment according to the protocol, which was elaborated in the “Study Implementation” section. After the procedure, all the patients will be scheduled a follow-up for 360 days or more, the patients will be followed up 7±2 days, 30±7 days, 60±10 days 90±14 days, 180±21 and 360±31 days after treatment, where their post-treatment status will be carefully evaluated.

#### 7. Statistical Considerations

This is an open-label, single-arm clinical study. The investigators conducted this research to evaluate the safety and efficacy of HSV-1-targeting CRISPR formulation HELP for in vivo treatment of refractory HSK patients. In this study, participants will receive a single dose of HELP formulation by intrastromal injection during the penetrating keratoplasty for acute corneal perforation. At each follow-up point, a series of safety and effectiveness evaluations will be performed.

The primary endpoint, HSV-1 in aqueous humour, corneal button, tear swabs will be evaluated at various test points according to the data collection plan, as summarized by simple descriptive summary. For safety evaluations, clinical examinations (slit-lamp examination, measurement of best-corrected visual acuity, intraocular pressure, B-scan ultrasonography, the time-domain OCT, the spectral-domain OCT, and the ERG) will be presented as simple descriptive statistics in the form of pictures, line charts or summarized tables. Deep sequencing of the corneal epithelial cells (acquired by corneal wipes) will also be reported as descriptively summarized tables. Blood ELISA will be reported graphically with the aid of the Student's test. Other laboratory and imaging correlative studies will also be reported descriptively and graphically.

#### 8. Adverse Event Reporting

##### 8.1 Adverse Event

AE is defined as an undesirable experience or medical condition that occurs after the administration of the procedures or operations specified in the protocol. AE includes all events that may or may not be related to the study. All observed or volunteered AEs regardless of suspected causal relationship to the CRISPR/Cas9 gene editing treatment will be reported as described in the following sections. For all AEs, the investigator must obtain adequate information both to evaluate the outcome of AEs and to assess whether it fits the criteria as a SAE, which requires immediate notification to designated representatives. Follow-ups by the investigator are also required until the event stabilizes at an acceptable level to the investigator.

##### 8.2 Serious Adverse Event Reporting

SAE is an AE with a potential outcome of death, prolonged hospitalization, disability or permanent damage, congenital anomaly/birth defect, or other serious important medical events. Specifically, SAE is any untoward medical occurrence that:

- Results in death
- Life-threatening
- Requires extra hospitalization or prolongation of existing hospitalization
- Results in persistent or significant disability/incapacity
- Results in congenital anomaly/birth defect

All suspected SAEs related to the study should be followed up according to the institution's standard of care, preferably in consultation with leading specialists.

**8.3 Recording and Reporting**

An emergency contact will be assigned to each participant in case of any unexpected circumstances. In occurrence of an AE, it will be graded according to the DAIDS severity grading system ICH 6:

- Mild: participant was able to perform all normal activities
- Moderate: the participant had to discontinue some activities due to the AE
- Severe: the participant was incapacitated by the AE and unable to perform normal activities
- Life-threatening: participant experienced extreme limitation in activity, significant assistance required; significant medical intervention/therapy required, hospitalization probable

Investigators will assess the occurrence of AEs and SAEs at all follow up time points during the study. All AE/SAE will be recorded and reported in detail, with further investigation into the relationship between each AE and the clinical procedures.

#### 9. Record Form & Retention

All data related to this study in both paper and electronic form will be available only to the research team and the legal supervisors. The original data are the property of the investigation team and are inaccessible to any other third parties. The researchers from the team are ultimately responsible for the collection and analyze of the data. Any alterations to the original data must be documented. All the original data will be retained securely by the investigators both during and after the closure of the study in order to ensure evaluations or audits from regulatory authorities. If the investigator is unable to continue retaining the study records for the required period for any reason, the Emergency Response Group should be prospectively notified.

#### 10. Ethics

The conduct of this study will abide by the laws and principles proclaimed in the Declaration of Helsinki. All managements and operations will be carried out under the supervision of The Institutional Review Board (IRB) & Hospital Ethics Committee (HEC).

##### 10.1 Ethical Conduct of the Study

The study will be conducted in accordance with the protocol and the requirements according to the concerned laws. The project will be submitted to the supervision parties for censorship. The study will be initiated only with the approval of the IRB & HEC. All the data and information collected from the study will be presented to the supervision parties, especially the information related to AE & SAE. Alteration of the project plan will not be made without the consideration and authorization of the supervision parties. A full report will be provided after the closure of the study.

##### 10.2 Patient Information and Consent

All participants and/or their legal representatives will be fully informed of the objectives and details of this study. All possible outcomes and risks will be acknowledged. A consent of written form will be obtained from each patient or their representatives. The consent process will be witnessed by an impartial supervisor. A copy of the consent will be provided if requested by the participants. The consent documents will be stored by the investigators along with other data collected during the study for a period required by the protocol. All the personal information and data from the patients will only be available to the investigators and supervision parties to guarantee privacy. The data will not be used for purposes other than this study.

### Summary of Protocol Changes

| **Section of Current protocol** | **Rationale** | **Current Version** |
|---|---|---|
| Outcomes Measures | Adjust the contents and priorities of outcomes | Primary outcome: HSV-1 testing outcome of the intervention eye; adverse effects<br>Secondary outcome: patient graft survival; best-corrected visual acuity |
| 2.1 | Adjust the inclusion criteria | Patients with refractory keratitis caused by herpes virus type I who has acute corneal perforation or had at least one time failed corneal transplant due to the virus relapse. |

### Original and Current Statistical Analysis Plan

#### Statistical Analysis Plan

This is an open-label, single-arm, non-randomized interventional trial. The investigators conducted this research to evaluate the safety and efficacy of HSV-1-targeting CRISPR formulation HELP (BD111) for in vivo treatment of refractory HSK patients. In this study, participants will receive a single dose of HELP formulation by intrastromal injection during the penetrating keratoplasty for acute corneal perforation. At each follow-up point, a series of safety and effectiveness evaluations will be performed.

The primary endpoint, HSV-1 in aqueous humour, corneal button, tear swabs will be evaluated at various test points according to the data collection plan, as summarized by simple descriptive summary. For safety evaluations, clinical examinations (slit-lamp examination, measurement of best-corrected visual acuity, intraocular pressure, B-scan ultrasonography, the time-domain OCT, the spectral-domain OCT, and the ERG) will be presented as simple descriptive statistics in the form of pictures, line charts or summarized tables. Deep sequencing of the corneal epithelial cells (acquired by corneal wipes) will also be reported as descriptively summarized tables. Blood ELISA will be reported graphically with the aid of the Student's test. Other laboratory and imaging correlative studies will also be reported descriptively and graphically. Data were analysed using GraphPad Prism 8 and presented as mean ± SEM in all experiments (n=3). Unpaired two-tailed Student's t-tests were performed to determine the P values (95% confidence interval). n.s., non-significant.
