## Supplementary figures and images for "In Vivo CRISPR Gene Editing in Patients with Herpes Stromal Keratitis"

### CONSORT extension for Pilot and Feasibility Trials Flow Diagram

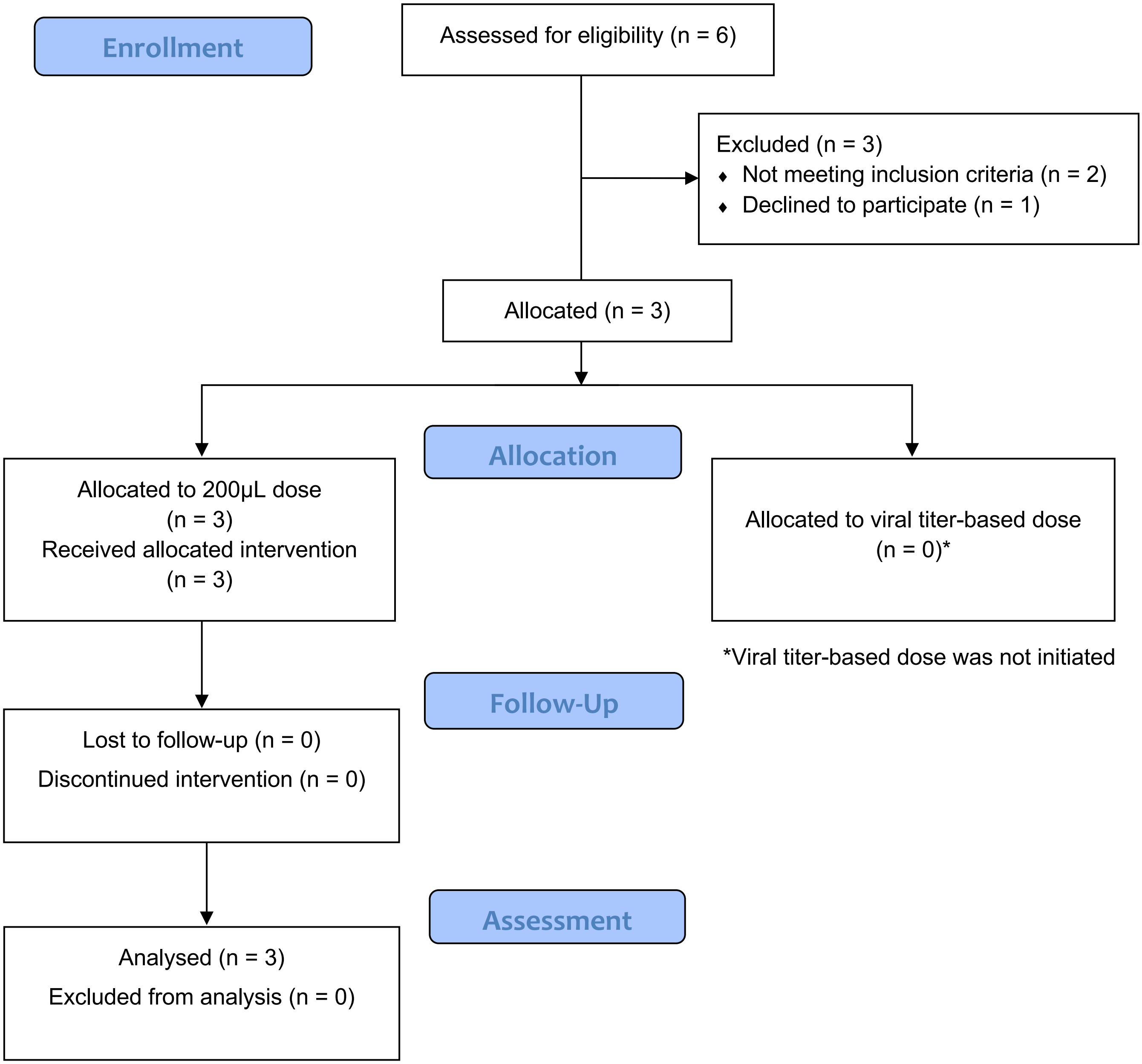
